# Structural brain network organization and aggressive behavior in schizophrenia: a mega-analysis of 2260 patients and 3058 healthy controls from the ENIGMA-Schizophrenia Working Group

**DOI:** 10.64898/2026.10.22.26363656

**Authors:** Jelle Lamsma, Paolo Fusar-Poli, Martijn van den Heuvel, Derin Cobia, Adrian Raine, Bin Wan, Matthias Kirschner, Wiepke Cahn, Dominic Arold, Mihai Avram, Nerisa Banaj, Annarita Barone, Stefan Borgwardt, Katharina Brosch, Rachel M. Brouwer, Arturo Brunetti, Vince D. Calhoun, Sean Carruthers, Giacomo Cecere, Simon Cervenka, Qian H. Chew, Sunah Choi, Young-Chul Chung, Mariateresa Ciccarelli, Sirio Cocozza, Benedicto Crespo-Facorro, Udo Dannlowski, Paola Dazzan, Andrea de Bartolomeis, Marta Di Forti, Stefan Ehrlich, Simone Elia, Asia Ferrari, Marc Fischer, Kira Flinkenflügel, David C. Glahn, Janik Goltermann, Melissa J. Green, Dominik Grotegerd, Oliver Gruber, Amalia Guerrero-Pedraza, Minji Ha, Philipp Homan, L. Elliot Hong, Matthew Hughes, Felice Iasevoli, Stefan Kaiser, Vasily Kaleda, Andriana Karuk, Minah Kim, Tilo Kircher, Peter Kochunov, Bernd Krämer, Jun Soo Kwon, Irina Lebedeva, Maria Lee, Rebekka Lencer, Tiago R. Marques, Susanne Meinert, Robin Murray, Igor Nenadić, Dana Nguyen, Soyolsaikhan Odkhuu, Wolfgang Omlor, Godfrey Pearlson, Rosanne Picotin, Fabrizio Piras, Edith Pomarol-Clotet, Giuseppe Pontillo, Adrian Preda, Rafael Romero-Garcia, Yann Quidé, Elysha Ringin, Amanda Rodrigue, Kelly Rootes-Murdy, Susan L. Rossell, Raymond Salvador, Freda Scheffler, Patricia Segura, Carl M. Sellgren, Kang Sim, Antonin Skoch, Filip Spaniel, Frederike Stein, Philip J. Sumner, Henk Temmingh, Florian Thomas-Odenthal, David Tomecek, Alexander Tomyshev, Mario Tranfa, Jessica A. Turner, Theo G. M. van Erp, Neeltje van Haren, Tamsyn E. Van Rheenen, Jim van Os, Daniela Vecchio, Lei Wang, Will Woods, Thomas Nickl-Jockschat

## Abstract

**Background:** Although most individuals with schizophrenia do not show aggressive behavior, rates are higher than in the general population. Identifying the neurobiological correlates of aggression may inform prevention.

**Methods:** We included 2260 individuals with schizophrenia spectrum disorders and 3058 healthy controls (HC) from 26 sites in the ENIGMA-Schizophrenia Working Group. The former were classified as showing at least moderate aggression (SZ+A; *N* = 231) or not (SZ−A; *N* = 2029) based on symptom rating scales. Structural covariance networks were constructed from cortical and subcortical volumes. We examined global and nodal network topology, as well as density within six large-scale brain networks and between each network and the rest of the brain. Metrics were summarized as the area under the curve (AUC) across a prespecified range of densities (*K* = 0.10–0.25, Δ*K* = 0.01). Group differences were assessed using pairwise permutation tests with Benjamini–Hochberg correction for multiple comparisons.

**Results:** Global and nodal network topology did not differ significantly between SZ+A and HC. Network–rest density of the salience network (SN) was higher in SZ+A than in HC, although this difference did not survive multiple-comparison correction (ΔAUC = 0.018, *p*_BH_ = .083). In exploratory subgroup analyses, higher SN network–rest density was observed only in SZ+A with high delusion severity (ΔAUC = 0.021, *p* = .007). By contrast, SZ−A demonstrated extensive alterations in global network topology compared with SZ+A and HC, alongside selective alterations in nodal topology and large-scale network density compared with HC. Results were broadly consistent across sensitivity analyses using alternative definitions of aggression and adjusting for current antipsychotic dose.

**Conclusions:** SZ+A was characterized by fewer—not more—abnormalities in structural covariance network organization than SZ−A, which may reflect etiological heterogeneity. In SZ+A with high delusion severity, higher network–rest density of the SN may point to aberrant salience processing in aggressive behavior.

## Introduction

Aggression among individuals with schizophrenia represents a major clinical challenge, with substantial consequences for themselves, victims, and mental healthcare systems (1,2). Although uncommon, the risk of aggressive behavior is higher in this population than in the general population (3). Several clinical and environmental risk factors have been identified, including positive symptoms, cognitive impairment, poor illness insight, substance misuse, and social adversity (1,4). However, the neurobiological mechanisms underlying aggression in schizophrenia remain unclear. Elucidating these mechanisms may improve risk stratification, inform preventive interventions, and identify novel therapeutic targets.

Converging evidence from animal, lesion, and neuroimaging studies implicates distributed cortical and subcortical regions in aggressive behavior, including prefrontal, cingulate, parietal, and insular cortices, as well as the hippocampus, amygdala, and striatum (5–7). Many of these regions are key nodes of the salience network (SN), central executive network (CEN), default mode network (DMN), and reward network (RN) (8,9). Several also belong to a network involved in emotion regulation (10–12), referred to here as the affective control network (ACN). However, aggression likely emerges from coordinated interactions throughout the brain rather than from individual regions or large-scale brain networks in isolation (13), highlighting the importance of examining the brain as an interconnected system.

Schizophrenia has long been conceptualized as a disorder of dysconnectivity, in which disrupted interactions among brain regions underlie its symptoms and clinical features (14). Consistent with this conceptualization, widespread abnormalities in gray matter (GM) volume and white matter (WM) microstructural integrity have been reported (15–17), alongside structural and functional abnormalities of the SN, CEN, DMN, RN, ACN, and language network (LN) (18–20). Abnormalities in these networks have been linked to positive symptoms (21–23), cognitive impairment (24–26), and poor insight (27,28). Together, these findings suggest overlap with networks implicated in aggression.

This overlap provides a rationale for examining brain network organization in relation to aggressive behavior in schizophrenia. Graph theory provides a framework for quantifying brain network organization (29). Brain networks can be constructed from magnetic resonance imaging (MRI) data as structural covariance networks (SCNs) based on interregional correlations in GM morphology, structural connectomes based on WM pathways, or functional connectomes based on interregional correlations in brain activity. These network types reflect partly distinct biological processes (30). In schizophrenia, a meta-analysis of global network topology found reduced segregation and integration in structural connectomes but no consistent alterations in functional connectomes (31). At the nodal level, alterations in structural and functional connectomes vary considerably in direction and anatomical distribution (32,33). Findings from the comparatively limited number of SCN studies are mixed across global and nodal metrics (34). However, neuroimaging studies of aggression in schizophrenia have largely focused on global or regional measures of GM volume, WM integrity, and functional connectivity (35). To our knowledge, only one study has investigated brain network organization. Graph-theoretical analysis of SCNs showed higher global efficiency and shorter characteristic path length in patients with a history of serious violence than in those without such a history and healthy controls. Average nodal degree was higher in nonviolent patients (36). Interpretation is limited by the small, all-male forensic sample and retrospective assessment of violence.

To address this gap, we investigated the association between brain network organization and aggressive behavior in a large international sample of individuals with schizophrenia spectrum disorders and healthy controls. We used graph-theoretical analysis of SCNs to examine differences in global and nodal network topology, as well as the density of large-scale brain networks, among patients with (SZ+A) and without (SZ−A) clinically significant aggression and healthy controls (HC). We expected both SZ+A and SZ−A to show altered brain network organization relative to HC, consistent with the dysconnectivity hypothesis of schizophrenia. We also expected SZ+A and SZ−A to differ in nodal topology and large-scale network density, particularly in regions and networks implicated in aggressive behavior. Given inconsistent graph-theoretical findings in schizophrenia and the paucity of studies on aggression, we did not formulate directional hypotheses.

## Methods

### Participants

This study was conducted within the Enhancing NeuroImaging Genetics through Meta-Analysis (ENIGMA) consortium’s Schizophrenia Working Group (ENIGMA-SZ), with data contributed by 26 sites across 14 countries. The pooled sample comprised 2260 individuals with schizophrenia spectrum disorders and 3058 healthy controls. Consistent with previous ENIGMA-SZ studies and for readability, the term “schizophrenia” is used throughout the manuscript. Patients were recruited from various mental healthcare settings, including psychiatric hospitals, outpatient clinics, and residential care facilities. Diagnoses were established according to Diagnostic and Statistical Manual of Mental Disorders (DSM)-III (37), DSM-IV (38), DSM-5 (39), or International Classification of Diseases (ICD)-10 criteria (40). Ethical approval was obtained from the local ethics committee at each site, and all participants provided written informed consent. Some participants were also included in two previous ENIGMA-SZ studies examining associations between aggression and regional brain structure (41,42).

### Image acquisition and processing

All sites acquired high-resolution T_1_-weighted magnetic resonance imaging scans (voxel size ≤1 mm^3^), which were processed using FreeSurfer (43) in accordance with ENIGMA-SZ protocols. Based on the Desikan–Killiany atlas (44), structural measures were extracted for 34 cortical regions (cortical thickness [CT] and surface area [SA]) and 7 subcortical regions (volume) in each hemisphere. Intracranial volume (ICV) was also extracted. Site-specific acquisition and processing details are provided in Table A1.1. Left and right hemisphere values were averaged to obtain bilateral measures. If a value for one hemisphere was unavailable, that from the contralateral hemisphere was used. Cortical volume was calculated as the product of CT and SA. Regional measures were then harmonized across sites using ComBat (45), while preserving biologically relevant variation attributable to age, sex, and study group. Finally, measures were residualized for age, sex, and ICV using linear regression models to minimize their potential confounding effects on group comparisons.

### Assessment of aggression and other clinical variables

Depending on the site, aggression was assessed using item 10 (“hostility”) of the Brief Psychiatric Rating Scale (BPRS) (46), item P7 (“hostility”) of the Positive and Negative Syndrome Scale (PANSS) (47), or item B3 (“aggressive and agitated behavior”) of the Scale for the Assessment of Positive Symptoms (SAPS) (48). The PANSS was administered at most sites (*k* = 20), followed by the SAPS (*k* = 4) and the BPRS (*k* = 2). To harmonize scores across instruments, we collapsed the highest two response categories of the BPRS and PANSS (scores 6 and 7). Because a score of 7 was rare (*n* = 4), this recoding was unlikely to materially affect the results. Scores were then recoded to a common 0–5 scale. Patients were classified as SZ+A or SZ–A based on a predefined cut-off corresponding to at least moderate aggression (harmonized score ≥ 3). This cut-off was chosen to identify clinically meaningful aggression and is consistent with those used in previous studies of other symptom domains based on the same instruments (49–51).

Positive symptoms (delusions, hallucinations, and disorganized thinking), negative symptoms (blunted affect, alogia avolition, and anhedonia/social withdrawal), poor impulse control, and lack of illness insight were assessed using corresponding items from the BPRS, PANSS, or SAPS (Table A1.2). The same harmonization procedure was applied to these symptom domains. Patients were classified as having early-course schizophrenia if they had first-episode psychosis (FEP) or a duration of illness (DOI) of ≤ 5 years. This accommodated the lack of DOI data in most FEP cohorts and was based on commonly used definitions of early-course schizophrenia (52). Patients with a DOI > 5 years were classified as having chronic schizophrenia. Definitions of FEP and DOI varied across contributing cohorts. FEP was typically defined as first presentation with psychosis, first treatment contact, first psychiatric admission, or entry into an early intervention program. DOI was calculated as the time between illness onset and assessment, with the former most commonly approximated by age at FEP or first diagnosis. Information was also available on current antipsychotic treatment status and dose. Antipsychotic doses were converted to chlorpromazine (CPZ) equivalents according to Woods et al. (53).

### Structural covariance network construction

We constructed SCNs separately for each group (SZ+A, SZ−A, and HC). Pearson correlation coefficients were calculated between all pairs of regional volumes (30), resulting in 41 × 41 association matrices. Diagonal elements and negative correlations were set to zero (54). Networks were thresholded across a range of densities (*K* = 0.10–0.25, Δ*K* = 0.01), retaining the strongest correlations at each density. Proportional thresholding facilitated comparisons by maintaining equivalent network densities across groups (55). The selected range is consistent with those used in previous ENIGMA studies of SCNs (56–59), including one in which it was selected based on prespecified criteria for connectedness, modularity, and small-world organization (56).

### Calculation of graph-theoretical metrics

Graph-theoretical metrics were summarized as the area under the curve (AUC) across the specified density range using the trapezoidal rule (60). Summarizing metrics across density thresholds reduced dependence on any single, arbitrarily selected density threshold (61).

### Global and nodal network topology

We selected a parsimonious set of graph-theoretical metrics to capture complementary aspects of network topology. For global network topology, we calculated transitivity, modularity, global efficiency, and small-worldness. These metrics quantify clustering, community segregation, network-wide integration, and the balance between segregation and integration, respectively (62). Modularity was estimated using the Louvain algorithm (63). Because the Louvain algorithm is non-deterministic and can yield different solutions across runs, it was repeated 100 times at each density threshold, with the median modularity value retained to provide a robust estimate (64). Small-worldness was calculated relative to 200 degree-preserving rewired graphs (10 × E rewiring steps) using the largest connected component (65).

For nodal network topology, we calculated degree, harmonic centrality, betweenness centrality, within-module degree (WMD), and participation coefficient (PC), which quantify nodal connectivity, proximity to other nodes, involvement in shortest paths, connectivity within communities, and connectivity across communities, respectively (62). Harmonic centrality was used instead of conventional closeness centrality, as it remains well-defined in disconnected graphs (66). To calculate WMD and PC, community partitions were first identified using Infomap at each density threshold (67). These partitions were then combined into a co-classification matrix. Each value in this matrix represented the proportion of partitions in which a pair of nodes was assigned to the same community. Values below τ = 0.50 were set to zero, and the resulting weighted matrix was repartitioned using Infomap to generate a consensus partition (68). This partition was held fixed across density thresholds, providing a consistent community reference for the calculation of WMD and PC (69).

### Large-scale brain network density

We calculated within-network density and network–rest density for six large-scale brain networks: SN, CEN, DMN, LN, RN, and ACN (see Table A1.3 for definitions). These networks were selected based on their relevance to schizophrenia and aggression, as outlined in the Introduction. Within-network density was defined as the number of connections that were present among nodes within a given network relative to the total number of possible connections among those nodes. Network–rest density was defined as the number of connections that were present between nodes in a given network and nodes outside that network, relative to the total number of possible connections between these two sets of nodes. These metrics quantify the internal connectivity of a network and its connectivity with the remainder of the brain, respectively (70).

### Statistical analyses

Differences in demographic and clinical characteristics between SZ+A and SZ−A patients were assessed using independent-samples *t*-tests, χ^2^ tests, or Mann–Whitney U tests, as appropriate. In the primary analyses, we compared graph-theoretical metrics derived from volume-based SCNs among SZ+A, SZ−A, and HC using pairwise permutation testing (10 000 permutations). For each permutation, group labels were randomly reassigned while preserving the original group sizes (71). The nominal level of statistical significance was set at 5% (two-sided). To account for multiple comparisons, false discovery rate (FDR)-corrected *p*-values were calculated using the Benjamini–Hochberg procedure (72). Within each group comparison, FDR correction was applied to predefined families of tests: all global metrics; within-network density and network–rest density, each across the six large-scale networks; and each nodal metric across the 41 brain regions. Nonparametric bootstrapping (73) (5000 resamples) was performed to estimate 95% confidence intervals (CIs) for all metrics.

We conducted two exploratory post hoc subgroup analyses. First, because illness stage differed between SZ+A and SZ−A, and GM loss is particularly pronounced during the early course of schizophrenia (74), we examined whether illness stage contributed to the group differences in global network topology. These analyses were repeated separately in patients with early-course and chronic schizophrenia, with HC included in both. Correction for multiple testing was applied as in the primary analyses. Second, patients were stratified by delusion severity (‘low’ vs ‘high’) to examine differences in SN network–rest density. This analysis was motivated by the nominally significant difference in SN network–rest density between SZ+A and HC, together with the higher delusion severity in SZ+A than in SZ−A and the hypothesized role of aberrant salience in delusion formation (75). Delusion severity was dichotomized using the same procedure as for aggression. As only SN network–rest density was examined within each group comparison, no correction for multiple testing was applied.

Prespecified sensitivity analyses evaluated (i) a cut-off corresponding to at least mild aggression (harmonized score ≥ 2), (ii) aggression defined using PANSS item P7 alone, and (iii) additional residualization of regional volumes for antipsychotic dose, expressed as chlorpromazine (CPZ) equivalents.

Analyses were performed in R (version 4.4.1), with graph-theoretical metrics calculated using the igraph package (76). The first author (JL) used ChatGPT (OpenAI) to assist with drafting, refining, and troubleshooting R code for data analyses and figure generation. Suggestions from ChatGPT were reviewed and, where applicable, tested before being incorporated. No study data were entered into ChatGPT, and all analytical decisions were made by the authors.

## Results

### Sample characteristics

Across sites, the mean age was 34 years (SD = 12) in patients and 33 years (SD = 12) in HC. The proportion of male participants was higher among patients (*n* = 1451, 64%) than among HC (*n* = 1466, 48%). Most patients were receiving antipsychotic medication (*n* = 1794, 88%), with a median CPZ-equivalent dose of 300 mg (interquartile range [IQR] = 378). Sample characteristics by site are summarized in Table A2.1.

Aggression scores were concentrated at the lower end of the harmonized scale, with similar distributions across instruments (Table A2.2). Overall, 231 patients (11%) met criteria for at least moderate aggression. SZ+A scored significantly higher than SZ−A on positive symptoms, impulsivity, poor illness insight, and several negative symptoms. They were also more likely to be in the early course of illness and had a lower median CPZ-equivalent dose (Table 1).

**Table 1.** Descriptive characteristics of SZ+A and SZ−A.

| Characteristic | SZ+A (N = 231) | SZ-A (N = 2029) | Test statistic |
| --- | --- | --- | --- |
| *Demographic characteristics* |  |  |  |
| Age, M (SD) | 33 (11) | 34 (12) | $t = -1.44, p = .152$ |
| Male, $n$ (%) | 140 (61) | 1311 (65) | $\chi^2 = 1.45, p = .229$ |
| *Positive symptoms* |  |  |  |
| Delusions, M (SD) | 4.09 (1.24) | 2.04 (1.66) | **$t = 21.48, p < .001$** |
| Hallucinations, M (SD) | 2.23 (1.99) | 1.55 (1.65) | **$t = 4.98, p < .001$** |
| Disorganized thinking, M (SD) | 1.43 (1.63) | 0.98 (1.32) | **$t = 4.10, p < .001$** |
| *Negative symptoms* |  |  |  |
| Blunted affect | 1.63 (1.56) | 1.22 (1.29) | **$t = 2.18, p = .033$** |
| Alogia | 1.25 (1.49) | 0.80 (1.16) | **$t = 2.54, p = .013$** |
| Avolition | 2.78 (1.30) | 2.03 (1.48) | $t = 1.69, p = .127$ |
| Anhedonia/social withdrawal | 2.31 (1.44) | 1.49 (1.32) | **$t = 4.72, p < .001$** |
| *Other clinical characteristics* |  |  |  |
| Impulsivity, M (SD) | 1.97 (1.34) | 0.46 (0.86) | **$t = 10.90, p < .001$** |
| Poor illness insight, M (SD) | 2.80 (1.60) | 1.46 (1.43) | **$t = 5.89, p < .001$** |
| Early-course schizophrenia, $n$ (%) | 162 (73) | 964 (52) | **$\chi^2 = 35.34, p < .001$** |
| Current antipsychotic treatment, $n$ (%) | 189 (86) | 1605 (89) | $\chi^2 = 1.52, p = .218$ |
| CPZ-equivalent dose, Mdn (IQR) | 200 (200) | 250 (400) | **$U = 117\,694, p = .001$** |

Bold values indicate statistically significant group differences ( $p < .05$ ). Abbreviations: SZ+A, schizophrenia with aggression; SZ-A, schizophrenia without aggression. M, mean; SD, standard deviation; CPZ, chlorpromazine; Mdn, median; IQR, interquartile range.

### Primary analyses

#### Global network topology

SZ+A and HC did not differ significantly on any metric of global network topology (Fig. 1; Table A3.1). Compared with both other groups, SZ−A showed lower modularity and small-worldness. Transitivity was also higher and efficiency lower in SZ−A than in HC.

**Figure 1.**
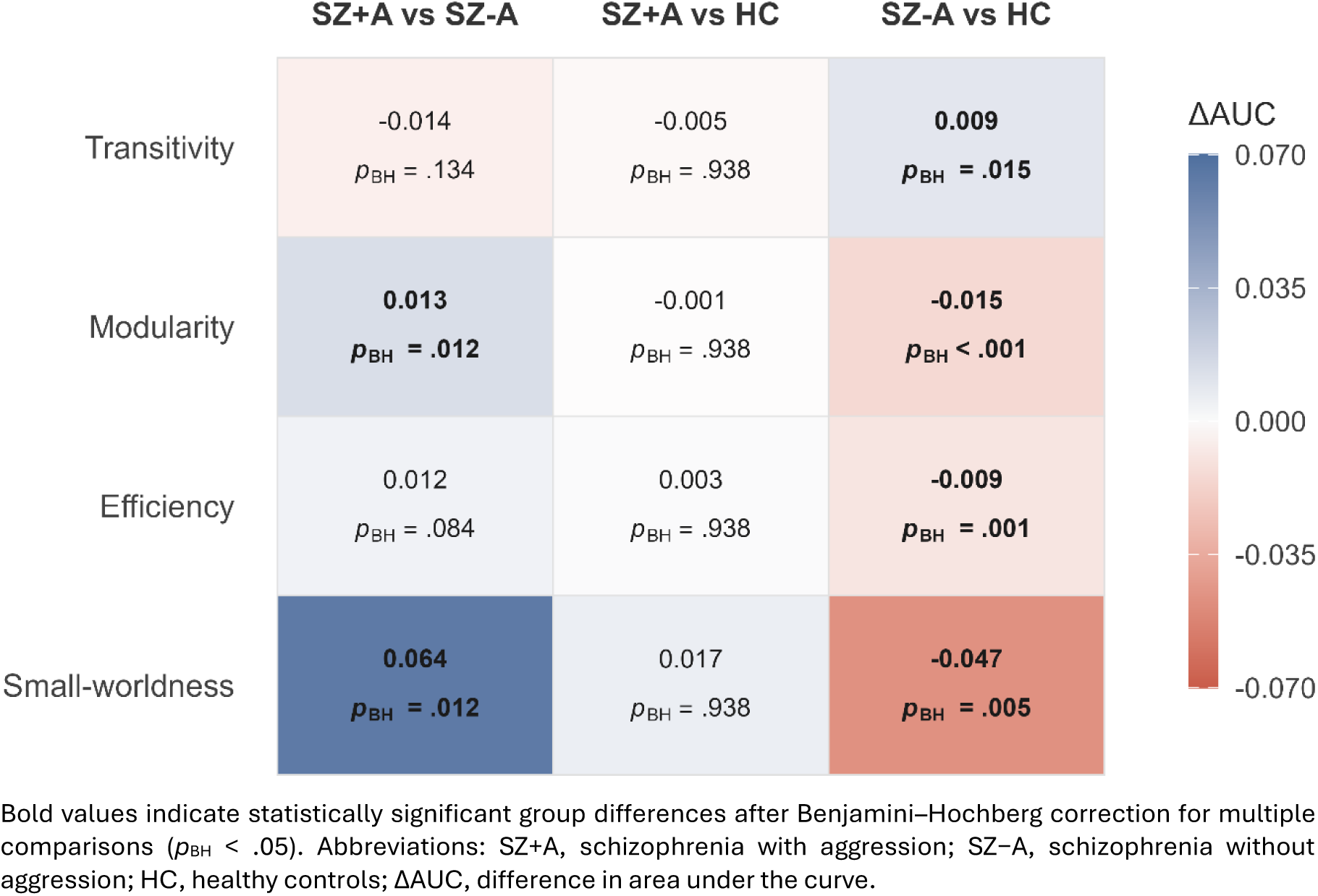
Group differences in global network topology. Bold values indicate statistically significant group differences after Benjamini–Hochberg correction for multiple comparisons (*p*_BH_ < .05). Abbreviations: SZ+A, schizophrenia with aggression; SZ−A, schizophrenia without aggression; HC, healthy controls; ΔAUC, difference in area under the curve.

#### Nodal network topology

At the nodal level, no significant differences were found between SZ+A and either SZ−A or HC. In contrast, SZ−A differed from HC on most metrics (Fig. 2; Tables A3.2–A3.6). Degree was lower in the temporal pole, entorhinal cortex, parahippocampal gyrus, lingual gyrus, cuneus, pericalcarine cortex, caudate nucleus, and nucleus accumbens. Betweenness centrality was lower in the entorhinal and pericalcarine cortices, while harmonic centrality was lower in the temporal pole, entorhinal cortex, parahippocampal gyrus, lingual gyrus, cuneus, pericalcarine cortex, and nucleus accumbens. Participation coefficient was lower in the parahippocampal gyrus and cuneus. Degree, betweenness centrality, and participation coefficient were higher in the globus pallidus.

**Figure 2.**
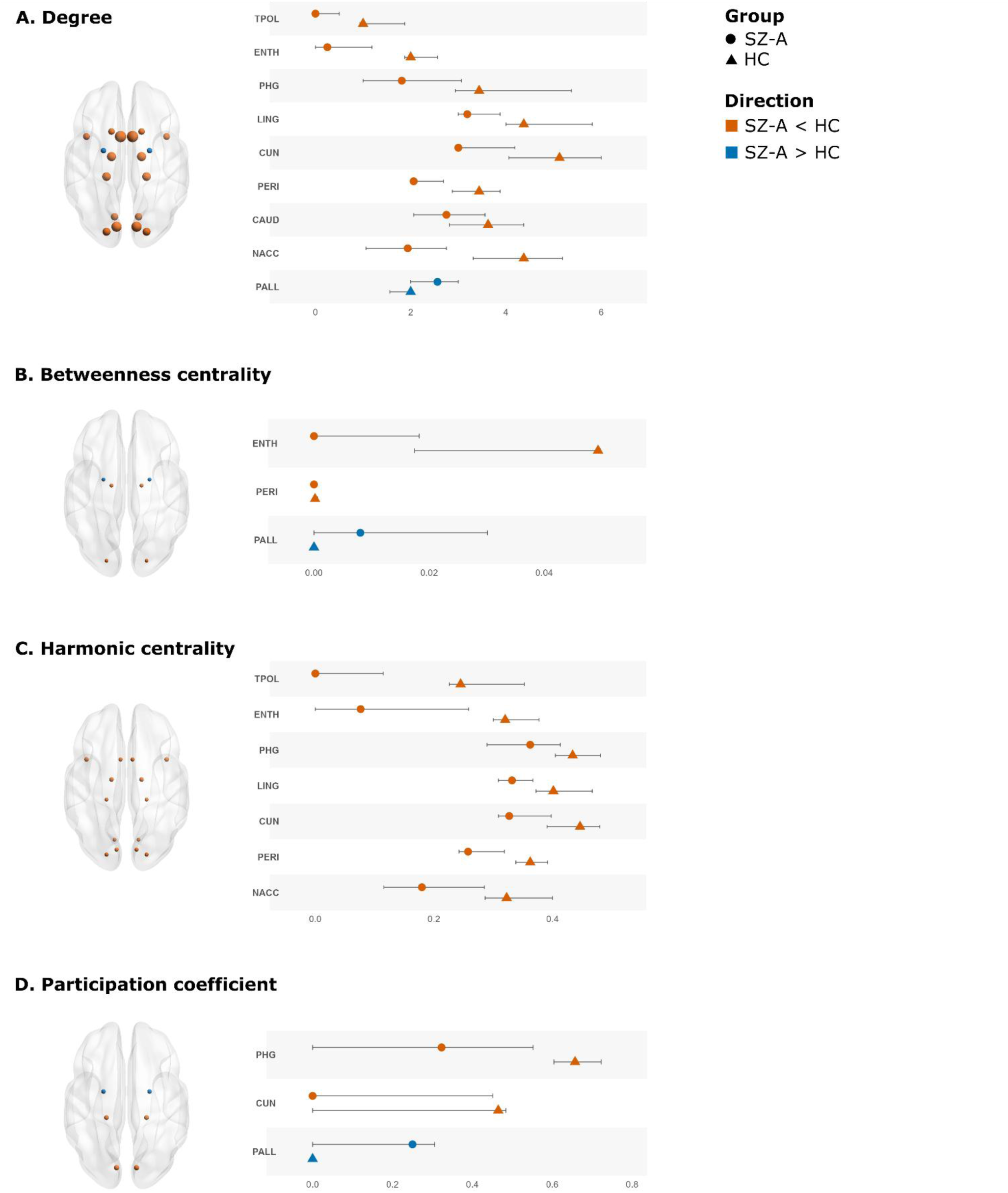
Differences in nodal network topology between SZ−A and HC. Only nodes for which group differences in area under the curve (ΔAUC) survived Benjamini–Hochberg correction for multiple comparisons (*p*_BH_ < .05) are displayed. In the brain maps, node size is proportional to ΔAUC. Symbols represent group means, and error bars represent 95% bootstrap confidence intervals. Abbreviations: SZ−A, schizophrenia without aggression; HC, healthy controls; TPOL, temporal pole; ENTH, entorhinal cortex; PHG, parahippocampal gyrus; LING, lingual gyrus; CUN, cuneus; PERI, pericalcarine cortex; CAUD, caudate nucleus; NACC, nucleus accumbens; PALL, globus pallidus.

#### Large-scale brain network density

Network–rest density of the ACN was significantly higher in SZ−A than in HC (Fig. 3; Table A3.7). At the nominal significance level, network–rest density of the SN was higher in SZ+A than in HC (*p* = .014).

**Figure 3.**
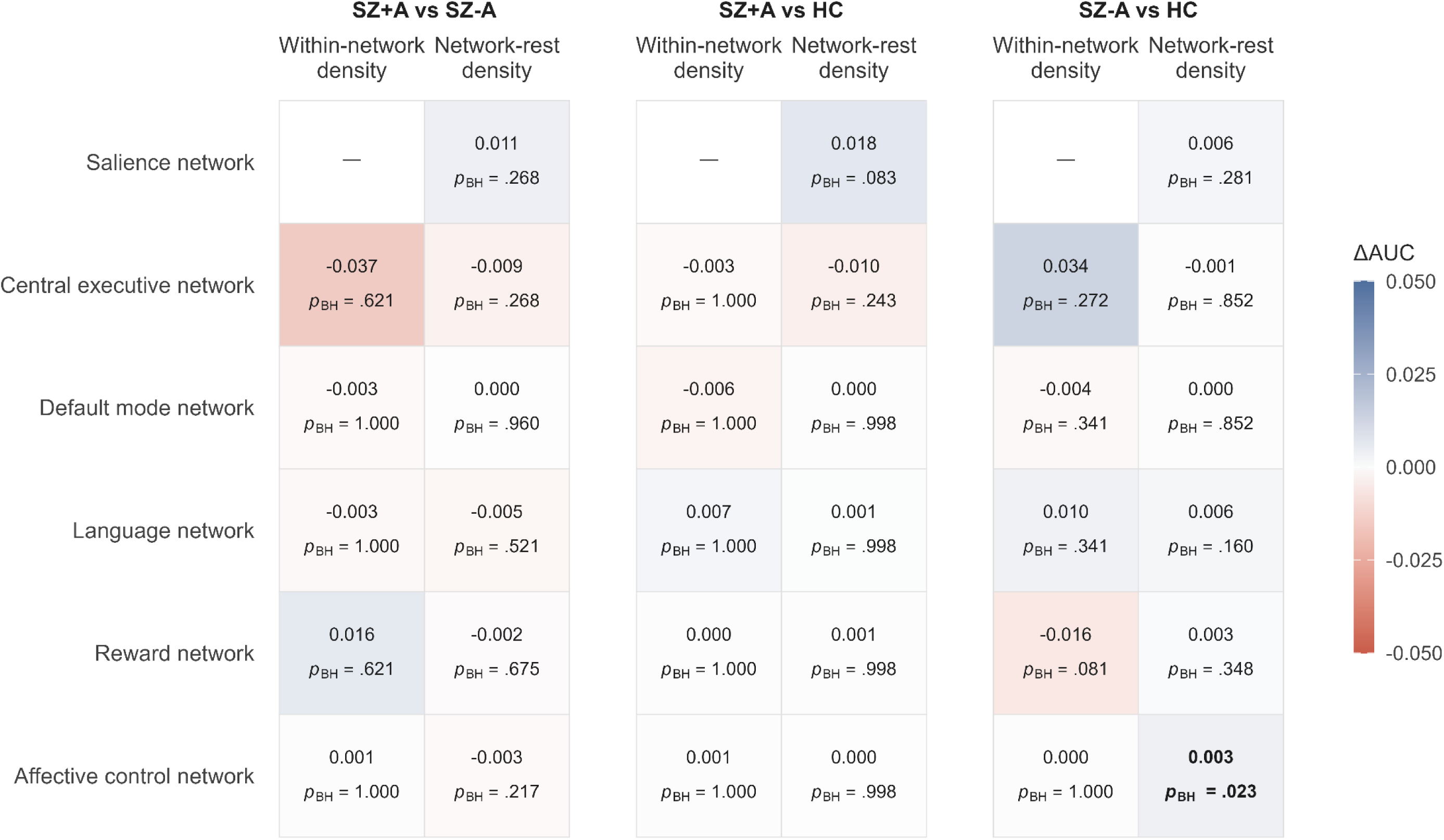
Group differences in canonical brain network density. Bold values indicate statistically significant group differences after Benjamini–Hochberg correction for multiple comparisons (*p*_BH_ < .05). Abbreviations: SZ+A, schizophrenia with aggression; SZ−A, schizophrenia without aggression; HC, healthy controls; ΔAUC, difference in area under the curve.

#### Exploratory post hoc subgroup analyses

The differences in global network topology observed for SZ−A in the primary analyses were confined to early-course patients (Table A4.1). No significant differences emerged in chronic SZ−A or between SZ+A and HC at either illness stage.

Network–rest density of the SN was higher in SZ+A with high delusion severity compared with HC (Table A4.2). No other group differences were significant.

### Sensitivity analyses

The findings were broadly similar when using a cut-off corresponding to mild aggression (Tables A5.1–7), restricting aggression to PANSS item P7 (Tables A5.8–14), and additionally residualizing regional volumes for CPZ-equivalent dose (Tables A5.15–21).

## Discussion

Overall, the findings suggest that aggression is not associated with more pronounced or widespread illness-related alterations in structural brain network organization. Despite more severe positive symptoms, greater impulsivity, and poorer illness insight, SZ+A did not significantly differ from HC in global or nodal network topology. The only indication of altered SCN organization in SZ+A was higher SN network-rest density than in HC, although this difference did not survive correction for multiple comparisons. By contrast, SZ−A showed extensive alterations in global network topology compared with SZ+A and HC, alongside selective alterations in nodal topology and large-scale network density compared with HC. These findings were robust across sensitivity analyses using alternative definitions of aggression and accounting for current antipsychotic dose.

There are several possible explanations for the limited evidence of altered SCN organization in SZ+A relative to HC. First, aggressive behavior among individuals with schizophrenia may arise through multiple pathways, including acute psychosis, impulsivity, substance misuse, and social adversity (1). The neurobiological basis of aggression may differ across these pathways, potentially obscuring group-level differences in structural covariance. Second, substance use— particularly cannabis use—may precipitate schizophrenia independently of genetic liability (77) or despite relatively preserved premorbid cognitive (78) or social functioning (79). Because substance use is also associated with aggression (80), patients following this pathway may be overrepresented in SZ+A. If they have less pronounced or distinct structural brain changes, as suggested by previous studies (81,82), SCN organization may appear preserved. This possibility could not be examined in the present study. Third, the stability of GM morphology over time (83) may mean that SCN alterations are more strongly associated with persistent than with state-dependent aggression. Our short reference period may have disproportionately captured the latter, particularly given the greater severity of concurrent positive symptoms we observed in SZ+A. Finally, aggression-related alterations in brain network organization may be more evident in structural or functional connectomes than in covariance of GM morphology. Although the relatively small SZ+A sample may have reduced statistical power, point estimates were generally similar to HC, providing little indication of substantial group differences.

The higher SN network–rest density may reflect aberrant salience processing—a mechanism implicated in the formation and maintenance of delusions (75). Consistent with this interpretation, higher SN network–rest density was observed only in SZ+A with high delusion severity in subgroup analyses. Our previous ENIGMA-SZ study similarly found that delusions partly mediated the associations of reduced global GM volume and WM integrity with aggression (41). Dysregulated salience processing could also contribute to aggression more directly through heightened attention to threatening or provocative stimuli (84) and an increased tendency to interpret ambiguous stimuli as such (85). However, the difference in SN network–rest density did not survive correction for multiple comparisons in the primary analyses, and the subgroup analyses were conducted post hoc. These findings should therefore be considered hypothesis-generating.

In SZ−A, we observed increased transitivity and reduced modularity, efficiency, and small-worldness, indicating greater local clustering, reduced global integration, and weaker community segregation. Notably, these alterations were confined to early-stage schizophrenia, possibly reflecting the rapid and unevenly distributed GM loss that occurs during the first five years after illness onset (74,86). Changes in the spatial distribution of GM loss over the course of illness may contribute to the absence of comparable alterations in chronic schizophrenia. This interpretation remains speculative and should be tested in longitudinal studies. At the nodal level, reduced centrality and participation in temporal, occipital, and subcortical regions may reflect a diminished role in information transfer and intermodular communication. The globus pallidus showed the opposite, with increased centrality and participation. This finding may partly relate to antipsychotic exposure, which has been associated with structural changes in the basal ganglia (87). Analysis of large-scale brain networks revealed increased network–rest density in the ACN. Together, these findings involved regions and large-scale networks associated with memory, visual processing, reward processing, and emotion regulation—domains commonly affected in schizophrenia (19,26,88,89).

### Strengths and limitations

To our knowledge, this is the largest study to date investigating the association between structural brain network organization and aggressive behavior in any population. Including SZ+A, SZ−A, and HC enabled us to distinguish alterations associated with aggression from those associated with schizophrenia more broadly. Additional strengths include recruitment of patients from diverse mental healthcare settings worldwide and standardized image processing protocols across sites.

Several limitations should be acknowledged. First, the study was cross-sectional, and aggression was assessed over a relatively short period. This precluded assessment of changes in SCN organization over time and distinction between transient and persistent patterns of aggressive behavior. Second, we investigated brain network organization based on regional GM volume because it captures the multidimensional nature of cortical morphology while providing a common measure across cortical and subcortical regions. However, this approach does not permit inference about the relative contributions of CT and SA. Given their distinct developmental and genetic determinants (90), analyses of these phenotypes may provide complementary insights. Third, aggression was assessed with single items from instruments developed to assess symptom dimensions of schizophrenia. Multi-item instruments specifically developed to assess aggression would likely provide greater sensitivity. Fourth, constructing SCNs at the group level required dichotomization of aggressive behavior, potentially reducing sensitivity to variation in severity. Group-level estimation of graph-theoretical metrics also required permutation testing of group differences, which precluded covariate adjustment for potential confounders.

Nevertheless, important sources of methodological and biological variation were reduced before network construction by harmonizing regional volumes across sites and residualizing them for age, sex, and intracranial volume. Individual-level SCN methods have recently been developed, allowing dimensional modeling. However, these approaches construct SCNs from perturbations of a normative covariance network (91) or from joint variation in regional deviations from normative morphometry (56,57). As such, they address different research questions. Fifth, although sensitivity analyses accounted for current antipsychotic dose, the influence of cumulative antipsychotic exposure—which may affect brain structure (92)—and confounding by indication cannot be fully ruled out. Finally, we lacked information on social risk factors, substance misuse, and specific cognitive deficits, which may confound or mediate associations between brain structure and aggression. Nevertheless, comparisons between SZ+A and SZ−A across a wide range of clinical characteristics and exploratory subgroup analyses provided insight into potential mechanisms.

### Clinical implications and recommendations for future research

Our findings provide little evidence that group-level SCNs derived from regional GM volume can serve as markers of aggressive behavior in individuals with schizophrenia. This is in line with the wider neuroimaging literature on aggression, in which associations with brain structure have been inconsistent (7). A better understanding of aggressive behavior will likely require the integration of neurobiological, psychological, and social risk factors (1). If replicated, the finding of increased SN network–rest density in SZ+A could motivate investigation of aberrant salience processing as a mechanism contributing to aggressive behavior in schizophrenia, particularly in the presence of severe delusions. More broadly, our findings suggest that associations between brain structure and aggressive behavior may vary according to the severity of specific symptoms. Such heterogeneity may help explain the inconsistent findings reported across neuroimaging studies of aggression in schizophrenia (35) and should be investigated further. Future studies should investigate associations between brain network alterations and recurrent aggressive behavior, ideally using longitudinal designs.

## Data Availability

The data used in this study are not publicly available and cannot be shared directly by the authors, as they are subject to the data-sharing requirements of the contributing sites. Requests may be submitted through ENIGMA-SZ, subject to site approval.

## Acknowledgements

The ENIGMA project is supported by the National Institute of Biomedical Imaging and Bioengineering of the National Institutes of Health (NIH) under grant number U54EB020403. Funding information for the participating sites can be found in Appendix 1. JL is supported by a Veni grant from the Dutch Research Council (grant VI.Veni.241R.005). AR is supported by the Richard Perry University Professorship and TN-J by the German Center for Mental Health (DZPG) and the Deutsche Forschungsgemeinschaft (Sonderforschungsbereich 1436). The funders had no role in study design, data collection or analysis, decision to publish or the preparation of the manuscript.

The data used in this study are not publicly available and cannot be shared directly by the authors, as they are subject to the data-sharing requirements of the participating sites. Requests may be submitted through ENIGMA-SZ, subject to site approval.

We also acknowledge the late Professor Dan J. Stein for his longstanding collaboration within ENIGMA and his contributions to psychiatry, neuroscience, and mental health research. His intellectual generosity, mentorship, and support have had a lasting impact on colleagues across the consortium and the field.

During the preparation of this manuscript, the first author (JL) used ChatGPT (OpenAI) to assist with identifying potentially relevant literature, language editing, and improving clarity. All literature suggested by ChatGPT was independently verified and assessed for relevance. The authors reviewed and edited the manuscript as needed and take full responsibility for its content.

## Disclosures

The authors report no conflicts of interest.

## Notes

### Competing Interest Statement

The authors have declared no competing interest.

### Author Declarations

The Ethics Committee or Institutional Review Board at each of the 26 participating sites gave ethical approval for the collection and use of the data included in this work. All participants provided written informed consent.

