## Appendices for "Structural brain network organization and aggressive behavior in schizophrenia: a mega-analysis of 2260 patients and 3058 healthy controls from the ENIGMA-Schizophrenia Working Group"

**Appendix 6: References**

#### Appendix 1
Study and methodological details

#### Funding information for participating sites

##### CIAM

The CIAM study was supported by the South African Medical Research Council and the National Research Foundation of South Africa. FS is supported by the South African Medical Research Council through its Division of Research Capacity Development under the Early Investigators Program.

##### CLiNG

This work was supported by the German Research Foundation (grant GR1950/5-1 to OG).

##### COBRE

The COBRE study was supported by the National Institutes of Health (grants R01EB006841 and P20GM103472) and the National Science Foundation (grant 1539067). JAT and VDC are supported by the National Institutes of Health (grants 5R01MH094524 and 5R01MH121246).

##### ESO

The ESO study was supported by the Ministry of Health of the Czech Republic (grants NU21-08-00432 and NU22-04-00143).

##### FIDMAG

The FIDMAG study was supported by a Miguel Servet Research Contract (grant MS14/00041), the Plan Nacional de I+D+i 2013–2016 (grant PI14/00292), a Juan de la Cierva-Formación contract (grant FJCI-2015-25278), the Instituto de Salud Carlos III-Subdirección General de Evaluación y Fomento de la Investigación, the European Regional Development Fund, and the Centro de Investigación Biomédica en Red de Salud Mental.

##### FOR2107

This work was supported by grants from the German Research Foundation (DFG) awarded to TK through FOR2107 (KI 588/14-1 and KI 588/14-2) and SFB/TRR 393 (Trajectories of Affective Disorders; grants 521379614 and KI 588/23-1), to UD (DA 1151/5-1, DA 1151/5-2, DA 1151/9-1, DA 1151/10-1, DA 1151/11-1, and DA 1151/21-1), and to IN (NE 2254/1-2, NE 2254/2-1, NE 2254/3-1, and NE 2254/4-1). Additional funding was provided by Germany's Excellence Strategy (EXC 3066/1 The Adaptive Mind; grant 533717223), the DYNAMIC Center, funded by the LOEWE program of the Hessian Ministry of Science and Arts (grant LOEWE1/16/519/03/09.001(0009)/98), and the Wellcome Trust-funded DIALOG consortium (Understanding Disorganisation: A Language-Focused Global Initiative in Psychosis; grant 314138/Z/24/Z). UD was also supported by the Interdisciplinary Centre for Clinical Research of the University of Münster (grant Dan3/016/26).

##### GAP

The GAP study was supported by the NIHR Biomedical Research Centre at South London and Maudsley NHS Foundation Trust and King's College London.

##### IGP

The IGP study was supported by the Australian National Health and Medical Research Council (grants APP630471 and APP1081603), the Schizophrenia Research Institute Grant-in-Aid program, and the Australian Research Council Centre of Excellence in Cognition and its Disorders (grant CE110001021). MJG was supported by an Australian National Health and Medical Research Council R.D. Wright Biomedical Career Development Fellowship (grant

APP1061875). Recruitment through the Australian Schizophrenia Research Bank was supported by an Australian National Health and Medical Research Council Enabling Grant (grant 386500), the Pratt Foundation, Ramsay Health Care, the Viertel Charitable Foundation, and the Schizophrenia Research Institute.

**IMH**
This work was supported by grants from the National Healthcare Group (SIG/05004 and SIG/05028) and the Singapore Bioimaging Consortium (RP C009/2006), awarded to KS.

**JBNU**
The JBNU study was supported by the Korea Health Technology R&D Project through the Korea Health Industry Development Institute (KHIDI), funded by the Ministry of Health & Welfare (grant HR18C0016), and the National Research Foundation of Korea (NRF), funded by the Korean government (grants RS-2024-00335201 and RS-2025-16071835).

**KaSP**
The KaSP study was supported by the Swedish Research Council (grants 2024-02812, 2023-02827, 2017-00875, and 523-2014-3467), the Swedish Brain Foundation (grant FO2023-0333), Stockholm County Council (grants 20160328, 20180487, 20190175, and 20190447), the Torsten Söderberg Foundation, the Swedish Medical Society, the Erling Persson Foundation, and doctoral funding from Karolinska Institutet.

**MCIC**
The MCIC study was supported by the National Institutes of Health (grants P41RR14075 and R01EB005846 to VDC), the Department of Energy (grant DE-FG02-99ER62764), the Mind Research Network, the Morphometry (grant 1U24RR021382A) and Function (grants U24RR021992-01, MO1RR025758-01, and 1RC1MH089257 to VDC) testbeds of the Biomedical Informatics Research Network, the German Research Foundation (research fellowship to SE), the National Alliance for Research on Schizophrenia & Depression (Young Investigator Award to SE), and the Else Kröner-Fresenius Foundation (grant 2019-A118 to DA).

**MPRC**
The MPRC study was supported by the National Institutes of Health (grants U01MH108148, 2R01EB015611, R01MH112180, R01DA027680, R01MH085646, P50MH103222, and T32MH067533), a State of Maryland contract (M00B6400091), and the National Science Foundation (grant 1620457).

**NU**
The NU study was supported by the National Institutes of Health (grants P50MH071616, R01EB020062, R01MH056584, R01MH084803, and U01MH097435 to LW, and grant T32NS047987 to DC), and the National Science Foundation (grants 1636893 and 1734853 to LW).

**Olin**
The Olin study was supported by the National Institutes of Health (grant R01MH077945).

**PAFIP**
The PAFIP study was supported by the Instituto de Salud Carlos III (grant FIS PI14/00639), MINECO (grant SAF2013-46292-R), and the PSYSCAN project (European Commission grant agreement no. 603196). RRG and PS were supported by the Plan de Generación de Conocimiento (grant PID2021-122853OA-I00) and the Plan de Consolidación (grant CNS2023-143647), both

from the Agencia Estatal de Investigación, and by ERANET Neuron JTC 2023 (grant ERP-2023-23684211).

**RomeSL**
FP was supported by the Italian Ministry of Health (Ricerca Corrente 2023).

**RSCZ**
The RSCZ study was supported by the Russian Foundation for Basic Research (grant 20-013-00748).

**SCORE**
The SCORE study was supported by the Swiss National Science Foundation (grant 3232B0_119382). The FePsy Study Group from the University of Basel contributed to participant recruitment and was supported by the Swiss National Science Foundation (grant 3200-057216/1 and extensions 2 and 3).

**SNUH**
This work was supported by the National Research Foundation of Korea and Korea Brain Research Institute (grants 2019R1C1C1002457, 2020M3E5D9079910, RS-2023-00266120, and 21-BR-03-01).

**SWIFT**
The SWIFT study was supported by the Brain & Behavior Research Foundation (grant 28445) and the Novartis Foundation (grant 20A058).

**UCISZ**
The UCISZ study was supported by the National Institute of Mental Health (grant R21MH097196 to TvE). Data were processed with the high-performance computing cluster at the University of California, Irvine, supported by Joseph Farran, Harry Mangalam, Adam Brenner, the National Center for Research Resources, and the National Center for Advancing Translational Sciences (grant UL1TR000153).

**UMCU**
This work was supported by the Geestkracht program of the Netherlands Organization for Health Research and Development (grant 10-000-1001), the University Medical Centre Utrecht, participating pharmaceutical companies (Lundbeck, AstraZeneca, Eli Lilly, and Janssen Cilag), and participating mental healthcare institutions (Altrecht, GGZ Centraal, and Delta).

**UNINA**
FI, AdB, and AB were supported by NextGenerationEU, the Italian Ministry of University and Research, the National Recovery and Resilience Plan, and MNESYS – A Multiscale Integrated Approach to the Study of the Nervous System in Health and Disease (project PE0000006; Decreto n. 1553 11.10.2022).

**VOICES**
The VOICES study was supported by the National Health and Medical Research (NHMRC) Council (grant GNT1060664). SLR was supported by an NHMRC Senior Research Fellowship (grant GNT1154651). TEVR was supported by an Al and Val Rosenstrauss Fellowship from the Rebecca L. Cooper Medical Research Foundation and an NHMRC Early Career Fellowship. MH and WW were supported by the National Imaging Facility (NIF), and SC and PS by Australian Government Research Training Program Scholarships. The authors also acknowledge the

facilities and scientific assistance of the NIF, a National Collaborative Research Infrastructure Strategy capability, at the Swinburne Neuroimaging Facility, Swinburne University of Technology.

**Zürich**

The Zürich study was supported by the Swiss National Science Foundation (SNSF). MK is supported by the SNSF (grant 32003B_219240).

**Table A1.1.** Scanner, acquisition, and processing details by site.

| Site | Scanner model | Field strength | Sequence | Parameters | Plane | FreeSurfer version | Operating system |
|---|---|---|---|---|---|---|---|
| CIAM | Siemens MAGNETOM Allegra | 3T | MPRAGE | TE: 1.53/3.21/4.89/6.57 ms<br>TR: 2.53 s<br>Flip angle: 7°<br>Voxel size: 1.3 × 1 × 1.3 mm<br>FOV: 256 × 256 mm<br>Number of slices: 128 | Sagittal | 5.3.0 | Linux |
| CLiNG | Siemens MAGNETOM TrioTim | 3T | MPRAGE | TE: 3.26 ms<br>TR: 2.25 s<br>TI: 0.9 s<br>Flip angle: 9°<br>Voxel size: 1 × 1 × 1 mm<br>Matrix size: 256 × 256<br>Number of slices: 192 | Sagittal | 5.3.0 | Ubuntu |
| COBRE | Siemens MAGNETOM Trio | 3T | MPRAGE | TE: 1.64/3.5/5.36/7.22/9.08 ms<br>TR: 2.53 s<br>TI: 1.2 s<br>Flip angle: 7°<br>Voxel size: 1 × 1 × 1 mm<br>Matrix size: 256 × 256 | Sagittal | 5.3.0 | RHEL |
| ESO | Siemens MAGNETOM Trio | 3T | MPRAGE | TE: 4.63 ms<br>TR: 2.3 s<br>TI: 0.9 s<br>Voxel size: 1 × 1 × 1 mm<br>Matrix size: 256 × 256 | Sagittal | 5.3.0 | Linux |

| | | | | | | | |
|---|---|---|---|---|---|---|---|
| FIDMAG | GE Signa | 1.5T | MPRAGE | TE: 4 ms<br>TR: 2 s<br>Flip angle: 15°<br>Voxel size: 0.5 × 0.5 × 1 mm<br>Matrix size: 512 × 512<br>Number of slices: 180 | Axial | 5.3.0 | Ubuntu |
| FOR2107 | Siemens MAGNETOM TrioTim | 3T | MPRAGE | TE: 2.26 ms<br>TR: 1.9 s<br>TI: 0.9 s<br>Flip angle: 9°<br>Voxel size: 1 × 1 × 1 mm<br>Matrix size: 256 × 256<br>Number of slices: 176 | Sagittal | 5.3.0 | RHEL |
| | Siemens MAGNETOM Prisma | 3T | MPRAGE | TE: 2.28 ms<br>TR: 2.13 s<br>TI: 0.9 s<br>Flip angle: 8°<br>Voxel size: 1 × 1 × 1 mm<br>Matrix size: 256 × 256<br>Number of slices: 192 | Sagittal | 5.3.0 | RHEL |
| GAP | GE Signa HDx | 3T | BRAVO | TE: 2.85 ms<br>TR: 6.99 s<br>TI: 0.65 s<br>Flip angle: 8°<br>Voxel size: 1.02 × 1.02 × 1.2 mm<br>Matrix size: 256 × 256<br>Number of slices: 166 | Sagittal | 5.3.0 | Linux |

| | | | | | | | |
|---|---|---|---|---|---|---|---|
| IGP | Philips Achieva TX | 3T | TFE | TE: 4.1 ms<br>TR: 8.9 ms<br>Flip angle: 8°<br>Voxel size: 0.83 × 0.83 × 0.9 mm<br>Matrix size: 288 × 288<br>Number of slices: 200 | Sagittal | 5.3.0 | Mac OS X |
| IMH | Philips Achieva | 3T | TFE | TE: 3.3 ms<br>TR: 7.2 s<br>Flip angle: 8°<br>Voxel size: 0.9 × 0.9 × 0.9 mm<br>Matrix size: 256 × 204<br>Number of slices: 180 | Axial | 5.3.0 | Mac OS X |
| JBNU | Siemens MAGNETOM Verio | 3T | MPRAGE | TE: 2.45 ms<br>TR: 1.9 s<br>Flip angle: 9°<br>Voxel size: 1 × 1 × 1 mm<br>FOV: 250 × 250 mm | Sagittal | 7.1.0 | Linux |
| KaSP | GE | 3T | BRAVO | TE: 3.06 ms<br>TR: 7.90 ms<br>TI: 0.45 s<br>Flip angle: 12°<br>Voxel size: 0.93 × 0.93 × 1.2 mm<br>Matrix size: 256 × 256<br>Number of slices: 146 | Sagittal | 5.3.0 | Linux |
| MCIC | Siemens | 1.5T | FLASH | TE: 4.76 ms<br>TR: 12 ms<br>Flip angle: 20°<br>Voxel size: 0.63 × 0.63 × 1.5 mm<br>Matrix size: 256 × 256<br>Number of slices: 128 | Coronal | 4.0.1 | Linux |

| | | | | | | | |
|---|---|---|---|---|---|---|---|
| | GE | 3T | BRAVO | TE: 3.79 ms<br>TR: 2.53 s<br>TI: 1.1 s<br>Flip angle: 7°<br>Voxel size: 0.63 × 0.63 × 1.5 mm<br>Matrix size: 256 × 256<br>Number of slices: 128 | Coronal | 4.0.1 | Linux |
| MPRC | Siemens MAGNETOM Allegra | 3T | MPRAGE | TE: 4.3 ms<br>TR: 2.5 s<br>TI: 1 s<br>Flip angle: 8°<br>Voxel size: 1 × 1 × 1 mm | Sagittal | 5.3.0 | Linux |
| | Siemens MAGNETOM Trio | 3T | MPRAGE | TE: 2.9 ms<br>TR: 2.3 s<br>TI: 0.9 s<br>Flip angle: 9°<br>Voxel size: 1 × 1 × 1 mm | Sagittal | 5.3.0 | Linux |
| NU | Siemens MAGNETOM Vision | 1.5T | turbo-FLASH | TE: 5.4 ms<br>TR: 20 ms<br>Flip angle: 30°<br>Voxel size: 1 × 1 × 1 mm<br>Matrix size: 256 × 256<br>Number of slices: 180 | Axial | 5.3.0 | CentOS |
| | | | MPRAGE | TE: 4 ms<br>TR: 9.7 ms<br>Flip angle: 10°<br>Voxel size: 1 × 1 × 1.25 mm<br>Matrix size: 256 × 256<br>Number of slices: 128 | Axial | 5.3.0 | CentOS |

| | | | | | | | |
|---|---|---|---|---|---|---|---|
| Olin | Siemens MAGNETOM Allegra | 3T | MPRAGE | TE: 4.13 ms<br>TR: 2.2 s<br>TI: 0.77 s<br>Flip angle: 13°<br>Voxel size: 0.8 × 0.8 × 0.8 mm<br>Matrix size: 240 × 320<br>Number of slices: 208 | Axial | 5.1.0 | CentOS |
| PAFIP | GE Signa | 1.5T | SPGR | TE: 5 ms<br>TR: 24 ms<br>Flip angle: 45°<br>Voxel size: 1.02 × 1.02 × 1.5 mm<br>Matrix size: 256 × 192 | Coronal | 6.0.0 | Ubuntu |
| | Philips Achieva | 3T | TFE | TE: 3.7 ms<br>TR: 8.2 ms<br>Flip angle: 8°<br>Voxel size: 0.94 × 0.94 × 1 mm<br>Matrix size: 256 × 256<br>Number of slices: 160 | Sagittal | 6.0.0 | Ubuntu |
| RomeSL | Siemens MAGNETOM Allegra | 3T | MDEFT | TE: 2.4 ms<br>TR: 7.92 ms<br>TI: 0.91 s<br>Flip angle: 15°<br>Voxel size: 1 × 1 × 1 mm<br>Matrix size: 224 × 256 | Sagittal | 6.0dev | Linux |
| RSCZ | Philips Achieva | 3T | TFE | TE: 3.7 ms<br>TR: 8.2 s<br>TI: 1.02 s<br>Flip angle: 8°<br>Voxel size: 0.83 × 0.83 × 1 mm<br>FOV: 240 × 240 mm | Sagittal | 5.3.0 | CentOS |

| | | | | | | | |
|---|---|---|---|---|---|---|---|
| SCORE | Philips Achieva | 3T | MPRAGE | TE: 3.4 ms<br>TR: 2 s<br>TI: 1 s<br>Flip angle: 8°<br>Voxel size: 1 × 1 × 1 mm<br>Matrix size: 256 × 256 | Sagittal | 7.4.1 | macOS |
| SNUH | Siemens MAGNETOM Trio | 3T | MPRAGE | TE: 1.89 ms<br>TR: 1.67 s<br>Flip angle: 9°<br>Voxel size: 1 × 1 × 1 mm<br>Matrix size: 250 × 250<br>Number of slices: 208 | Sagittal | 5.3.0 | Mac OS X |
| SWIFT | Siemens MAGNETOM Verio | 3T | MPRAGE | TE: 2.98 ms<br>TR: 2.3 s<br>TI: 0.9 s<br>Voxel size: 1 × 1 × 1 mm<br>Matrix size: 256 × 256<br>Number of slices: 160 | Sagittal | 7.1.1 | Linux |
| UCISZ | Philips Achieva | 3T | TFE | TE: 4.56 ms<br>TR: 11 ms<br>Flip angle: 18°<br>Voxel size: 0.75 × 0.75 × 0.75 mm<br>Matrix size: 320 × 274<br>Number of slices: 200 | Sagittal | 6.0dev | CentOS |
| UMCU | Philips Intera/Achieva | 1.5T | FFE | TE: 4.6 ms<br>TR: 30 ms<br>Flip angle: 30°<br>Voxel size: 1 × 1 × 1.2 mm<br>Matrix size: 256 × 256<br>Number of slices: 160–180 | Coronal | 5.1.0 | CentOS |

| | | | | | | | |
|---|---|---|---|---|---|---|---|
| UNINA | Siemens MAGNETOM TrioTim | 3T | MPRAGE | TE: 3.4 ms<br>TR: 1.9 s<br>TI: 0.9 s<br>Flip angle: 9°<br>Voxel size: 1 × 1 × 1 mm<br>Number of slices: 160 | Axial | 6.0.1 | Ubuntu |
| VOICES | Siemens MAGNETOM TrioTim | 3T | MPRAGE | TE: 2.52 ms<br>TR: 1.9 s<br>Voxel size: 1 × 1 × 1 mm<br>Matrix size: 256 × 256<br>Number of slices: 176 | Sagittal | 6.0.0 | macOS |
| Zürich | Philips | 3T | TFE | TE: 3.8 ms<br>TR: 8.4 ms<br>Flip angle: 8°<br>Voxel size: 1 × 1 × 1 mm<br>Matrix size: 240 × 240<br>Number of slices: 160 | Sagittal | 6.0.0 | Linux |

Abbreviations: MPRAGE, Magnetization-Prepared Rapid Gradient Echo; TE, echo time; TR, repetition time; FOV, field of view; TI, inversion time; RHEL, Red Hat Enterprise Linux; GE, General Electric; BRAVO, Brain Volume Imaging; FLASH, Fast Low-Angle Shot; MDEFT, Modified Driven Equilibrium Fourier Transform; SPGR, spoiled gradient recalled echo; TFE, Turbo Field Echo; FFE, Fast Field Echo.

**Table A1.2.** Items used to assess aggressive behavior and other clinical variables.

| Variable | Item |
|---|---|
| Aggressive behavior | BPRS item 10 ‘Hostility’<br>PANSS item P7 ‘Hostility’<br>SAPS item B3 ‘Aggressive and agitated behavior’ |
| Delusions | PANSS item P1 ‘Delusions’<br>SAPS item D13 ‘Global rating of severity of delusions’ |
| Hallucinations | BPRS item 12 ‘Hallucinatory behavior’<br>PANSS item P3 ‘Hallucinatory behavior’<br>SAPS item H7 ‘Global rating of severity of hallucinations’ |
| Disorganized thinking | BPRS item 4 ‘Conceptual disorganization’<br>PANSS item P2 ‘Conceptual disorganization’<br>SAPS item P9 ‘Global rating of positive formal thought disorder’ |
| Negative symptoms | Mean score on the negative symptoms subscale of the BPRS, PANSS, or SAPS |
| Poor impulse control | PANSS item P14 ‘Poor impulse control’ |
| Lack of illness insight | PANSS item G12 ‘Lack of judgment and insight’ |

Abbreviations: BPRS, Brief Psychiatric Rating Scale; PANSS, Positive and Negative Syndrome Scale; SAPS, Scale for the Assessment of Positive Symptoms.

**Table A1.3.** Definitions of large-scale brain networks used in the present study.

| Network | Constituent regions |
|---|---|
| Salience network (8) | INS, CACC |
| Central executive network (8) | RMFG, CMFG, IPL, SPL |
| Default mode network (93) | PCC, ISTH, PREC, MOFC, IPL, BANKS, MTG, TPOL, PHG, ENTH, HIPPO, NACC THAL |
| Language network (94) | POP, PTRI, STG, MTG, IPL |
| Reward network (95,96) | MOFC, LOFC, RACC, AMYG, NACC CAUD,PUT, THAL |
| Affective control network (10–12) | MOFC, LOFC, POP, PTRI, PORB, RMFG CMFG, SFG, RACC, CACC, INS, IPL, AMYG, NACC, THAL |

Networks were selected based on their relevance to schizophrenia and aggression, as outlined in the Introduction, and operationalized using regions from the Desikan–Killiany atlas (44). Abbreviations: INS, insular cortex; CACC, caudal anterior cingulate cortex; RMFG, rostral middle frontal gyrus; CMFG, caudal middle frontal gyrus; IPL, inferior parietal lobule; SPL, superior parietal lobule; PCC, posterior cingulate cortex; ISTH, isthmus of the cingulate cortex; PREC, precuneus; MOFC, medial orbitofrontal cortex; IPL, inferior parietal lobule; BANKS, banks of the superior temporal sulcus; MTG, middle temporal gyrus; TPOL, temporal pole; PHG, parahippocampal gyrus; ENTH, entorhinal cortex; HIPPO, hippocampus; NACC, nucleus accumbens; THAL, thalamus; POP, pars opercularis; PTRI, pars triangularis; STG, superior temporal gyrus; MTG, middle temporal gyrus; IPL, inferior parietal lobule; MOFC, medial orbitofrontal cortex; LOFC, lateral orbitofrontal cortex; RACC, rostral anterior cingulate cortex; AMYG, amygdala; NACC, nucleus accumbens; CAUD, caudate nucleus; PUT, putamen; THAL, thalamus; MOFC, medial orbitofrontal cortex; LOFC, lateral orbitofrontal cortex; POP, pars opercularis; PTRI, pars triangularis; PORB, pars orbitalis; RMFG, rostral middle frontal gyrus; CMFG, caudal middle frontal gyrus; SFG, superior frontal gyrus; RACC, rostral anterior cingulate cortex; CACC, caudal anterior cingulate cortex; INS, insular cortex; IPL, inferior parietal lobule; AMYG, amygdala; NACC, nucleus accumbens; THAL, thalamus.

#### Appendix 2
Sample characteristics

**Table A2.1.** Descriptive characteristics of participants by site.

| Site | Country | SZ ($N$ = 2260) | | | HC ($N$ = 3058) | | |
|---|---|---|---|---|---|---|---|
| | | $n$ | Male, $n$ (%) | Age, M (SD) | $n$ | Male, $n$ (%) | Age, M (SD) |
| CIAM | South Africa | 18 | 11 (61) | 31 (7) | 31 | 17 (55) | 27 (5) |
| CLiNG | Germany | 48 | 36 (75) | 33 (9) | 322 | 132 (41) | 25 (5) |
| COBRE | USA | 62 | 51 (82) | 37 (13) | 63 | 44 (70) | 37 (12) |
| ESO | Czechia | 26 | 12 (46) | 30 (7) | 24 | 11 (46) | 29 (7) |
| FIDMAG | Spain | 124 | 95 (77) | 38 (11) | 123 | 54 (44) | 38 (10) |
| FOR2107 | Germany | 93 | 45 (48) | 38 (12) | 733 | 250 (34) | 34 (13) |
| GAP | UK | 53 | 10 (19) | 27 (6) | 94 | 57 (61) | 26 (7) |
| IGP | Australia | 30 | 20 (67) | 43 (9) | 39 | 24 (62) | 36 (11) |
| IMH | Singapore | 160 | 108 (68) | 34 (9) | 102 | 59 (58) | 33 (10) |
| JBNU | South Korea | 370 | 184 (50) | 38 (14) | 354 | 171 (48) | 41 (14) |
| KaSP | Sweden | 81 | 50 (62) | 29 (7) | 56 | 28 (50) | 27 (5) |
| MCIC | USA | 107 | 81 (76) | 34 (11) | 92 | 63 (68) | 33 (12) |
| MPRC | USA | 104 | 79 (76) | 34 (12) | 175 | 76 (43) | 37 (14) |
| NU | USA | 105 | 72 (69) | 34 (13) | 77 | 43 (56) | 31 (14) |
| Olin | USA | 45 | 34 (76) | 29 (10) | 54 | 20 (37) | 35 (12) |
| PAFIP | Spain | 214 | 123 (57) | 30 (9) | 194 | 120 (62) | 29 (8) |
| RomeSL | Italy | 130 | 84 (65) | 39 (11) | 101 | 63 (62) | 37 (11) |
| RSCZ | Russia | 36 | 36 (100) | 22 (3) | 37 | 37 (100) | 22 (3) |
| SCORE | Germany | 63 | 45 (71) | 27 (7) | 44 | 17 (39) | 26 (4) |
| SNUH | South Korea | 39 | 18 (46) | 23 (6) | 40 | 20 (50) | 23 (4) |
| SWIFT | Switzerland | 23 | 16 (70) | 35 (11) | 13 | 5 (38) | 29 (4) |
| UCISZ | USA | 27 | 22 (81) | 43 (11) | 30 | 23 (77) | 41 (12) |
| UMCU | Netherlands | 167 | 134 (80) | 27 (6) | 138 | 67 (49) | 27 (9) |
| UNINA | Italy | 45 | 31 (69) | 38 (10) | 54 | 29 (54) | 43 (15) |
| VOICES | Australia | 30 | 9 (30) | 42 (10) | 40 | 18 (45) | 32 (12) |
| Zürich | Switzerland | 60 | 45 (75) | 31 (8) | 28 | 18 (64) | 33 (9) |

Abbreviations: SZ, schizophrenia; HC, healthy controls; M, mean; SD, standard deviation.

**Table A2.2.** Distribution of aggression scores in individuals with schizophrenia.

| Score | BPRS item 10 | PANSS item P7 | SAPS item B3 | Total |
|---|---|---|---|---|
| 0 'None' | 88 (53) | 1111 (71) | 318 (61) | 1517 (67) |
| 1 'Minimal' | 28 (17) | 232 (15) | 54 (10) | 314 (14) |
| 2 'Mild' | 20 (12) | 134 (9) | 44 (8) | 198 (9) |
| 3 'Moderate' | 4 (2) | 72 (5) | 19 (4) | 95 (4) |
| 4 'Marked' | 14 (8) | 18 (1) | 39 (8) | 71 (3) |
| 5 'Severe' | 13 (8) | 7 (<1) | 45 (9) | 65 (3) |

Data are presented as *n* (%). Abbreviations: BPRS, Brief Psychiatric Rating Scale; PANSS, Positive and Negative Syndrome Scale; SAPS, Scale for the Assessment of Positive Symptoms.

#### **Appendix 3**
Primary analyses

**Table A3.1.** Group differences in global network topology.

| Metric | M (95% CI) | | | ΔAUC, $p_{BH}$ | | |
|---|---|---|---|---|---|---|
| | SZ+A (*N* = 231) | SZ-A (*N* = 2029) | HC (*N* = 3058) | SZ+A vs SZ-A | SZ+A vs HC | SZ-A vs HC |
| Transitivity | 0.47 (0.40, 0.51) | 0.56 (0.51, 0.58) | 0.50 (0.46, 0.52) | −0.014, .134 | −0.005, .938 | **0.009, .015** |
| Modularity | 0.39 (0.31, 0.42) | 0.30 (0.27, 0.33) | 0.39 (0.36, 0.42) | **0.013, .012** | −0.001, .938 | **−0.015, < .001** |
| Efficiency | 0.47 (0.44, 0.51) | 0.39 (0.37, 0.44) | 0.45 (0.44, 0.48) | 0.012, .084 | 0.003, .938 | **−0.009, .001** |
| Small-worldness | 1.57 (1.32, 1.81) | 1.14 (1.11, 1.29) | 1.47 (1.33, 1.65) | **0.064, .012** | 0.017, .938 | **−0.047, .005** |

Means were calculated across a range of density thresholds ($K$ = 0.10–0.25, $\Delta K$ = 0.01) and are reported with 95% bootstrap confidence intervals (CIs; 5000 resamples). Group differences in area under the curve (ΔAUC) were assessed using permutation testing (10 000 permutations). Bold values indicate statistically significant group differences after Benjamini–Hochberg correction for multiple comparisons ($p_{BH}$ < .05). Abbreviations: SZ+A, schizophrenia with aggression; SZ−A, schizophrenia without aggression; HC, healthy controls.

**Table A3.2.** Group differences in degree.

| | M (95% CI) | | | ΔAUC, $p_{BH}$ | | |
|---|---|---|---|---|---|---|
| Node | SZ+A (*N* = 231) | SZ-A (*N* = 2029) | HC (*N* = 3058) | SZ+A vs SZ-A | SZ+A vs HC | SZ-A vs HC |
| FPOL | 2.1 (0.0, 7.8) | 0.0 (0.0, 0.4) | 1.0 (0.0, 2.1) | 0.324, .434 | 0.180, .889 | −0.145, .361 |
| POP | 3.5 (1.4, 9.5) | 3.7 (1.8, 7.2) | 3.9 (2.4, 6.4) | −0.025, .985 | −0.070, .929 | −0.045, .898 |
| PTRI | 5.0 (2.2, 8.5) | 4.6 (3.0, 6.2) | 5.1 (3.3, 6.1) | 0.050, .950 | −0.024, .948 | −0.074, .883 |
| PORB | 6.0 (2.8, 8.6) | 4.4 (3.1, 5.8) | 4.7 (3.4, 7.2) | 0.245, .759 | 0.200, .889 | −0.045, .883 |
| MOFC | 11.8 (7.0, 16.4) | 11.6 (8.4, 14.5) | 9.0 (6.5, 11.2) | 0.051, .976 | 0.432, .889 | 0.381, .580 |
| LOFC | 11.9 (7.6, 15.5) | 18.4 (14.5, 20.6) | 18.2 (13.4, 20.4) | −1.017, .672 | −0.981, .889 | 0.036, .924 |
| CMFG | 6.8 (3.4, 11.0) | 7.5 (5.2, 9.4) | 8.2 (6.8, 10.0) | −0.136, .913 | −0.215, .889 | −0.079, .883 |
| RMFG | 12.4 (7.2, 16.1) | 18.1 (13.9, 20.7) | 14.9 (10.5, 17.8) | −0.840, .672 | −0.361, .889 | 0.479, .580 |
| SFG | 16.6 (12.1, 19.1) | 20.4 (15.8, 23.2) | 17.9 (13.6, 21.3) | −0.573, .672 | −0.202, .889 | 0.371, .779 |
| PARL | 5.7 (2.1, 9.9) | 7.4 (4.8, 9.6) | 8.1 (5.4, 10.4) | −0.246, .856 | −0.346, .889 | −0.099, .883 |
| PREG | 7.8 (4.8, 13.4) | 11.5 (7.6, 15.2) | 9.3 (6.9, 12.4) | −0.565, .759 | −0.230, .889 | 0.335, .709 |
| CACC | 3.6 (2.4, 8.0) | 2.5 (2.0, 3.4) | 3.1 (2.1, 4.4) | 0.159, .759 | 0.070, .889 | −0.090, .580 |
| RACC | 8.9 (5.1, 11.9) | 11.5 (8.6, 13.3) | 8.5 (6.4, 11.2) | −0.381, .759 | 0.085, .929 | 0.466, .381 |
| PCC | 8.8 (3.3, 13.1) | 11.9 (8.2, 15.1) | 10.8 (7.9, 14.8) | −0.469, .759 | −0.298, .889 | 0.171, .883 |
| ISTH | 4.6 (0.8, 9.4) | 3.2 (1.6, 6.0) | 3.6 (2.6, 6.6) | 0.204, .897 | 0.149, .889 | −0.054, .883 |
| INS | 16.7 (9.5, 19.4) | 11.8 (8.4, 15.2) | 7.9 (6.2, 10.7) | 0.738, .672 | 1.327, .328 | 0.589, .231 |
| TPOL | 2.2 (0.0, 4.9) | 0.0 (0.0, 0.5) | 1.0 (0.9, 1.9) | 0.345, .434 | 0.195, .889 | **−0.150, .039** |
| ENTH | 2.4 (0.2, 4.1) | 0.2 (0.0, 1.2) | 2.0 (1.9, 2.6) | 0.329, .434 | 0.065, .889 | **−0.265, .003** |
| PHG | 4.4 (1.8, 6.9) | 1.8 (1.0, 3.1) | 3.4 (2.9, 5.4) | 0.384, .672 | 0.145, .889 | **−0.239, .039** |
| TVTG | 7.0 (4.8, 11.2) | 5.9 (3.6, 8.9) | 8.2 (5.6, 9.8) | 0.165, .913 | −0.179, .889 | −0.345, .490 |
| ITG | 10.1 (5.5, 15.0) | 6.2 (3.9, 8.8) | 8.1 (5.4, 11.1) | 0.575, .672 | 0.284, .889 | −0.291, .580 |
| MTG | 8.2 (4.9, 12.9) | 11.3 (8.2, 13.7) | 6.9 (5.4, 9.1) | −0.461, .759 | 0.195, .889 | 0.655, .071 |
| STG | 15.8 (9.9, 19.9) | 13.9 (11.1, 15.8) | 9.8 (7.8, 12.4) | 0.261, .881 | 0.901, .889 | 0.640, .093 |
| BANKS | 3.6 (2.0, 5.8) | 3.2 (3.0, 4.6) | 3.6 (3.0, 4.7) | 0.070, .913 | 0.015, .948 | −0.055, .850 |
| FUSI | 13.4 (6.9, 17.8) | 12.2 (8.9, 15.6) | 10.7 (6.8, 14.6) | 0.206, .913 | 0.424, .889 | 0.218, .883 |

| | | | | | | |
|---|---|---|---|---|---|---|
| POSG | 12.2 (6.8, 16.4) | 9.0 (6.0, 12.7) | 8.2 (5.9, 11.2) | 0.502, .759 | 0.598, .889 | 0.096, .883 |
| PREC | 10.7 (5.2, 14.6) | 17.6 (12.4, 20.6) | 18.9 (15.2, 21.0) | −1.051, .672 | −1.252, .889 | −0.201, .850 |
| SMG | 8.2 (4.8, 12.0) | 7.6 (4.9, 10.8) | 8.6 (6.2, 10.6) | 0.082, .950 | −0.066, .929 | −0.148, .850 |
| IPL | 4.7 (2.8, 9.2) | 9.7 (6.9, 12.4) | 9.2 (7.0, 11.7) | −0.738, .672 | −0.679, .889 | 0.060, .888 |
| SPL | 5.0 (2.8, 9.8) | 5.5 (3.2, 8.8) | 6.8 (4.2, 9.8) | −0.085, .950 | −0.262, .889 | −0.177, .850 |
| LING | 3.6 (2.1, 6.4) | 3.2 (3.0, 3.9) | 4.4 (4.0, 5.8) | 0.055, .913 | −0.120, .889 | **−0.175, .039** |
| CUN | 3.8 (2.9, 7.2) | 3.0 (3.0, 4.2) | 5.1 (4.1, 6.0) | 0.115, .763 | −0.201, .889 | **−0.315, .039** |
| PERI | 2.9 (2.0, 4.3) | 2.1 (2.0, 2.7) | 3.4 (2.9, 3.9) | 0.135, .672 | −0.075, .889 | **−0.210, .003** |
| LOC | 3.8 (1.6, 9.4) | 5.7 (4.2, 8.0) | 6.1 (4.4, 8.8) | −0.293, .759 | −0.350, .889 | −0.057, .883 |
| HIPPO | 3.0 (2.1, 7.4) | 3.1 (2.2, 4.9) | 4.4 (3.4, 5.5) | −0.005, .985 | −0.220, .889 | −0.215, .319 |
| AMYG | 5.9 (2.5, 10.3) | 2.9 (2.1, 5.3) | 4.2 (3.2, 6.2) | 0.434, .672 | 0.244, .889 | −0.190, .580 |
| CAUD | 2.9 (1.5, 4.0) | 2.8 (2.1, 3.6) | 3.6 (2.8, 4.4) | 0.035, .950 | −0.100, .889 | **−0.135, .039** |
| PUT | 6.8 (3.6, 9.4) | 4.4 (3.4, 5.4) | 5.8 (4.9, 6.4) | 0.365, .434 | 0.150, .889 | −0.215, .231 |
| NACC | 6.2 (2.6, 11.2) | 1.9 (1.1, 2.8) | 4.4 (3.3, 5.2) | 0.635, .246 | 0.265, .889 | **−0.370, .003** |
| PALL | 3.4 (2.2, 7.1) | 2.6 (2.0, 3.0) | 2.0 (1.6, 2.0) | 0.120, .759 | 0.205, .889 | **0.085, .004** |
| THAL | 4.6 (2.1, 8.4) | 2.6 (1.5, 4.9) | 3.9 (2.4, 5.6) | 0.305, .759 | 0.105, .889 | −0.200, .580 |

Means were calculated across a range of density thresholds ($K = 0.10–0.25$, $\Delta K = 0.01$) and are reported with 95% bootstrap confidence intervals (CIs; 5000 resamples). Group differences in area under the curve (ΔAUC) were assessed using permutation testing (10 000 permutations). Bold values indicate statistically significant group differences after Benjamini–Hochberg correction for multiple comparisons ($p_{BH} < .05$). Abbreviations: SZ+A, schizophrenia with aggression; SZ−A, schizophrenia without aggression; HC, healthy controls; FPOL, frontal pole; POP, pars opercularis; PTRI, pars triangularis; PORB, pars orbitalis; MOFC, medial orbitofrontal cortex; LOFC, lateral orbitofrontal cortex; CMFG, caudal middle frontal gyrus; RMFG, rostral middle frontal gyrus; SFG, superior frontal gyrus; PARL, paracentral lobule; PREG, precentral gyrus; CACC, caudal anterior cingulate cortex; RACC, rostral anterior cingulate cortex; PCC, posterior cingulate cortex; ISTH, isthmus of the cingulate cortex; INS, insular cortex; TPOL, temporal pole; ENTH, entorhinal cortex; PHG, parahippocampal gyrus; TVTG, transverse temporal gyrus; ITG, inferior temporal gyrus; MTG, middle temporal gyrus; STG, superior temporal gyrus; BANKS, banks of the superior temporal sulcus; FUSI, fusiform gyrus; POSG, postcentral gyrus; PREC, precuneus; SMG, supramarginal gyrus; IPL, inferior parietal lobule; SPL, superior parietal lobule; LING, lingual gyrus; CUN, cuneus; PERI, pericalcarine cortex; LOC, lateral occipital cortex; HIPPO, hippocampus; AMYG, amygdala; CAUD, caudate nucleus; PUT, putamen; NACC, nucleus accumbens; PALL, globus pallidus; THAL, thalamus.

**Table A3.3.** Group differences in betweenness centrality.

| | M (95% CI) | | | ΔAUC, $p_{BH}$ | | |
|---|---|---|---|---|---|---|
| Node | SZ+A (*N* = 231) | SZ-A (*N* = 2029) | HC (*N* = 3058) | SZ+A vs SZ-A | SZ+A vs HC | SZ-A vs HC |
| FPOL | 0.000 (0.000, 0.013) | 0.000 (0.000, 0.000) | 0.000 (0.000, 0.000) | 0.00000, 1.000 | 0.00000, 1.000 | 0.00000, 1.000 |
| POP | 0.001 (0.000, 0.034) | 0.001 (0.000, 0.009) | 0.001 (0.000, 0.007) | −0.00002, 1.000 | −0.00001, 1.000 | 0.00001, 1.000 |
| PTRI | 0.015 (0.001, 0.044) | 0.008 (0.001, 0.029) | 0.004 (0.001, 0.022) | 0.00111, .942 | 0.00166, .848 | 0.00055, 1.000 |
| PORB | 0.005 (0.000, 0.033) | 0.010 (0.000, 0.035) | 0.002 (0.000, 0.027) | −0.00043, .942 | 0.00047, .990 | 0.00090, .805 |
| MOFC | 0.038 (0.009, 0.131) | 0.006 (0.002, 0.028) | 0.018 (0.002, 0.043) | 0.00496, .723 | 0.00311, .848 | −0.00185, .670 |
| LOFC | 0.040 (0.011, 0.137) | 0.067 (0.029, 0.124) | 0.117 (0.037, 0.192) | −0.00428, .942 | −0.01150, .848 | −0.00722, .694 |
| CMFG | 0.003 (0.000, 0.050) | 0.001 (0.000, 0.005) | 0.016 (0.002, 0.028) | 0.00022, .942 | −0.00186, .848 | −0.00208, .116 |
| RMFG | 0.054 (0.012, 0.117) | 0.080 (0.031, 0.151) | 0.069 (0.022, 0.131) | −0.00384, .942 | −0.00237, .990 | 0.00147, 1.000 |
| SFG | 0.086 (0.033, 0.156) | 0.174 (0.034, 0.286) | 0.081 (0.030, 0.210) | −0.01257, .784 | 0.00135, .990 | 0.01392, .670 |
| PARL | 0.002 (0.000, 0.024) | 0.001 (0.000, 0.004) | 0.004 (0.001, 0.012) | 0.00012, .942 | −0.00038, .990 | −0.00049, .259 |
| PREG | 0.004 (0.001, 0.083) | 0.009 (0.002, 0.061) | 0.011 (0.004, 0.068) | −0.00079, .942 | −0.00085, .990 | −0.00007, 1.000 |
| CACC | 0.001 (0.000, 0.027) | 0.000 (0.000, 0.000) | 0.000 (0.000, 0.000) | 0.00017, .777 | 0.00017, .962 | 0.00000, 1.000 |
| RACC | 0.050 (0.005, 0.090) | 0.016 (0.008, 0.030) | 0.011 (0.005, 0.040) | 0.00491, .553 | 0.00571, .848 | 0.00079, .895 |
| PCC | 0.031 (0.003, 0.103) | 0.019 (0.009, 0.046) | 0.036 (0.012, 0.119) | 0.00209, .942 | −0.00066, .990 | −0.00275, .670 |
| ISTH | 0.064 (0.000, 0.125) | 0.007 (0.000, 0.026) | 0.022 (0.007, 0.056) | 0.00885, .553 | 0.00687, .848 | −0.00199, .521 |
| INS | 0.139 (0.024, 0.231) | 0.012 (0.004, 0.118) | 0.005 (0.002, 0.052) | 0.01953, .553 | 0.02067, .848 | 0.00115, .670 |
| TPOL | 0.004 (0.000, 0.043) | 0.000 (0.000, 0.000) | 0.000 (0.000, 0.013) | 0.00062, .723 | 0.00062, .886 | 0.00000, 1.000 |
| ENTH | 0.003 (0.000, 0.035) | 0.000 (0.000, 0.018) | 0.049 (0.018, 0.050) | 0.00054, .784 | −0.00686, .848 | **−0.00740, .008** |
| PHG | 0.027 (0.002, 0.111) | 0.081 (0.000, 0.184) | 0.297 (0.131, 0.346) | −0.00883, 1.000 | −0.04093, .848 | −0.03210, .116 |
| TVTG | 0.002 (0.001, 0.032) | 0.001 (0.000, 0.009) | 0.008 (0.002, 0.026) | 0.00010, .956 | −0.00096, .990 | −0.00106, .521 |
| ITG | 0.050 (0.013, 0.158) | 0.000 (0.000, 0.011) | 0.086 (0.006, 0.173) | 0.00749, .642 | −0.00515, .990 | −0.01264, .122 |
| MTG | 0.031 (0.007, 0.107) | 0.016 (0.007, 0.083) | 0.012 (0.004, 0.050) | 0.00227, .942 | 0.00283, .848 | 0.00057, 1.000 |
| STG | 0.078 (0.019, 0.228) | 0.032 (0.019, 0.072) | 0.037 (0.017, 0.088) | 0.00681, .723 | 0.00609, .848 | −0.00072, 1.000 |
| BANKS | 0.001 (0.000, 0.015) | 0.000 (0.000, 0.001) | 0.001 (0.000, 0.002) | 0.00012, .942 | 0.00003, .990 | −0.00008, .116 |
| FUSI | 0.233 (0.069, 0.304) | 0.152 (0.056, 0.279) | 0.322 (0.157, 0.382) | 0.01303, .942 | −0.01294, .990 | −0.02597, .238 |

| | | | | | | |
|---|---|---|---|---|---|---|
| POSG | 0.036 (0.008, 0.128) | 0.005 (0.002, 0.018) | 0.013 (0.003, 0.035) | 0.00465, .642 | 0.00344, .848 | −0.00121, .670 |
| PREC | 0.084 (0.008, 0.159) | 0.083 (0.038, 0.195) | 0.255 (0.128, 0.336) | 0.00003, 1.000 | −0.02495, .848 | −0.02498, .122 |
| SMG | 0.009 (0.002, 0.048) | 0.003 (0.001, 0.014) | 0.011 (0.004, 0.027) | 0.00091, .942 | −0.00027, .990 | −0.00118, .238 |
| IPL | 0.008 (0.001, 0.053) | 0.013 (0.006, 0.030) | 0.029 (0.013, 0.111) | −0.00067, .942 | −0.00310, .886 | −0.00243, .536 |
| SPL | 0.024 (0.000, 0.074) | 0.001 (0.000, 0.021) | 0.019 (0.003, 0.045) | 0.00342, .723 | 0.00117, .990 | −0.00225, .515 |
| LING | 0.031 (0.000, 0.083) | 0.021 (0.007, 0.027) | 0.010 (0.002, 0.056) | 0.00163, .942 | 0.00365, .848 | 0.00202, .339 |
| CUN | 0.014 (0.001, 0.092) | 0.018 (0.007, 0.034) | 0.020 (0.005, 0.039) | −0.00058, .956 | −0.00059, .990 | −0.00001, 1.000 |
| PERI | 0.000 (0.000, 0.014) | 0.000 (0.000, 0.000) | 0.000 (0.000, 0.001) | 0.00001, .742 | −0.00002, .990 | **−0.00003, .037** |
| LOC | 0.039 (0.001, 0.130) | 0.113 (0.067, 0.131) | 0.051 (0.019, 0.100) | −0.01105, .777 | −0.00214, .990 | 0.00891, .116 |
| HIPPO | 0.002 (0.000, 0.073) | 0.139 (0.003, 0.186) | 0.173 (0.054, 0.228) | −0.02082, .784 | −0.02605, .848 | −0.00523, .933 |
| AMYG | 0.025 (0.002, 0.129) | 0.046 (0.007, 0.145) | 0.104 (0.015, 0.176) | −0.00352, .942 | −0.01099, .900 | −0.00747, .694 |
| CAUD | 0.001 (0.000, 0.016) | 0.001 (0.000, 0.010) | 0.001 (0.001, 0.014) | 0.00014, .942 | 0.00007, .990 | −0.00007, 1.000 |
| PUT | 0.058 (0.010, 0.108) | 0.043 (0.010, 0.097) | 0.074 (0.042, 0.092) | 0.00221, .942 | −0.00250, .990 | −0.00471, .410 |
| NACC | 0.067 (0.005, 0.147) | 0.001 (0.000, 0.010) | 0.013 (0.003, 0.061) | 0.00932, .553 | 0.00753, .848 | −0.00179, .670 |
| PALL | 0.002 (0.000, 0.045) | 0.008 (0.000, 0.030) | 0.000 (0.000, 0.000) | −0.00102, .942 | 0.00028, .848 | **0.00129, .004** |
| THAL | 0.032 (0.001, 0.097) | 0.030 (0.000, 0.112) | 0.019 (0.002, 0.115) | 0.00028, 1.000 | 0.00202, .990 | 0.00174, 1.000 |

Means were calculated across a range of density thresholds ($K$ = 0.10–0.25, $\Delta K$ = 0.01) and are reported with 95% bootstrap confidence intervals (CIs; 5000 resamples). Group differences in area under the curve (ΔAUC) were assessed using permutation testing (10 000 permutations). Bold values indicate statistically significant group differences after Benjamini–Hochberg correction for multiple comparisons ($p_{BH} < .05$). Abbreviations: SZ+A, schizophrenia with aggression; SZ−A, schizophrenia without aggression; HC, healthy controls; FPOL, frontal pole; POP, pars opercularis; PTRI, pars triangularis; PORB, pars orbitalis; MOFC, medial orbitofrontal cortex; LOFC, lateral orbitofrontal cortex; CMFG, caudal middle frontal gyrus; RMFG, rostral middle frontal gyrus; SFG, superior frontal gyrus; PARL, paracentral lobule; PREG, precentral gyrus; CACC, caudal anterior cingulate cortex; RACC, rostral anterior cingulate cortex; PCC, posterior cingulate cortex; ISTH, isthmus of the cingulate cortex; INS, insular cortex; TPOL, temporal pole; ENTH, entorhinal cortex; PHG, parahippocampal gyrus; TVTG, transverse temporal gyrus; ITG, inferior temporal gyrus; MTG, middle temporal gyrus; STG, superior temporal gyrus; BANKS, banks of the superior temporal sulcus; FUSI, fusiform gyrus; POSG, postcentral gyrus; PREC, precuneus; SMG, supramarginal gyrus; IPL, inferior parietal lobule; SPL, superior parietal lobule; LING, lingual gyrus; CUN, cuneus; PERI, pericalcarine cortex; LOC, lateral occipital cortex; HIPPO, hippocampus; AMYG, amygdala; CAUD, caudate nucleus; PUT, putamen; NACC, nucleus accumbens; PALL, globus pallidus; THAL, thalamus.

**Table A3.4.** Group differences in harmonic centrality.

| | M (95% CI) | | | ΔAUC, $p_{BH}$ | | |
|---|---|---|---|---|---|---|
| Node | SZ+A (*N* = 231) | SZ-A (*N* = 2029) | HC (*N* = 3058) | SZ+A vs SZ-A | SZ+A vs HC | SZ-A vs HC |
| FPOL | 0.384 (0.000, 0.519) | 0.000 (0.000, 0.160) | 0.204 (0.000, 0.377) | 0.059, .465 | 0.029, .928 | −0.030, .337 |
| POP | 0.408 (0.309, 0.542) | 0.388 (0.305, 0.469) | 0.412 (0.359, 0.472) | 0.003, .904 | −0.001, .976 | −0.003, .764 |
| PTRI | 0.463 (0.346, 0.531) | 0.410 (0.354, 0.458) | 0.441 (0.393, 0.469) | 0.008, .787 | 0.003, .928 | −0.005, .484 |
| PORB | 0.482 (0.408, 0.539) | 0.418 (0.390, 0.457) | 0.445 (0.423, 0.496) | 0.010, .588 | 0.006, .928 | −0.004, .371 |
| MOFC | 0.590 (0.507, 0.661) | 0.525 (0.486, 0.575) | 0.520 (0.480, 0.562) | 0.010, .588 | 0.011, .858 | 0.001, .940 |
| LOFC | 0.595 (0.520, 0.654) | 0.620 (0.572, 0.666) | 0.655 (0.590, 0.696) | −0.004, .828 | −0.009, .909 | −0.005, .484 |
| CMFG | 0.490 (0.432, 0.566) | 0.463 (0.435, 0.501) | 0.495 (0.479, 0.538) | 0.004, .828 | −0.001, .976 | −0.005, .306 |
| RMFG | 0.586 (0.507, 0.650) | 0.622 (0.568, 0.670) | 0.602 (0.539, 0.654) | −0.005, .828 | −0.002, .976 | 0.003, .764 |
| SFG | 0.638 (0.579, 0.692) | 0.657 (0.587, 0.718) | 0.648 (0.592, 0.711) | −0.003, .869 | −0.001, .976 | 0.001, .925 |
| PARL | 0.470 (0.400, 0.549) | 0.457 (0.423, 0.505) | 0.508 (0.465, 0.547) | 0.002, .869 | −0.005, .928 | −0.008, .253 |
| PREG | 0.498 (0.450, 0.609) | 0.524 (0.472, 0.590) | 0.527 (0.492, 0.580) | −0.004, .869 | −0.004, .928 | −0.000, .965 |
| CACC | 0.426 (0.389, 0.528) | 0.363 (0.338, 0.406) | 0.406 (0.370, 0.440) | 0.009, .698 | 0.003, .928 | −0.006, .217 |
| RACC | 0.552 (0.468, 0.600) | 0.523 (0.484, 0.562) | 0.508 (0.475, 0.565) | 0.005, .828 | 0.007, .928 | 0.002, .745 |
| PCC | 0.553 (0.409, 0.619) | 0.529 (0.480, 0.582) | 0.553 (0.505, 0.622) | 0.004, .828 | 0.000, .976 | −0.004, .643 |
| ISTH | 0.379 (0.159, 0.552) | 0.401 (0.359, 0.464) | 0.448 (0.425, 0.498) | −0.003, .903 | −0.010, .928 | −0.007, .266 |
| INS | 0.659 (0.549, 0.708) | 0.528 (0.485, 0.598) | 0.505 (0.482, 0.560) | 0.020, .465 | 0.023, .623 | 0.004, .609 |
| TPOL | 0.344 (0.000, 0.472) | 0.000 (0.000, 0.114) | 0.245 (0.226, 0.352) | 0.053, .465 | 0.016, .928 | **−0.037, .023** |
| ENTH | 0.345 (0.040, 0.445) | 0.076 (0.000, 0.259) | 0.320 (0.300, 0.377) | 0.042, .465 | 0.005, .928 | **−0.037, .023** |
| PHG | 0.440 (0.314, 0.505) | 0.362 (0.290, 0.413) | 0.434 (0.405, 0.481) | 0.012, .694 | 0.001, .976 | **−0.011, .040** |
| TVTG | 0.495 (0.467, 0.576) | 0.442 (0.403, 0.504) | 0.517 (0.470, 0.547) | 0.008, .787 | −0.003, .928 | −0.011, .115 |
| ITG | 0.560 (0.479, 0.645) | 0.450 (0.407, 0.505) | 0.529 (0.471, 0.589) | 0.017, .465 | 0.004, .928 | −0.012, .150 |
| MTG | 0.529 (0.458, 0.611) | 0.524 (0.479, 0.576) | 0.493 (0.457, 0.541) | 0.001, .928 | 0.005, .928 | 0.005, .509 |
| STG | 0.644 (0.560, 0.713) | 0.559 (0.528, 0.603) | 0.538 (0.506, 0.585) | 0.013, .465 | 0.016, .734 | 0.003, .574 |
| BANKS | 0.428 (0.361, 0.494) | 0.371 (0.362, 0.424) | 0.408 (0.390, 0.448) | 0.009, .763 | 0.003, .928 | −0.005, .253 |
| FUSI | 0.623 (0.506, 0.692) | 0.554 (0.505, 0.619) | 0.574 (0.516, 0.643) | 0.011, .694 | 0.008, .928 | −0.003, .745 |

| | | | | | | |
|---|---|---|---|---|---|---|
| POSG | 0.576 (0.496, 0.652) | 0.484 (0.445, 0.552) | 0.520 (0.478, 0.567) | 0.014, .516 | 0.009, .928 | −0.006, .475 |
| PREC | 0.584 (0.467, 0.642) | 0.616 (0.549, 0.675) | 0.676 (0.632, 0.716) | −0.005, .828 | −0.014, .827 | −0.009, .220 |
| SMG | 0.511 (0.454, 0.588) | 0.461 (0.422, 0.520) | 0.520 (0.484, 0.557) | 0.008, .792 | −0.001, .976 | −0.009, .160 |
| IPL | 0.462 (0.390, 0.556) | 0.502 (0.464, 0.558) | 0.538 (0.507, 0.585) | −0.006, .828 | −0.011, .827 | −0.006, .337 |
| SPL | 0.470 (0.396, 0.560) | 0.436 (0.395, 0.499) | 0.490 (0.447, 0.545) | 0.005, .828 | −0.003, .976 | −0.008, .337 |
| LING | 0.313 (0.203, 0.484) | 0.332 (0.309, 0.367) | 0.401 (0.372, 0.467) | −0.003, .883 | −0.013, .909 | **−0.010, .046** |
| CUN | 0.313 (0.220, 0.500) | 0.327 (0.309, 0.398) | 0.446 (0.391, 0.480) | −0.002, .904 | −0.020, .623 | **−0.018, .023** |
| PERI | 0.290 (0.188, 0.423) | 0.258 (0.242, 0.319) | 0.362 (0.338, 0.392) | 0.005, .828 | −0.011, .827 | **−0.016, .008** |
| LOC | 0.349 (0.226, 0.560) | 0.454 (0.413, 0.509) | 0.478 (0.417, 0.541) | −0.016, .516 | −0.020, .827 | −0.004, .607 |
| HIPPO | 0.367 (0.310, 0.522) | 0.276 (0.145, 0.439) | 0.377 (0.335, 0.453) | 0.014, .828 | −0.001, .976 | −0.015, .217 |
| AMYG | 0.455 (0.334, 0.573) | 0.235 (0.150, 0.416) | 0.366 (0.310, 0.468) | 0.033, .588 | 0.014, .928 | −0.019, .175 |
| CAUD | 0.357 (0.261, 0.415) | 0.188 (0.129, 0.287) | 0.278 (0.254, 0.347) | 0.026, .465 | 0.012, .928 | −0.014, .118 |
| PUT | 0.475 (0.352, 0.536) | 0.236 (0.155, 0.348) | 0.346 (0.309, 0.406) | 0.036, .312 | 0.020, .827 | −0.017, .118 |
| NACC | 0.491 (0.359, 0.582) | 0.180 (0.116, 0.285) | 0.323 (0.286, 0.400) | 0.047, .160 | 0.025, .827 | **−0.021, .033** |
| PALL | 0.370 (0.295, 0.489) | 0.198 (0.127, 0.296) | 0.258 (0.230, 0.291) | 0.026, .465 | 0.017, .827 | −0.009, .217 |
| THAL | 0.437 (0.301, 0.540) | 0.223 (0.137, 0.399) | 0.328 (0.290, 0.439) | 0.032, .588 | 0.017, .928 | −0.016, .276 |

Means were calculated across a range of density thresholds ($K$ = 0.10–0.25, $\Delta K$ = 0.01) and are reported with 95% bootstrap confidence intervals (CIs; 5000 resamples). Group differences in area under the curve (ΔAUC) were assessed using permutation testing (10 000 permutations). Bold values indicate statistically significant group differences after Benjamini–Hochberg correction for multiple comparisons ($p_{BH} < .05$). Abbreviations: SZ+A, schizophrenia with aggression; SZ−A, schizophrenia without aggression; HC, healthy controls; FPOL, frontal pole; POP, pars opercularis; PTRI, pars triangularis; PORB, pars orbitalis; MOFC, medial orbitofrontal cortex; LOFC, lateral orbitofrontal cortex; CMFG, caudal middle frontal gyrus; RMFG, rostral middle frontal gyrus; SFG, superior frontal gyrus; PARL, paracentral lobule; PREG, precentral gyrus; CACC, caudal anterior cingulate cortex; RACC, rostral anterior cingulate cortex; PCC, posterior cingulate cortex; ISTH, isthmus of the cingulate cortex; INS, insular cortex; TPOL, temporal pole; ENTH, entorhinal cortex; PHG, parahippocampal gyrus; TVTG, transverse temporal gyrus; ITG, inferior temporal gyrus; MTG, middle temporal gyrus; STG, superior temporal gyrus; BANKS, banks of the superior temporal sulcus; FUSI, fusiform gyrus; POSG, postcentral gyrus; PREC, precuneus; SMG, supramarginal gyrus; IPL, inferior parietal lobule; SPL, superior parietal lobule; LING, lingual gyrus; CUN, cuneus; PERI, pericalcarine cortex; LOC, lateral occipital cortex; HIPPO, hippocampus; AMYG, amygdala; CAUD, caudate nucleus; PUT, putamen; NACC, nucleus accumbens; PALL, globus pallidus; THAL, thalamus.

**Table A3.5.** Group differences in within-module degree.

| Node | M (95% CI) | | | ΔAUC, $p_{BH}$ | | |
|---|---|---|---|---|---|---|
| | SZ+A (*N* = 231) | SZ-A (*N* = 2029) | HC (*N* = 3058) | SZ+A vs SZ-A | SZ+A vs HC | SZ-A vs HC |
| FPOL | −1.52 (−1.85, 0.00) | 0.00 (0.00, 0.00) | −1.76 (−1.87, 0.00) | −0.226, .827 | 0.037, .991 | 0.263, .198 |
| POP | −1.22 (−1.61, 0.36) | −1.15 (−1.43, −0.23) | −1.07 (−1.46, −0.50) | −0.011, .991 | −0.024, .991 | −0.013, .917 |
| PTRI | −0.77 (−1.43, 0.21) | −0.94 (−1.18, 0.72) | −0.78 (−1.24, −0.50) | 0.023, .917 | −0.001, .991 | −0.024, .896 |
| PORB | −0.51 (−1.34, 0.16) | −0.99 (−1.20, −0.36) | −0.87 (−1.20, −0.35) | 0.070, .861 | 0.052, .991 | −0.018, .896 |
| MOFC | 0.87 (−0.36, 1.90) | 0.40 (−0.20, 1.01) | 0.08 (−0.50, 0.62) | 0.073, .908 | 0.119, .991 | 0.047, .896 |
| LOFC | 0.81 (−0.22, 1.62) | 1.78 (0.97, 2.20) | 2.10 (1.13, 2.68) | −0.152, .827 | −0.199, .977 | −0.047, .896 |
| CMFG | −0.39 (−1.15, 0.68) | −0.38 (−0.79, 0.06) | −0.02 (−0.36, 0.37) | −0.006, .991 | −0.054, .991 | −0.048, .683 |
| RMFG | 1.07 (−0.26, 1.78) | 1.57 (0.80, 2.10) | 1.25 (0.38, 1.90) | −0.073, .908 | −0.026, .991 | 0.047, .896 |
| SFG | 2.04 (1.07, 2.66) | 1.99 (1.24, 2.50) | 2.22 (1.31, 2.85) | 0.012, .991 | −0.023, .991 | −0.035, .896 |
| PARL | −0.64 (−1.44, 0.39) | −0.43 (−0.86, 0.06) | −0.11 (−0.67, 0.46) | −0.030, .917 | −0.077, .991 | −0.047, .896 |
| PREG | −0.09 (−0.79, 1.18) | 0.38 (−0.34, 1.11) | 0.18 (−0.36, 0.85) | −0.074, .908 | −0.043, .991 | 0.031, .896 |
| CACC | −1.20 (−1.51, −0.02) | −1.34 (−1.53, −1.09) | −1.24 (−1.53, −0.93) | 0.019, .917 | 0.004, .991 | −0.016, .896 |
| RACC | −0.09 (−0.92, 0.74) | 0.41 (−0.14, 0.81) | −0.02 (−0.49, 0.62) | −0.073, .908 | −0.006, .991 | 0.067, .683 |
| PCC | 0.06 (−1.30, 1.00) | 0.49 (−0.22, 1.14) | 0.56 (−0.13, 1.33) | −0.060, .908 | −0.070, .991 | −0.009, .943 |
| ISTH | −0.67 (−1.59, 0.71) | −1.34 (−1.58, −0.81) | −1.38 (−1.63, −0.52) | 0.099, .885 | 0.110, .991 | 0.011, .915 |
| INS | 1.78 (0.22, 2.34) | 0.42 (−0.21, 1.05) | −0.11 (−0.52, 0.48) | 0.205, .827 | 0.287, .377 | 0.082, .683 |
| TPOL | 0.00 (−1.74, 0.00) | 0.00 (0.00, 0.00) | −0.58 (−0.61, 0.00) | 0.000, 1.000 | 0.087, .991 | 0.087, .683 |
| ENTH | 0.00 (−1.88, 0.87) | 0.00 (−1.52, 0.00) | 1.15 (0.00, 1.15) | 0.000, 1.000 | −0.173, .991 | −0.173, .453 |
| PHG | −1.01 (−1.69, 0.98) | 0.00 (−1.78, 0.01) | −0.58 (−1.04, −0.43) | −0.153, .908 | −0.066, .991 | 0.087, .683 |
| TVTG | −0.29 (−0.80, 0.74) | −0.69 (−1.12, −0.09) | −0.11 (−0.71, 0.27) | 0.060, .908 | −0.030, .991 | −0.089, .453 |
| ITG | 0.12 (−0.81, 1.50) | −0.64 (−1.09, −0.10) | −0.20 (−0.77, 0.37) | 0.114, .827 | 0.047, .991 | −0.067, .683 |
| MTG | −0.11 (−0.75, 0.98) | 0.38 (−0.21, 0.84) | −0.36 (−0.71, 0.12) | −0.074, .908 | 0.036, .991 | 0.110, .329 |
| STG | 1.74 (0.35, 2.50) | 0.90 (0.40, 1.29) | 0.31 (−0.14, 0.88) | 0.121, .827 | 0.214, .715 | 0.093, .453 |
| BANKS | −1.14 (−1.52, 0.30) | −1.20 (−1.33, −0.89) | −1.12 (−1.33, −0.85) | 0.010, .991 | −0.002, .991 | −0.012, .896 |
| FUSI | 0.07 (−0.96, 1.91) | 0.17 (−0.44, 0.93) | −0.01 (−0.77, 0.77) | −0.009, .991 | 0.013, .991 | 0.023, .917 |

| | | | | | | |
|---|---|---|---|---|---|---|
| POSG | 0.89 (−0.36, 1.82) | −0.12 (−0.63, 0.65) | −0.10 (−0.62, 0.54) | 0.155, .827 | 0.150, .977 | −0.006, .944 |
| PREC | 0.35 (−0.96, 1.30) | 1.46 (0.51, 1.97) | 1.78 (1.11, 2.51) | −0.171, .827 | −0.218, .977 | −0.047, .896 |
| SMG | 0.00 (−0.82, 0.95) | −0.39 (−0.87, 0.29) | −0.01 (−0.51, 0.48) | 0.058, .908 | −0.001, .991 | −0.059, .683 |
| IPL | −0.84 (−1.22, 0.88) | 0.05 (−0.48, 0.59) | 0.14 (−0.34, 0.68) | −0.132, .827 | −0.149, .977 | −0.017, .915 |
| SPL | −0.97 (−1.43, 0.28) | −0.79 (−1.22, −0.20) | −0.73 (−1.30, −0.01) | −0.030, .934 | −0.036, .991 | −0.006, .943 |
| LING | 0.61 (−0.87, 1.17) | 0.81 (0.00, 1.02) | 0.92 (−0.74, 1.10) | −0.032, .917 | −0.048, .991 | −0.016, .896 |
| CUN | 0.56 (−0.62, 1.31) | 0.81 (0.00, 0.87) | −0.20 (−0.63, 0.38) | −0.042, .917 | 0.117, .977 | 0.159, .056 |
| PERI | 0.06 (−1.16, 0.78) | −0.81 (−0.87, 0.00) | 0.40 (−1.07, 0.65) | 0.135, .827 | −0.055, .991 | −0.189, .056 |
| LOC | −0.56 (−1.46, 0.61) | −0.81 (−0.89, 0.11) | 0.26 (−0.40, 0.62) | 0.042, .908 | −0.124, .981 | −0.166, .153 |
| HIPPO | −0.62 (−1.29, 1.11) | 0.29 (−1.07, 1.31) | −0.31 (−0.80, 0.37) | −0.136, .908 | −0.049, .991 | 0.087, .846 |
| AMYG | 0.84 (−1.01, 1.35) | −0.14 (−0.54, 0.78) | −0.18 (−0.45, 0.46) | 0.151, .827 | 0.160, .977 | 0.009, .943 |
| CAUD | −0.68 (−1.52, 0.55) | 0.60 (−0.41, 0.81) | −0.13 (−0.48, 0.36) | −0.190, .827 | −0.077, .991 | 0.113, .453 |
| PUT | 1.71 (−0.22, 2.02) | 0.98 (0.89, 1.92) | 1.59 (1.27, 1.84) | 0.113, .827 | 0.019, .991 | −0.095, .453 |
| NACC | −0.27 (−1.34, 1.24) | −0.98 (−1.31, −0.26) | 0.46 (−0.05, 0.81) | 0.103, .827 | −0.112, .991 | −0.215, .056 |
| PALL | −0.27 (−0.95, 1.13) | −0.60 (−0.82, 0.29) | −1.45 (−1.73, −1.03) | 0.048, .908 | 0.177, .977 | 0.130, .056 |
| THAL | 0.31 (−1.24, 1.07) | −0.14 (−0.74, 0.69) | 0.02 (−0.82, 0.45) | 0.065, .908 | 0.035, .991 | −0.030, .896 |

Means were calculated across a range of density thresholds ($K$ = 0.10–0.25, $\Delta K$ = 0.01) and are reported with 95% bootstrap confidence intervals (CIs; 5000 resamples). Group differences in area under the curve (ΔAUC) were assessed using permutation testing (10 000 permutations). Abbreviations: SZ+A, schizophrenia with aggression; SZ−A, schizophrenia without aggression; HC, healthy controls; FPOL, frontal pole; POP, pars opercularis; PTRI, pars triangularis; PORB, pars orbitalis; MOFC, medial orbitofrontal cortex; LOFC, lateral orbitofrontal cortex; CMFG, caudal middle frontal gyrus; RMFG, rostral middle frontal gyrus; SFG, superior frontal gyrus; PARL, paracentral lobule; PREG, precentral gyrus; CACC, caudal anterior cingulate cortex; RACC, rostral anterior cingulate cortex; PCC, posterior cingulate cortex; ISTH, isthmus of the cingulate cortex; INS, insular cortex; TPOL, temporal pole; ENTH, entorhinal cortex; PHG, parahippocampal gyrus; TVTG, transverse temporal gyrus; ITG, inferior temporal gyrus; MTG, middle temporal gyrus; STG, superior temporal gyrus; BANKS, banks of the superior temporal sulcus; FUSI, fusiform gyrus; POSG, postcentral gyrus; PREC, precuneus; SMG, supramarginal gyrus; IPL, inferior parietal lobule; SPL, superior parietal lobule; LING, lingual gyrus; CUN, cuneus; PERI, pericalcarine cortex; LOC, lateral occipital cortex; HIPPO, hippocampus; AMYG, amygdala; CAUD, caudate nucleus; PUT, putamen; NACC, nucleus accumbens; PALL, globus pallidus; THAL, thalamus.

**Table A3.6.** Group differences in participation coefficient.

| | M (95% CI) | | | ΔAUC, $p_{BH}$ | | |
|---|---|---|---|---|---|---|
| Node | SZ+A (*N* = 231) | SZ-A (*N* = 2029) | HC (*N* = 3058) | SZ+A vs SZ-A | SZ+A vs HC | SZ-A vs HC |
| FPOL | 0.00 (0.00, 0.16) | 0.00 (0.00, 0.00) | 0.00 (0.00, 0.00) | 0.000, 1.000 | 0.000, 1.000 | 0.000, 1.000 |
| POP | 0.00 (0.00, 0.28) | 0.00 (0.00, 0.20) | 0.00 (0.00, 0.00) | 0.000, 1.000 | 0.000, 1.000 | 0.000, 1.000 |
| PTRI | 0.00 (0.00, 0.36) | 0.00 (0.00, 0.33) | 0.00 (0.00, 0.00) | 0.000, 1.000 | 0.000, 1.000 | 0.000, 1.000 |
| PORB | 0.00 (0.00, 0.22) | 0.00 (0.00, 0.45) | 0.02 (0.00, 0.16) | 0.000, 1.000 | −0.001, .968 | −0.001, .722 |
| MOFC | 0.00 (0.00, 0.41) | 0.00 (0.00, 0.05) | 0.01 (0.00, 0.11) | 0.000, 1.000 | −0.001, 1.000 | −0.001, 1.000 |
| LOFC | 0.04 (0.00, 0.40) | 0.00 (0.00, 0.17) | 0.06 (0.00, 0.11) | 0.005, .762 | −0.004, 1.000 | −0.009, .722 |
| CMFG | 0.01 (0.00, 0.24) | 0.00 (0.00, 0.00) | 0.00 (0.00, 0.07) | 0.001, .562 | 0.001, .968 | 0.000, 1.000 |
| RMFG | 0.00 (0.00, 0.36) | 0.06 (0.00, 0.20) | 0.09 (0.00, 0.18) | −0.010, .693 | −0.013, .968 | −0.003, 1.000 |
| SFG | 0.02 (0.00, 0.21) | 0.09 (0.01, 0.19) | 0.00 (0.00, 0.10) | −0.010, .762 | 0.003, 1.000 | 0.013, .722 |
| PARL | 0.00 (0.00, 0.21) | 0.00 (0.00, 0.00) | 0.00 (0.00, 0.00) | 0.000, 1.000 | 0.000, 1.000 | 0.000, 1.000 |
| PREG | 0.02 (0.00, 0.27) | 0.00 (0.00, 0.07) | 0.00 (0.00, 0.08) | 0.002, .822 | 0.002, .968 | 0.000, 1.000 |
| CACC | 0.13 (0.00, 0.44) | 0.00 (0.00, 0.00) | 0.00 (0.00, 0.00) | 0.019, .138 | 0.019, .698 | 0.000, 1.000 |
| RACC | 0.21 (0.00, 0.48) | 0.00 (0.00, 0.00) | 0.02 (0.00, 0.12) | 0.031, .138 | 0.029, .698 | −0.002, .351 |
| PCC | 0.04 (0.00, 0.47) | 0.00 (0.00, 0.03) | 0.00 (0.00, 0.10) | 0.005, .635 | 0.005, 1.000 | 0.000, 1.000 |
| ISTH | 0.35 (0.00, 0.53) | 0.13 (0.00, 0.38) | 0.49 (0.00, 0.50) | 0.035, .635 | −0.019, 1.000 | −0.054, .219 |
| INS | 0.11 (0.00, 0.38) | 0.00 (0.00, 0.13) | 0.00 (0.00, 0.09) | 0.017, .635 | 0.017, .968 | 0.000, 1.000 |
| TPOL | 0.38 (0.00, 0.56) | 0.00 (0.00, 0.00) | 0.00 (0.00, 0.38) | 0.058, .257 | 0.058, .698 | 0.000, 1.000 |
| ENTH | 0.49 (0.00, 0.58) | 0.00 (0.00, 0.20) | 0.00 (0.00, 0.50) | 0.075, .138 | 0.075, .698 | 0.000, 1.000 |
| PHG | 0.53 (0.00, 0.63) | 0.32 (0.00, 0.55) | 0.66 (0.60, 0.72) | 0.031, .850 | −0.018, 1.000 | **−0.050, .002** |
| TVTG | 0.00 (0.00, 0.23) | 0.00 (0.00, 0.09) | 0.00 (0.00, 0.02) | 0.000, 1.000 | 0.000, 1.000 | 0.000, 1.000 |
| ITG | 0.17 (0.00, 0.63) | 0.00 (0.00, 0.04) | 0.05 (0.00, 0.24) | 0.026, .591 | 0.019, .968 | −0.007, .722 |
| MTG | 0.09 (0.00, 0.59) | 0.00 (0.00, 0.08) | 0.00 (0.00, 0.10) | 0.014, .635 | 0.014, .968 | 0.000, 1.000 |
| STG | 0.08 (0.00, 0.40) | 0.00 (0.00, 0.07) | 0.00 (0.00, 0.08) | 0.012, .635 | 0.012, .968 | 0.000, 1.000 |
| BANKS | 0.00 (0.00, 0.48) | 0.00 (0.00, 0.00) | 0.00 (0.00, 0.00) | 0.000, 1.000 | 0.000, 1.000 | 0.000, 1.000 |
| FUSI | 0.51 (0.01, 0.69) | 0.30 (0.13, 0.40) | 0.33 (0.22, 0.45) | 0.032, .635 | 0.027, .968 | −0.005, 1.000 |

| | | | | | | |
|---|---|---|---|---|---|---|
| POSG | 0.05 (0.00, 0.39) | 0.00 (0.00, 0.00) | 0.00 (0.00, 0.11) | 0.006, .635 | 0.006, .968 | 0.000, 1.000 |
| PREC | 0.14 (0.00, 0.51) | 0.08 (0.01, 0.20) | 0.26 (0.00, 0.33) | 0.008, .939 | −0.018, 1.000 | −0.027, .258 |
| SMG | 0.00 (0.00, 0.25) | 0.00 (0.00, 0.00) | 0.00 (0.00, 0.05) | 0.000, 1.000 | 0.000, 1.000 | 0.000, 1.000 |
| IPL | 0.00 (0.00, 0.55) | 0.00 (0.00, 0.06) | 0.00 (0.00, 0.14) | 0.000, 1.000 | 0.000, 1.000 | 0.000, 1.000 |
| SPL | 0.24 (0.00, 0.47) | 0.00 (0.00, 0.16) | 0.32 (0.00, 0.43) | 0.036, .591 | −0.013, 1.000 | −0.049, .237 |
| LING | 0.02 (0.00, 0.55) | 0.07 (0.00, 0.45) | 0.11 (0.00, 0.52) | −0.008, 1.000 | −0.014, 1.000 | −0.006, 1.000 |
| CUN | 0.11 (0.00, 0.49) | 0.00 (0.00, 0.45) | 0.46 (0.00, 0.48) | 0.015, .939 | −0.055, .968 | **−0.070, .015** |
| PERI | 0.00 (0.00, 0.30) | 0.00 (0.00, 0.00) | 0.00 (0.00, 0.19) | 0.000, 1.000 | 0.000, 1.000 | 0.000, 1.000 |
| LOC | 0.25 (0.00, 0.58) | 0.44 (0.40, 0.49) | 0.43 (0.00, 0.51) | −0.028, .635 | −0.029, .968 | −0.001, 1.000 |
| HIPPO | 0.00 (0.00, 0.55) | 0.32 (0.00, 0.48) | 0.35 (0.03, 0.48) | −0.048, .635 | −0.053, .698 | −0.004, 1.000 |
| AMYG | 0.21 (0.00, 0.59) | 0.44 (0.00, 0.54) | 0.18 (0.00, 0.42) | −0.036, .850 | 0.006, 1.000 | 0.042, .722 |
| CAUD | 0.00 (0.00, 0.39) | 0.02 (0.00, 0.12) | 0.00 (0.00, 0.00) | −0.002, .756 | 0.000, 1.000 | 0.002, .381 |
| PUT | 0.27 (0.00, 0.54) | 0.38 (0.00, 0.42) | 0.00 (0.00, 0.11) | −0.017, .939 | 0.042, .968 | 0.059, .237 |
| NACC | 0.49 (0.00, 0.63) | 0.11 (0.00, 0.22) | 0.00 (0.00, 0.15) | 0.057, .366 | 0.073, .698 | 0.016, .674 |
| PALL | 0.00 (0.00, 0.46) | 0.25 (0.00, 0.31) | 0.00 (0.00, 0.00) | −0.038, .641 | 0.000, 1.000 | **0.038, .002** |
| THAL | 0.20 (0.00, 0.57) | 0.26 (0.00, 0.41) | 0.00 (0.00, 0.26) | −0.009, 1.000 | 0.030, 1.000 | 0.040, .351 |

Means were calculated across a range of density thresholds ($K$ = 0.10–0.25, $\Delta K$ = 0.01) and are reported with 95% bootstrap confidence intervals (CIs; 5000 resamples). Group differences in area under the curve (ΔAUC) were assessed using permutation testing (10 000 permutations). Bold values indicate statistically significant group differences after Benjamini–Hochberg correction for multiple comparisons ($p_{BH} < .05$). Abbreviations: SZ+A, schizophrenia with aggression; SZ−A, schizophrenia without aggression; HC, healthy controls; FPOL, frontal pole; POP, pars opercularis; PTRI, pars triangularis; PORB, pars orbitalis; MOFC, medial orbitofrontal cortex; LOFC, lateral orbitofrontal cortex; CMFG, caudal middle frontal gyrus; RMFG, rostral middle frontal gyrus; SFG, superior frontal gyrus; PARL, paracentral lobule; PREG, precentral gyrus; CACC, caudal anterior cingulate cortex; RACC, rostral anterior cingulate cortex; PCC, posterior cingulate cortex; ISTH, isthmus of the cingulate cortex; INS, insular cortex; TPOL, temporal pole; ENTH, entorhinal cortex; PHG, parahippocampal gyrus; TVTG, transverse temporal gyrus; ITG, inferior temporal gyrus; MTG, middle temporal gyrus; STG, superior temporal gyrus; BANKS, banks of the superior temporal sulcus; FUSI, fusiform gyrus; POSG, postcentral gyrus; PREC, precuneus; SMG, supramarginal gyrus; IPL, inferior parietal lobule; SPL, superior parietal lobule; LING, lingual gyrus; CUN, cuneus; PERI, pericalcarine cortex; LOC, lateral occipital cortex; HIPPO, hippocampus; AMYG, amygdala; CAUD, caudate nucleus; PUT, putamen; NACC, nucleus accumbens; PALL, globus pallidus; THAL, thalamus.

**Table A3.7.** Group differences in canonical brain network density.

| Metric | M (95% CI) | | | ΔAUC, $p_{BH}$ | | |
|---|---|---|---|---|---|---|
| | SZ+A (*N* = 231) | SZ-A (*N* = 2029) | HC (*N* = 3058) | SZ+A vs SZ-A | SZ+A vs HC | SZ-A vs HC |
| *Salience network* | | | | | | |
| Network-rest | 0.26 (0.17, 0.31) | 0.18 (0.14, 0.23) | 0.14 (0.12, 0.18) | 0.011, .268 | 0.018, .083 | 0.006, .281 |
| *Central executive network* | | | | | | |
| Within-network | 0.26 (0.06, 0.48) | 0.51 (0.36, 0.62) | 0.29 (0.17, 0.51) | −0.037, .621 | −0.003, 1.000 | 0.034, .272 |
| Network-rest | 0.17 (0.14, 0.23) | 0.23 (0.20, 0.26) | 0.24 (0.21, 0.26) | −0.009, .268 | −0.010, .243 | −0.001, .852 |
| *Default mode network* | | | | | | |
| Within-network | 0.13 (0.10, 0.20) | 0.15 (0.13, 0.18) | 0.18 (0.15, 0.20) | −0.003, 1.000 | −0.006, 1.000 | −0.004, .341 |
| Network-rest | 0.15 (0.12, 0.17) | 0.15 (0.14, 0.16) | 0.15 (0.14, 0.16) | 0.000, .960 | 0.000, .998 | 0.000, .852 |
| *Language network* | | | | | | |
| Within-network | 0.36 (0.24, 0.52) | 0.38 (0.30, 0.41) | 0.31 (0.30, 0.38) | −0.003, 1.000 | 0.007, 1.000 | 0.010, .341 |
| Network-rest | 0.17 (0.14, 0.22) | 0.20 (0.17, 0.23) | 0.16 (0.14, 0.18) | −0.005, .521 | 0.001, .998 | 0.006, .160 |
| *Reward network* | | | | | | |
| Within-network | 0.39 (0.25, 0.46) | 0.28 (0.22, 0.34) | 0.39 (0.33, 0.43) | 0.016, .621 | 0.000, 1.000 | −0.016, .081 |
| Network-rest | 0.14 (0.12, 0.17) | 0.15 (0.13, 0.17) | 0.14 (0.12, 0.15) | −0.002, .675 | 0.001, .998 | 0.003, .348 |
| *Affective control network* | | | | | | |
| Within-network | 0.33 (0.25, 0.38) | 0.32 (0.28, 0.36) | 0.32 (0.28, 0.34) | 0.001, 1.000 | 0.001, 1.000 | 0.000, 1.000 |
| Network-rest | 0.14 (0.13, 0.17) | 0.16 (0.15, 0.17) | 0.14 (0.14, 0.15) | −0.003, .217 | 0.000, .998 | **0.003, .023** |

Means were calculated across a range of density thresholds ($K$ = 0.10–0.25, $\Delta K$ = 0.01) and are reported with 95% bootstrap confidence intervals (CIs; 5000 resamples). Group differences in area under the curve (ΔAUC) were assessed using permutation testing (10 000 permutations). Bold values indicate statistically significant group differences after Benjamini–Hochberg correction for multiple comparisons ($p_{BH}$ < .05). Abbreviations: SZ+A, schizophrenia with aggression; SZ−A, schizophrenia without aggression; HC, healthy controls.

#### Appendix 4
Exploratory subgroup analyses

**Table A4.1.** Group differences in global network topology by illness stage.

(a) Early-course schizophrenia

| | M (95% CI) | | | ΔAUC, $p_{BH}$ | | |
|---|---|---|---|---|---|---|
| Metric | SZ+A (*N* = 162) | SZ-A (*N* = 964) | HC (*N* = 3058) | SZ+A vs SZ-A | SZ+A vs HC | SZ-A vs HC |
| Transitivity | 0.45 (0.39, 0.51) | 0.56 (0.50, 0.59) | 0.50 (0.46, 0.52) | −0.017, .075 | −0.008, .856 | **0.009, .037** |
| Modularity | 0.37 (0.30, 0.42) | 0.28 (0.25, 0.32) | 0.39 (0.36, 0.42) | **0.014, .023** | −0.003, .856 | **−0.017, < .001** |
| Efficiency | 0.48 (0.44, 0.51) | 0.38 (0.35, 0.44) | 0.45 (0.44, 0.48) | 0.015, .051 | 0.004, .856 | **−0.011, .004** |
| Small-worldness | 1.49 (1.30, 1.80) | 1.13 (1.06, 1.27) | 1.47 (1.33, 1.65) | **0.054, .023** | 0.005, .856 | **−0.048, .019** |

(b) Chronic schizophrenia

| | M (95% CI) | | | ΔAUC, $p_{BH}$ | | |
|---|---|---|---|---|---|---|
| Metric | SZ+A (*N* = 60) | SZ-A (*N* = 892) | HC (*N* = 3058) | SZ+A vs SZ-A | SZ+A vs HC | SZ-A vs HC |
| Transitivity | 0.40 (0.37, 0.54) | 0.50 (0.45, 0.54) | 0.50 (0.46, 0.52) | −0.016, .246 | −0.015, .615 | 0.001, .931 |
| Modularity | 0.38 (0.27, 0.45) | 0.33 (0.28, 0.35) | 0.39 (0.36, 0.42) | 0.007, .359 | −0.003, .953 | −0.010, .147 |
| Efficiency | 0.50 (0.43, 0.51) | 0.45 (0.42, 0.48) | 0.45 (0.44, 0.48) | 0.008, .246 | 0.006, .615 | −0.001, .855 |
| Small-worldness | 1.57 (1.23, 2.06) | 1.39 (1.23, 1.51) | 1.47 (1.33, 1.65) | 0.027, .314 | 0.017, .615 | −0.010, .855 |

Means were calculated across a range of density thresholds ($K$ = 0.10–0.25, $\Delta K$ = 0.01) and are reported with 95% bootstrap confidence intervals (CIs; 5000 resamples). Group differences in area under the curve (ΔAUC) were assessed using permutation testing (10 000 permutations). Bold values indicate statistically significant group differences after Benjamini–Hochberg correction for multiple comparisons ($p_{BH} < .05$). Abbreviations: SZ+A, schizophrenia with aggression; SZ−A, schizophrenia without aggression; HC, healthy controls.

**Table A4.2.** Group differences in salience network-to-rest-of-brain density by delusion severity.

(a) Low-severity delusions

| M (95% CI) | | | ΔAUC, *p* | | |
|---|---|---|---|---|---|
| SZ+A (*N* = 31) | SZ-A (*N* = 1149) | HC (*N* = 3058) | SZ+A vs SZ-A | SZ+A vs HC | SZ-A vs HC |
| 0.18 (0.07, 0.26) | 0.19 (0.15, 0.24) | 0.14 (0.12, 0.18) | −0.002, .822 | 0.006, .580 | 0.008, .104 |

(b) High-severity delusions

| M (95% CI) | | | ΔAUC, *p* | | |
|---|---|---|---|---|---|
| SZ+A (*N* = 169) | SZ-A (*N* = 739) | HC (*N* = 3058) | SZ+A vs SZ-A | SZ+A vs HC | SZ-A vs HC |
| 0.28 (0.18, 0.34) | 0.20 (0.14, 0.28) | 0.14 (0.12, 0.18) | 0.012, .237 | **0.021, .007** | 0.009, .093 |

Means were calculated across a range of density thresholds ($K$ = 0.10–0.25, $\Delta K$ = 0.01) and are reported with 95% bootstrap confidence intervals (CIs; 5000 resamples). Group differences in area under the curve (ΔAUC) were assessed using permutation testing (10 000 permutations). Bold values indicate statistically significant group differences ($p < .05$). Abbreviations: SZ+A, schizophrenia with aggression; SZ−A, schizophrenia without aggression; HC, healthy controls.

#### Appendix 5

Sensitivity analyses

**Table A5.1.** Group differences in global network topology using a mild aggression cut-off.

| | M (95% CI) | | | ΔAUC, $p_{BH}$ | | |
|---|---|---|---|---|---|---|
| Metric | SZ+A (*N* = 429) | SZ-A (*N* = 1831) | HC (*N* = 3058) | SZ+A vs SZ-A | SZ+A vs HC | SZ-A vs HC |
| Transitivity | 0.50 (0.43, 0.54) | 0.56 (0.50, 0.58) | 0.50 (0.46, 0.52) | −0.009, .233 | −0.001, .944 | **0.008, .028** |
| Modularity | 0.35 (0.30, 0.39) | 0.30 (0.27, 0.33) | 0.39 (0.36, 0.42) | 0.008, .095 | −0.006, .944 | **−0.014, .001** |
| Efficiency | 0.46 (0.42, 0.50) | 0.39 (0.37, 0.44) | 0.45 (0.44, 0.48) | 0.011, .095 | 0.002, .944 | **−0.010, .002** |
| Small-worldness | 1.41 (1.26, 1.64) | 1.18 (1.12, 1.30) | 1.47 (1.33, 1.65) | 0.037, .095 | −0.005, .944 | **−0.041, .020** |

Means were calculated across a range of density thresholds ($K$ = 0.10–0.25, $\Delta K$ = 0.01) and are reported with 95% bootstrap confidence intervals (CIs; 5000 resamples). Group differences in area under the curve (ΔAUC) were assessed using permutation testing (10 000 permutations). Bold values indicate statistically significant group differences after Benjamini–Hochberg correction for multiple comparisons ($p_{BH} < .05$). Abbreviations: SZ+A, schizophrenia with aggression; SZ−A, schizophrenia without aggression; HC, healthy controls.

**Table A5.2.** Group differences in degree using a mild aggression cut-off.

| | M (95% CI) | | | ΔAUC, $p_{BH}$ | | |
|---|---|---|---|---|---|---|
| Node | SZ+A (*N* = 429) | SZ-A (*N* = 1831) | HC (*N* = 3058) | SZ+A vs SZ-A | SZ+A vs HC | SZ-A vs HC |
| FPOL | 1.1 (0.0, 2.8) | 0.0 (0.0, 0.6) | 1.0 (0.0, 2.1) | 0.170, .700 | 0.026, .914 | −0.145, .407 |
| POP | 3.4 (1.6, 8.1) | 3.8 (1.7, 7.4) | 3.9 (2.4, 6.4) | −0.051, .967 | −0.079, .914 | −0.029, .986 |
| PTRI | 5.4 (2.6, 8.1) | 4.3 (2.9, 6.2) | 5.1 (3.3, 6.1) | 0.170, .789 | 0.045, .914 | −0.125, .755 |
| PORB | 3.5 (2.1, 5.9) | 4.6 (3.2, 6.2) | 4.7 (3.4, 7.2) | −0.176, .753 | −0.180, .914 | −0.005, .986 |
| MOFC | 10.1 (5.4, 15.1) | 12.1 (8.7, 14.8) | 9.0 (6.5, 11.2) | −0.296, .789 | 0.166, .914 | 0.463, .445 |
| LOFC | 12.2 (9.0, 16.0) | 18.7 (14.3, 20.9) | 18.2 (13.4, 20.4) | −0.977, .700 | −0.918, .914 | 0.059, .986 |
| CMFG | 5.6 (3.0, 9.3) | 7.8 (5.5, 9.9) | 8.2 (6.8, 10.0) | −0.345, .753 | −0.384, .914 | −0.040, .986 |
| RMFG | 16.9 (11.2, 19.5) | 17.3 (12.8, 20.2) | 14.9 (10.5, 17.8) | −0.056, .967 | 0.313, .914 | 0.370, .755 |
| SFG | 15.8 (12.7, 18.4) | 20.9 (15.8, 23.8) | 17.9 (13.6, 21.3) | −0.760, .702 | −0.324, .914 | 0.436, .739 |
| PARL | 6.5 (3.0, 9.5) | 7.5 (4.9, 9.8) | 8.1 (5.4, 10.4) | −0.155, .914 | −0.231, .914 | −0.076, .969 |
| PREG | 8.4 (5.9, 13.4) | 11.1 (7.2, 15.2) | 9.3 (6.9, 12.4) | −0.409, .789 | −0.140, .914 | 0.269, .755 |
| CACC | 4.0 (2.3, 7.2) | 2.6 (2.0, 3.6) | 3.1 (2.1, 4.4) | 0.214, .702 | 0.135, .914 | −0.079, .739 |
| RACC | 8.1 (5.1, 11.4) | 11.6 (9.0, 13.4) | 8.5 (6.4, 11.2) | −0.530, .702 | −0.056, .914 | 0.474, .407 |
| PCC | 13.8 (8.6, 17.1) | 10.9 (7.2, 14.1) | 10.8 (7.9, 14.8) | 0.420, .753 | 0.436, .914 | 0.016, .986 |
| ISTH | 6.6 (2.9, 11.4) | 2.7 (1.2, 5.6) | 3.6 (2.6, 6.6) | 0.596, .702 | 0.456, .914 | −0.140, .755 |
| INS | 14.6 (9.4, 18.2) | 11.6 (8.0, 15.2) | 7.9 (6.2, 10.7) | 0.462, .753 | 1.010, .517 | 0.549, .353 |
| TPOL | 1.6 (0.0, 3.2) | 0.0 (0.0, 0.5) | 1.0 (0.9, 1.9) | 0.235, .700 | 0.085, .914 | −0.150, .054 |
| ENTH | 1.1 (0.0, 2.7) | 0.2 (0.0, 1.2) | 2.0 (1.9, 2.6) | 0.135, .702 | −0.130, .914 | **−0.265, .004** |
| PHG | 3.1 (1.3, 5.3) | 1.7 (1.0, 3.1) | 3.4 (2.9, 5.4) | 0.210, .702 | −0.049, .914 | −0.260, .055 |
| TVTG | 6.9 (3.6, 10.5) | 6.2 (3.9, 9.2) | 8.2 (5.6, 9.8) | 0.126, .914 | −0.179, .914 | −0.305, .594 |
| ITG | 9.2 (5.4, 13.3) | 6.4 (3.7, 8.8) | 8.1 (5.4, 11.1) | 0.440, .748 | 0.179, .914 | −0.260, .739 |
| MTG | 7.8 (5.3, 12.6) | 11.8 (8.5, 14.0) | 6.9 (5.4, 9.1) | −0.622, .702 | 0.114, .914 | 0.736, .054 |
| STG | 16.7 (12.1, 19.7) | 13.4 (10.6, 15.6) | 9.8 (7.8, 12.4) | 0.479, .702 | 1.040, .406 | 0.561, .198 |
| BANKS | 3.6 (2.1, 5.5) | 3.4 (3.0, 4.8) | 3.6 (3.0, 4.7) | 0.045, .914 | 0.015, .918 | −0.030, .969 |
| FUSI | 12.8 (7.1, 16.8) | 12.2 (8.8, 16.0) | 10.7 (6.8, 14.6) | 0.105, .967 | 0.320, .914 | 0.214, .937 |

| | | | | | | |
|---|---|---|---|---|---|---|
| POSG | 11.3 (6.9, 14.3) | 9.2 (5.8, 13.1) | 8.2 (5.9, 11.2) | 0.335, .789 | 0.460, .914 | 0.125, .969 |
| PREC | 13.7 (8.6, 17.3) | 17.2 (11.4, 20.4) | 18.9 (15.2, 21.0) | −0.552, .753 | −0.808, .914 | −0.256, .755 |
| SMG | 7.2 (3.4, 11.2) | 8.1 (5.2, 11.1) | 8.6 (6.2, 10.6) | −0.134, .914 | −0.201, .914 | −0.067, .969 |
| IPL | 7.3 (5.4, 10.9) | 9.7 (6.5, 12.5) | 9.2 (7.0, 11.7) | −0.349, .753 | −0.284, .914 | 0.065, .969 |
| SPL | 7.5 (4.0, 10.8) | 4.9 (2.7, 8.8) | 6.8 (4.2, 9.8) | 0.385, .753 | 0.123, .914 | −0.262, .755 |
| LING | 4.6 (2.5, 5.1) | 3.2 (2.9, 3.9) | 4.4 (4.0, 5.8) | 0.220, .700 | 0.045, .914 | −0.175, .054 |
| CUN | 4.6 (3.1, 6.9) | 3.0 (3.0, 3.9) | 5.1 (4.1, 6.0) | 0.235, .702 | −0.080, .914 | −0.315, .055 |
| PERI | 2.4 (2.0, 3.8) | 2.1 (2.0, 2.7) | 3.4 (2.9, 3.9) | 0.055, .748 | −0.154, .914 | **−0.210, .004** |
| LOC | 3.8 (2.2, 7.3) | 6.2 (4.6, 8.9) | 6.1 (4.4, 8.8) | −0.359, .702 | −0.340, .914 | 0.019, .986 |
| HIPPO | 3.1 (2.6, 6.6) | 3.1 (2.1, 5.3) | 4.4 (3.4, 5.5) | −0.010, .967 | −0.215, .914 | −0.205, .407 |
| AMYG | 5.2 (2.5, 9.5) | 3.0 (2.0, 5.4) | 4.2 (3.2, 6.2) | 0.324, .702 | 0.139, .914 | −0.185, .606 |
| CAUD | 2.9 (1.8, 3.8) | 2.8 (2.0, 3.6) | 3.6 (2.8, 4.4) | 0.015, .967 | −0.110, .914 | −0.125, .129 |
| PUT | 4.5 (3.2, 6.7) | 4.5 (3.3, 5.5) | 5.8 (4.9, 6.4) | −0.006, .967 | −0.201, .914 | −0.195, .407 |
| NACC | 3.4 (1.4, 6.2) | 1.8 (1.1, 2.8) | 4.4 (3.3, 5.2) | 0.251, .700 | −0.139, .914 | **−0.390, .004** |
| PALL | 2.9 (2.0, 4.7) | 2.6 (2.0, 3.0) | 2.0 (1.6, 2.0) | 0.060, .847 | 0.145, .914 | **0.085, .004** |
| THAL | 3.6 (1.4, 6.9) | 2.9 (1.6, 5.2) | 3.9 (2.4, 5.6) | 0.099, .914 | −0.051, .914 | −0.149, .755 |

Means were calculated across a range of density thresholds ($K$ = 0.10–0.25, $\Delta K$ = 0.01) and are reported with 95% bootstrap confidence intervals (CIs; 5000 resamples). Group differences in area under the curve (ΔAUC) were assessed using permutation testing (10 000 permutations). Bold values indicate statistically significant group differences after Benjamini–Hochberg correction for multiple comparisons ($p_{BH}$ < .05). Abbreviations: SZ+A, schizophrenia with aggression; SZ−A, schizophrenia without aggression; HC, healthy controls; FPOL, frontal pole; POP, pars opercularis; PTRI, pars triangularis; PORB, pars orbitalis; MOFC, medial orbitofrontal cortex; LOFC, lateral orbitofrontal cortex; CMFG, caudal middle frontal gyrus; RMFG, rostral middle frontal gyrus; SFG, superior frontal gyrus; PARL, paracentral lobule; PREG, precentral gyrus; CACC, caudal anterior cingulate cortex; RACC, rostral anterior cingulate cortex; PCC, posterior cingulate cortex; ISTH, isthmus of the cingulate cortex; INS, insular cortex; TPOL, temporal pole; ENTH, entorhinal cortex; PHG, parahippocampal gyrus; TVTG, transverse temporal gyrus; ITG, inferior temporal gyrus; MTG, middle temporal gyrus; STG, superior temporal gyrus; BANKS, banks of the superior temporal sulcus; FUSI, fusiform gyrus; POSG, postcentral gyrus; PREC, precuneus; SMG, supramarginal gyrus; IPL, inferior parietal lobule; SPL, superior parietal lobule; LING, lingual gyrus; CUN, cuneus; PERI, pericalcarine cortex; LOC, lateral occipital cortex; HIPPO, hippocampus; AMYG, amygdala; CAUD, caudate nucleus; PUT, putamen; NACC, nucleus accumbens; PALL, globus pallidus; THAL, thalamus.

**Table A5.3.** Group differences in betweenness centrality using a mild aggression cut-off.

| | M (95% CI) | | | ΔAUC, $p_{BH}$ | | |
|---|---|---|---|---|---|---|
| Node | SZ+A (*N* = 428) | SZ-A (*N* = 1831) | HC (*N* = 3058) | SZ+A vs SZ-A | SZ+A vs HC | SZ-A vs HC |
| FPOL | 0.000 (0.000, 0.000) | 0.000 (0.000, 0.000) | 0.000 (0.000, 0.000) | 0.00000, 1.000 | 0.00000, 1.000 | 0.00000, 1.000 |
| POP | 0.002 (0.000, 0.015) | 0.001 (0.000, 0.011) | 0.001 (0.000, 0.007) | 0.00002, 1.000 | 0.00012, .948 | 0.00010, .950 |
| PTRI | 0.009 (0.002, 0.035) | 0.008 (0.000, 0.028) | 0.004 (0.001, 0.022) | 0.00016, 1.000 | 0.00068, .948 | 0.00052, .959 |
| PORB | 0.001 (0.000, 0.029) | 0.015 (0.000, 0.036) | 0.002 (0.000, 0.027) | −0.00188, .803 | −0.00017, .948 | 0.00171, .553 |
| MOFC | 0.015 (0.001, 0.071) | 0.008 (0.003, 0.036) | 0.018 (0.002, 0.043) | 0.00105, .873 | −0.00052, .948 | −0.00157, .816 |
| LOFC | 0.039 (0.019, 0.128) | 0.065 (0.026, 0.136) | 0.117 (0.037, 0.192) | −0.00361, .873 | −0.01144, .948 | −0.00783, .685 |
| CMFG | 0.001 (0.000, 0.014) | 0.001 (0.000, 0.006) | 0.016 (0.002, 0.028) | −0.00000, 1.000 | −0.00205, .700 | −0.00205, .117 |
| RMFG | 0.100 (0.036, 0.170) | 0.071 (0.025, 0.140) | 0.069 (0.022, 0.131) | 0.00435, .873 | 0.00473, .948 | 0.00038, 1.000 |
| SFG | 0.069 (0.028, 0.130) | 0.198 (0.038, 0.311) | 0.081 (0.030, 0.210) | −0.01882, .770 | −0.00115, .958 | 0.01768, .540 |
| PARL | 0.002 (0.000, 0.011) | 0.001 (0.000, 0.004) | 0.004 (0.001, 0.012) | 0.00019, .873 | −0.00033, .948 | −0.00052, .318 |
| PREG | 0.004 (0.002, 0.044) | 0.007 (0.002, 0.076) | 0.011 (0.004, 0.068) | −0.00041, 1.000 | −0.00085, .948 | −0.00045, 1.000 |
| CACC | 0.000 (0.000, 0.007) | 0.000 (0.000, 0.000) | 0.000 (0.000, 0.000) | 0.00003, .803 | 0.00003, .948 | 0.00000, 1.000 |
| RACC | 0.007 (0.002, 0.067) | 0.017 (0.009, 0.032) | 0.011 (0.005, 0.040) | −0.00140, .873 | −0.00054, .948 | 0.00086, .817 |
| PCC | 0.051 (0.015, 0.146) | 0.014 (0.007, 0.037) | 0.036 (0.012, 0.119) | 0.00536, .671 | 0.00203, .948 | −0.00333, .540 |
| ISTH | 0.033 (0.002, 0.123) | 0.006 (0.000, 0.021) | 0.022 (0.007, 0.056) | 0.00431, .671 | 0.00206, .948 | −0.00226, .439 |
| INS | 0.086 (0.014, 0.205) | 0.011 (0.004, 0.116) | 0.005 (0.002, 0.052) | 0.01114, .731 | 0.01215, .575 | 0.00101, .685 |
| TPOL | 0.016 (0.000, 0.046) | 0.000 (0.000, 0.000) | 0.000 (0.000, 0.013) | 0.00249, .398 | 0.00249, .663 | 0.00000, 1.000 |
| ENTH | 0.001 (0.000, 0.034) | 0.000 (0.000, 0.018) | 0.049 (0.018, 0.050) | 0.00015, .873 | −0.00726, .563 | **−0.00740, .006** |
| PHG | 0.130 (0.000, 0.153) | 0.031 (0.000, 0.154) | 0.297 (0.131, 0.346) | 0.01597, .873 | −0.02427, .948 | **−0.04025, .041** |
| TVTG | 0.001 (0.000, 0.017) | 0.001 (0.000, 0.012) | 0.008 (0.002, 0.026) | −0.00002, 1.000 | −0.00104, .948 | −0.00102, .540 |
| ITG | 0.062 (0.007, 0.167) | 0.000 (0.000, 0.008) | 0.086 (0.006, 0.173) | 0.00957, .398 | −0.00306, .948 | −0.01263, .115 |
| MTG | 0.034 (0.006, 0.128) | 0.015 (0.008, 0.085) | 0.012 (0.004, 0.050) | 0.00258, .873 | 0.00314, .948 | 0.00055, .959 |
| STG | 0.103 (0.034, 0.194) | 0.024 (0.016, 0.071) | 0.037 (0.017, 0.088) | 0.01208, .573 | 0.01014, .575 | −0.00194, .766 |
| BANKS | 0.000 (0.000, 0.004) | 0.000 (0.000, 0.001) | 0.001 (0.000, 0.002) | 0.00002, .873 | −0.00005, .948 | −0.00008, .135 |
| FUSI | 0.274 (0.047, 0.312) | 0.090 (0.050, 0.249) | 0.322 (0.157, 0.382) | 0.02928, .731 | −0.00666, .956 | −0.03594, .115 |

| | | | | | | |
|---|---|---|---|---|---|---|
| POSG | 0.020 (0.005, 0.055) | 0.007 (0.002, 0.020) | 0.013 (0.003, 0.035) | 0.00202, .731 | 0.00098, .948 | −0.00103, .685 |
| PREC | 0.077 (0.024, 0.179) | 0.075 (0.036, 0.174) | 0.255 (0.128, 0.336) | −0.00029, 1.000 | −0.02648, .700 | −0.02620, .115 |
| SMG | 0.013 (0.001, 0.038) | 0.004 (0.001, 0.015) | 0.011 (0.004, 0.027) | 0.00141, .731 | 0.00039, .948 | −0.00102, .434 |
| IPL | 0.022 (0.007, 0.071) | 0.011 (0.005, 0.027) | 0.029 (0.013, 0.111) | 0.00149, .873 | −0.00114, .948 | −0.00263, .530 |
| SPL | 0.024 (0.002, 0.069) | 0.001 (0.000, 0.014) | 0.019 (0.003, 0.045) | 0.00350, .671 | 0.00121, .948 | −0.00230, .439 |
| LING | 0.026 (0.001, 0.054) | 0.020 (0.006, 0.026) | 0.010 (0.002, 0.056) | 0.00103, .873 | 0.00289, .948 | 0.00186, .439 |
| CUN | 0.022 (0.004, 0.079) | 0.018 (0.007, 0.032) | 0.020 (0.005, 0.039) | 0.00030, 1.000 | 0.00025, .956 | −0.00005, 1.000 |
| PERI | 0.000 (0.000, 0.003) | 0.000 (0.000, 0.000) | 0.000 (0.000, 0.001) | 0.00000, 1.000 | −0.00003, .948 | **−0.00003, .048** |
| LOC | 0.044 (0.005, 0.094) | 0.112 (0.075, 0.137) | 0.051 (0.019, 0.100) | −0.01026, .675 | −0.00163, .948 | 0.00863, .145 |
| HIPPO | 0.095 (0.003, 0.125) | 0.098 (0.001, 0.168) | 0.173 (0.054, 0.228) | −0.00014, 1.000 | −0.01123, .948 | −0.01108, .670 |
| AMYG | 0.072 (0.007, 0.191) | 0.027 (0.004, 0.139) | 0.104 (0.015, 0.176) | 0.00742, .873 | −0.00362, .948 | −0.01104, .540 |
| CAUD | 0.003 (0.000, 0.017) | 0.001 (0.000, 0.012) | 0.001 (0.001, 0.014) | 0.00032, .811 | 0.00025, .948 | −0.00007, 1.000 |
| PUT | 0.016 (0.009, 0.097) | 0.038 (0.009, 0.092) | 0.074 (0.042, 0.092) | −0.00355, .873 | −0.00883, .563 | −0.00528, .434 |
| NACC | 0.042 (0.000, 0.140) | 0.001 (0.000, 0.010) | 0.013 (0.003, 0.061) | 0.00657, .398 | 0.00470, .948 | −0.00187, .685 |
| PALL | 0.009 (0.000, 0.053) | 0.003 (0.000, 0.031) | 0.000 (0.000, 0.000) | 0.00082, .962 | 0.00135, .160 | **0.00054, .004** |
| THAL | 0.062 (0.000, 0.145) | 0.019 (0.000, 0.114) | 0.019 (0.002, 0.115) | 0.00684, .873 | 0.00670, .948 | −0.00014, 1.000 |

Means were calculated across a range of density thresholds ($K$ = 0.10–0.25, $\Delta K$ = 0.01) and are reported with 95% bootstrap confidence intervals (CIs; 5000 resamples). Group differences in area under the curve (ΔAUC) were assessed using permutation testing (10 000 permutations). Bold values indicate statistically significant group differences after Benjamini–Hochberg correction for multiple comparisons ($p_{BH} < .05$). Abbreviations: SZ+A, schizophrenia with aggression; SZ−A, schizophrenia without aggression; HC, healthy controls; FPOL, frontal pole; POP, pars opercularis; PTRI, pars triangularis; PORB, pars orbitalis; MOFC, medial orbitofrontal cortex; LOFC, lateral orbitofrontal cortex; CMFG, caudal middle frontal gyrus; RMFG, rostral middle frontal gyrus; SFG, superior frontal gyrus; PARL, paracentral lobule; PREG, precentral gyrus; CACC, caudal anterior cingulate cortex; RACC, rostral anterior cingulate cortex; PCC, posterior cingulate cortex; ISTH, isthmus of the cingulate cortex; INS, insular cortex; TPOL, temporal pole; ENTH, entorhinal cortex; PHG, parahippocampal gyrus; TVTG, transverse temporal gyrus; ITG, inferior temporal gyrus; MTG, middle temporal gyrus; STG, superior temporal gyrus; BANKS, banks of the superior temporal sulcus; FUSI, fusiform gyrus; POSG, postcentral gyrus; PREC, precuneus; SMG, supramarginal gyrus; IPL, inferior parietal lobule; SPL, superior parietal lobule; LING, lingual gyrus; CUN, cuneus; PERI, pericalcarine cortex; LOC, lateral occipital cortex; HIPPO, hippocampus; AMYG, amygdala; CAUD, caudate nucleus; PUT, putamen; NACC, nucleus accumbens; PALL, globus pallidus; THAL, thalamus.

**Table A5.4.** Group differences in harmonic centrality using a mild aggression cut-off.

| | M (95% CI) | | | ΔAUC, $p_{BH}$ | | |
|---|---|---|---|---|---|---|
| Node | SZ+A (*N* = 429) | SZ-A (*N* = 1831) | HC (*N* = 3058) | SZ+A vs SZ-A | SZ+A vs HC | SZ-A vs HC |
| FPOL | 0.268 (0.000, 0.433) | 0.000 (0.000, 0.210) | 0.204 (0.000, 0.377) | 0.041, .458 | 0.010, .908 | −0.030, .359 |
| POP | 0.409 (0.317, 0.507) | 0.385 (0.305, 0.475) | 0.412 (0.359, 0.472) | 0.004, .938 | −0.000, .997 | −0.004, .781 |
| PTRI | 0.460 (0.360, 0.513) | 0.397 (0.348, 0.457) | 0.441 (0.393, 0.469) | 0.010, .483 | 0.003, .908 | −0.007, .306 |
| PORB | 0.422 (0.378, 0.475) | 0.420 (0.395, 0.467) | 0.445 (0.423, 0.496) | 0.000, .974 | −0.003, .908 | −0.004, .431 |
| MOFC | 0.558 (0.472, 0.630) | 0.530 (0.492, 0.583) | 0.520 (0.480, 0.562) | 0.005, .738 | 0.006, .908 | 0.001, .844 |
| LOFC | 0.588 (0.531, 0.646) | 0.621 (0.571, 0.675) | 0.655 (0.590, 0.696) | −0.005, .738 | −0.010, .908 | −0.006, .486 |
| CMFG | 0.468 (0.415, 0.529) | 0.467 (0.440, 0.509) | 0.495 (0.479, 0.538) | 0.000, .974 | −0.004, .908 | −0.004, .359 |
| RMFG | 0.649 (0.567, 0.690) | 0.609 (0.554, 0.665) | 0.602 (0.539, 0.654) | 0.006, .722 | 0.007, .908 | 0.001, .900 |
| SFG | 0.623 (0.577, 0.672) | 0.662 (0.592, 0.730) | 0.648 (0.592, 0.711) | −0.005, .738 | −0.003, .908 | 0.002, .844 |
| PARL | 0.478 (0.406, 0.538) | 0.458 (0.424, 0.511) | 0.508 (0.465, 0.547) | 0.003, .824 | −0.004, .908 | −0.008, .254 |
| PREG | 0.511 (0.461, 0.593) | 0.518 (0.467, 0.596) | 0.527 (0.492, 0.580) | −0.001, .974 | −0.002, .908 | −0.002, .844 |
| CACC | 0.435 (0.390, 0.508) | 0.363 (0.337, 0.407) | 0.406 (0.370, 0.440) | 0.011, .458 | 0.004, .908 | −0.007, .207 |
| RACC | 0.522 (0.457, 0.582) | 0.522 (0.489, 0.565) | 0.508 (0.475, 0.565) | 0.000, .974 | 0.003, .908 | 0.002, .790 |
| PCC | 0.608 (0.528, 0.662) | 0.511 (0.466, 0.570) | 0.553 (0.505, 0.622) | 0.015, .458 | 0.008, .908 | −0.007, .471 |
| ISTH | 0.489 (0.405, 0.581) | 0.387 (0.347, 0.455) | 0.448 (0.425, 0.498) | 0.016, .458 | 0.007, .908 | −0.009, .128 |
| INS | 0.620 (0.538, 0.684) | 0.523 (0.479, 0.601) | 0.505 (0.482, 0.560) | 0.015, .458 | 0.018, .683 | 0.003, .768 |
| TPOL | 0.281 (0.000, 0.424) | 0.000 (0.000, 0.113) | 0.245 (0.226, 0.352) | 0.043, .458 | 0.006, .908 | **−0.037, .031** |
| ENTH | 0.208 (0.000, 0.406) | 0.076 (0.000, 0.265) | 0.320 (0.300, 0.377) | 0.021, .722 | −0.016, .908 | **−0.037, .031** |
| PHG | 0.412 (0.232, 0.475) | 0.353 (0.300, 0.413) | 0.434 (0.405, 0.481) | 0.010, .483 | −0.002, .908 | **−0.012, .033** |
| TVTG | 0.493 (0.438, 0.555) | 0.446 (0.408, 0.512) | 0.517 (0.470, 0.547) | 0.008, .603 | −0.003, .908 | −0.011, .117 |
| ITG | 0.543 (0.462, 0.616) | 0.452 (0.404, 0.506) | 0.529 (0.471, 0.589) | 0.014, .458 | 0.002, .908 | −0.012, .117 |
| MTG | 0.526 (0.462, 0.608) | 0.526 (0.485, 0.580) | 0.493 (0.457, 0.541) | −0.000, .994 | 0.005, .908 | 0.005, .486 |
| STG | 0.651 (0.581, 0.702) | 0.549 (0.521, 0.601) | 0.538 (0.506, 0.585) | 0.016, .458 | 0.017, .656 | 0.002, .792 |
| BANKS | 0.431 (0.367, 0.484) | 0.370 (0.361, 0.430) | 0.408 (0.390, 0.448) | 0.010, .483 | 0.004, .908 | −0.006, .245 |
| FUSI | 0.612 (0.498, 0.676) | 0.547 (0.500, 0.623) | 0.574 (0.516, 0.643) | 0.011, .483 | 0.006, .908 | −0.005, .728 |

| | | | | | | |
|---|---|---|---|---|---|---|
| POSG | 0.557 (0.484, 0.610) | 0.487 (0.444, 0.557) | 0.520 (0.478, 0.567) | 0.011, .483 | 0.006, .908 | −0.005, .486 |
| PREC | 0.615 (0.530, 0.668) | 0.608 (0.540, 0.673) | 0.676 (0.632, 0.716) | 0.001, .974 | −0.009, .908 | −0.010, .175 |
| SMG | 0.502 (0.412, 0.560) | 0.465 (0.428, 0.530) | 0.520 (0.484, 0.557) | 0.006, .727 | −0.002, .908 | −0.008, .198 |
| IPL | 0.516 (0.467, 0.579) | 0.499 (0.456, 0.558) | 0.538 (0.507, 0.585) | 0.003, .872 | −0.003, .908 | −0.006, .339 |
| SPL | 0.500 (0.428, 0.567) | 0.424 (0.385, 0.500) | 0.490 (0.447, 0.545) | 0.012, .483 | 0.002, .908 | −0.010, .245 |
| LING | 0.409 (0.288, 0.451) | 0.333 (0.311, 0.373) | 0.401 (0.372, 0.467) | 0.012, .458 | 0.002, .908 | −0.010, .093 |
| CUN | 0.405 (0.309, 0.487) | 0.329 (0.311, 0.395) | 0.446 (0.391, 0.480) | 0.012, .483 | −0.006, .908 | **−0.018, .033** |
| PERI | 0.326 (0.255, 0.396) | 0.258 (0.242, 0.327) | 0.362 (0.338, 0.392) | 0.011, .483 | −0.005, .908 | **−0.016, .029** |
| LOC | 0.437 (0.326, 0.514) | 0.464 (0.421, 0.525) | 0.478 (0.417, 0.541) | −0.004, .840 | −0.006, .908 | −0.003, .781 |
| HIPPO | 0.360 (0.269, 0.497) | 0.255 (0.139, 0.449) | 0.377 (0.335, 0.453) | 0.017, .722 | −0.002, .920 | −0.019, .135 |
| AMYG | 0.400 (0.281, 0.553) | 0.222 (0.142, 0.427) | 0.366 (0.310, 0.468) | 0.028, .483 | 0.006, .908 | −0.022, .117 |
| CAUD | 0.292 (0.208, 0.382) | 0.180 (0.121, 0.290) | 0.278 (0.254, 0.347) | 0.018, .483 | 0.003, .908 | −0.015, .117 |
| PUT | 0.344 (0.250, 0.477) | 0.229 (0.146, 0.354) | 0.346 (0.309, 0.406) | 0.018, .483 | 0.000, .997 | −0.018, .117 |
| NACC | 0.365 (0.217, 0.495) | 0.170 (0.109, 0.287) | 0.323 (0.286, 0.400) | 0.031, .458 | 0.007, .908 | **−0.023, .033** |
| PALL | 0.307 (0.212, 0.397) | 0.182 (0.120, 0.298) | 0.258 (0.230, 0.291) | 0.020, .483 | 0.008, .908 | −0.012, .117 |
| THAL | 0.380 (0.227, 0.514) | 0.209 (0.132, 0.410) | 0.328 (0.290, 0.439) | 0.027, .483 | 0.009, .908 | −0.018, .245 |

Means were calculated across a range of density thresholds ($K$ = 0.10–0.25, $\Delta K$ = 0.01) and are reported with 95% bootstrap confidence intervals (CIs; 5000 resamples). Group differences in area under the curve (ΔAUC) were assessed using permutation testing (10 000 permutations). Bold values indicate statistically significant group differences after Benjamini–Hochberg correction for multiple comparisons ($p_{BH} < .05$). Abbreviations: SZ+A, schizophrenia with aggression; SZ−A, schizophrenia without aggression; HC, healthy controls; FPOL, frontal pole; POP, pars opercularis; PTRI, pars triangularis; PORB, pars orbitalis; MOFC, medial orbitofrontal cortex; LOFC, lateral orbitofrontal cortex; CMFG, caudal middle frontal gyrus; RMFG, rostral middle frontal gyrus; SFG, superior frontal gyrus; PARL, paracentral lobule; PREG, precentral gyrus; CACC, caudal anterior cingulate cortex; RACC, rostral anterior cingulate cortex; PCC, posterior cingulate cortex; ISTH, isthmus of the cingulate cortex; INS, insular cortex; TPOL, temporal pole; ENTH, entorhinal cortex; PHG, parahippocampal gyrus; TVTG, transverse temporal gyrus; ITG, inferior temporal gyrus; MTG, middle temporal gyrus; STG, superior temporal gyrus; BANKS, banks of the superior temporal sulcus; FUSI, fusiform gyrus; POSG, postcentral gyrus; PREC, precuneus; SMG, supramarginal gyrus; IPL, inferior parietal lobule; SPL, superior parietal lobule; LING, lingual gyrus; CUN, cuneus; PERI, pericalcarine cortex; LOC, lateral occipital cortex; HIPPO, hippocampus; AMYG, amygdala; CAUD, caudate nucleus; PUT, putamen; NACC, nucleus accumbens; PALL, globus pallidus; THAL, thalamus.

**Table A5.5.** Group differences in within-module degree using a mild aggression cut-off.

| | M (95% CI) | | | ΔAUC, $p_{BH}$ | | |
|---|---|---|---|---|---|---|
| Node | SZ+A (*N* = 429) | SZ-A (*N* = 1831) | HC (*N* = 3058) | SZ+A vs SZ-A | SZ+A vs HC | SZ-A vs HC |
| FPOL | −1.66 (−1.87, 0.00) | 0.00 (−1.56, 0.00) | −1.76 (−1.87, 0.00) | −0.248, .988 | 0.015, .934 | 0.263, .320 |
| POP | −1.14 (−1.53, 0.00) | −1.05 (−1.42, −0.11) | −1.07 (−1.46, −0.50) | −0.013, 1.000 | −0.012, .934 | 0.001, .993 |
| PTRI | −0.69 (−1.24, 1.08) | −0.92 (−1.21, 0.36) | −0.78 (−1.24, −0.50) | 0.037, .988 | 0.016, .934 | −0.022, .986 |
| PORB | −1.10 (−1.48, −0.44) | −0.85 (−1.18, −0.48) | −0.87 (−1.20, −0.35) | −0.039, .988 | −0.037, .934 | 0.002, .993 |
| MOFC | 0.33 (−0.71, 1.49) | 0.53 (−0.13, 1.09) | 0.08 (−0.50, 0.62) | −0.032, .989 | 0.035, .934 | 0.067, .876 |
| LOFC | 0.78 (0.01, 1.60) | 1.83 (0.99, 2.28) | 2.10 (1.13, 2.68) | −0.158, .988 | −0.201, .791 | −0.043, .986 |
| CMFG | −0.65 (−1.23, 0.17) | −0.23 (−0.73, 0.15) | −0.02 (−0.36, 0.37) | −0.067, .988 | −0.094, .791 | −0.027, .986 |
| RMFG | 1.87 (0.49, 2.36) | 1.42 (0.61, 2.01) | 1.25 (0.38, 1.90) | 0.069, .988 | 0.095, .934 | 0.026, .986 |
| SFG | 1.64 (1.00, 2.32) | 2.05 (1.27, 2.58) | 2.22 (1.31, 2.85) | −0.060, .988 | −0.085, .934 | −0.025, .986 |
| PARL | −0.48 (−1.26, 0.26) | −0.34 (−0.83, 0.13) | −0.11 (−0.67, 0.46) | −0.023, .989 | −0.055, .934 | −0.032, .986 |
| PREG | −0.04 (−0.56, 1.07) | 0.36 (−0.38, 1.13) | 0.18 (−0.36, 0.85) | −0.061, .988 | −0.035, .934 | 0.026, .986 |
| CACC | −1.02 (−1.48, −0.28) | −1.24 (−1.51, −1.05) | −1.24 (−1.53, −0.93) | 0.033, .988 | 0.033, .934 | 0.000, .993 |
| RACC | −0.08 (−0.84, 0.53) | 0.51 (−0.03, 0.88) | −0.02 (−0.49, 0.62) | −0.087, .988 | −0.008, .934 | 0.079, .655 |
| PCC | 1.13 (−0.06, 1.77) | 0.33 (−0.39, 0.96) | 0.56 (−0.13, 1.33) | 0.118, .988 | 0.086, .934 | −0.032, .986 |
| ISTH | −0.84 (−1.42, 0.48) | −1.35 (−1.62, −0.89) | −1.38 (−1.63, −0.52) | 0.079, .988 | 0.088, .934 | 0.009, .986 |
| INS | 1.21 (0.12, 1.94) | 0.40 (−0.28, 1.08) | −0.11 (−0.52, 0.48) | 0.122, .988 | 0.200, .454 | 0.078, .655 |
| TPOL | 0.00 (−1.74, 0.00) | 0.00 (0.00, 0.00) | −0.58 (−0.61, 0.00) | 0.000, 1.000 | 0.087, .934 | 0.087, .651 |
| ENTH | 0.00 (−1.83, 1.05) | 0.00 (−1.53, 0.00) | 1.15 (0.00, 1.15) | 0.000, 1.000 | −0.173, .934 | −0.173, .395 |
| PHG | −1.56 (−1.81, 0.87) | −1.53 (−1.73, 0.00) | −0.58 (−1.04, −0.43) | −0.010, 1.000 | −0.154, .710 | −0.143, .395 |
| TVTG | −0.37 (−1.10, 0.43) | −0.57 (−1.06, −0.03) | −0.11 (−0.71, 0.27) | 0.031, .989 | −0.038, .934 | −0.070, .655 |
| ITG | −0.14 (−0.81, 0.97) | −0.53 (−1.11, −0.10) | −0.20 (−0.77, 0.37) | 0.060, .988 | 0.010, .934 | −0.050, .972 |
| MTG | −0.38 (−0.79, 0.80) | 0.53 (−0.13, 0.92) | −0.36 (−0.71, 0.12) | −0.141, .988 | −0.007, .934 | 0.134, .141 |
| STG | 1.85 (0.73, 2.37) | 0.85 (0.33, 1.25) | 0.31 (−0.14, 0.88) | 0.148, .988 | 0.232, .217 | 0.085, .502 |
| BANKS | −1.08 (−1.47, −0.21) | −1.07 (−1.28, −0.84) | −1.12 (−1.33, −0.85) | 0.000, 1.000 | 0.006, .934 | 0.006, .986 |
| FUSI | 0.27 (−0.94, 1.26) | 0.41 (−0.44, 1.02) | −0.01 (−0.77, 0.77) | −0.014, 1.000 | 0.046, .934 | 0.060, .986 |

| | | | | | | |
|---|---|---|---|---|---|---|
| POSG | 0.64 (−0.35, 1.29) | −0.03 (−0.66, 0.73) | −0.10 (−0.62, 0.54) | 0.103, .988 | 0.111, .934 | 0.008, .986 |
| PREC | 0.94 (−0.34, 1.69) | 1.43 (0.36, 2.00) | 1.78 (1.11, 2.51) | −0.075, .988 | −0.127, .934 | −0.052, .986 |
| SMG | −0.31 (−1.18, 0.60) | −0.22 (−0.78, 0.39) | −0.01 (−0.51, 0.48) | −0.014, 1.000 | −0.046, .934 | −0.032, .986 |
| IPL | −0.24 (−0.68, 0.84) | 0.10 (−0.53, 0.65) | 0.14 (−0.34, 0.68) | −0.052, .988 | −0.059, .934 | −0.007, .986 |
| SPL | −0.45 (−1.23, 0.32) | −0.82 (−1.30, −0.17) | −0.73 (−1.30, −0.01) | 0.054, .988 | 0.043, .934 | −0.011, .986 |
| LING | 0.60 (−0.89, 1.00) | 0.81 (0.00, 1.14) | 0.92 (−0.74, 1.10) | −0.031, .988 | −0.047, .934 | −0.016, .986 |
| CUN | 0.67 (−0.57, 1.22) | 0.81 (0.00, 0.87) | −0.20 (−0.63, 0.38) | −0.025, .989 | 0.134, .710 | 0.159, .050 |
| PERI | −0.60 (−1.28, 0.35) | −0.81 (−0.87, 0.00) | 0.40 (−1.07, 0.65) | 0.031, .988 | −0.159, .934 | −0.189, .050 |
| LOC | −0.67 (−1.28, 0.24) | −0.81 (−0.89, 0.44) | 0.26 (−0.40, 0.62) | 0.025, .989 | −0.141, .710 | −0.166, .126 |
| HIPPO | −0.12 (−1.17, 1.36) | 0.14 (−1.03, 1.24) | −0.31 (−0.80, 0.37) | −0.036, 1.000 | 0.027, .934 | 0.063, .986 |
| AMYG | 1.11 (−0.84, 1.58) | −0.07 (−0.58, 0.76) | −0.18 (−0.45, 0.46) | 0.177, .988 | 0.198, .219 | 0.021, .986 |
| CAUD | −0.25 (−1.00, 0.87) | 0.54 (−0.42, 0.81) | −0.13 (−0.48, 0.36) | −0.117, .988 | −0.012, .934 | 0.105, .502 |
| PUT | 1.36 (0.46, 1.90) | 1.00 (0.89, 1.94) | 1.59 (1.27, 1.84) | 0.049, .988 | −0.042, .934 | −0.091, .502 |
| NACC | −0.25 (−1.30, 0.45) | −1.00 (−1.37, −0.33) | 0.46 (−0.05, 0.81) | 0.115, .988 | −0.103, .934 | −0.218, .050 |
| PALL | −0.18 (−0.87, 0.93) | −0.54 (−0.84, 0.28) | −1.45 (−1.73, −1.03) | 0.058, .988 | 0.196, .217 | 0.138, .050 |
| THAL | −0.11 (−1.38, 0.65) | −0.07 (−0.71, 0.76) | 0.02 (−0.82, 0.45) | −0.005, 1.000 | −0.023, .934 | −0.018, .986 |

Means were calculated across a range of density thresholds ($K$ = 0.10–0.25, $\Delta K$ = 0.01) and are reported with 95% bootstrap confidence intervals (CIs; 5000 resamples). Group differences in area under the curve (ΔAUC) were assessed using permutation testing (10 000 permutations). Abbreviations: SZ+A, schizophrenia with aggression; SZ−A, schizophrenia without aggression; HC, healthy controls; FPOL, frontal pole; POP, pars opercularis; PTRI, pars triangularis; PORB, pars orbitalis; MOFC, medial orbitofrontal cortex; LOFC, lateral orbitofrontal cortex; CMFG, caudal middle frontal gyrus; RMFG, rostral middle frontal gyrus; SFG, superior frontal gyrus; PARL, paracentral lobule; PREG, precentral gyrus; CACC, caudal anterior cingulate cortex; RACC, rostral anterior cingulate cortex; PCC, posterior cingulate cortex; ISTH, isthmus of the cingulate cortex; INS, insular cortex; TPOL, temporal pole; ENTH, entorhinal cortex; PHG, parahippocampal gyrus; TVTG, transverse temporal gyrus; ITG, inferior temporal gyrus; MTG, middle temporal gyrus; STG, superior temporal gyrus; BANKS, banks of the superior temporal sulcus; FUSI, fusiform gyrus; POSG, postcentral gyrus; PREC, precuneus; SMG, supramarginal gyrus; IPL, inferior parietal lobule; SPL, superior parietal lobule; LING, lingual gyrus; CUN, cuneus; PERI, pericalcarine cortex; LOC, lateral occipital cortex; HIPPO, hippocampus; AMYG, amygdala; CAUD, caudate nucleus; PUT, putamen; NACC, nucleus accumbens; PALL, globus pallidus; THAL, thalamus.

**Table A5.6.** Group differences in participation coefficient using a mild aggression cut-off.

| | M (95% CI) | | | ΔAUC, $p_{BH}$ | | |
|---|---|---|---|---|---|---|
| Node | SZ+A (*N* = 429) | SZ-A (*N* = 1831) | HC (*N* = 3058) | SZ+A vs SZ-A | SZ+A vs HC | SZ-A vs HC |
| FPOL | 0.00 (0.00, 0.00) | 0.00 (0.00, 0.00) | 0.00 (0.00, 0.00) | 0.000, 1.000 | 0.000, 1.000 | 0.000, 1.000 |
| POP | 0.00 (0.00, 0.31) | 0.00 (0.00, 0.18) | 0.00 (0.00, 0.00) | 0.000, 1.000 | 0.000, 1.000 | 0.000, 1.000 |
| PTRI | 0.00 (0.00, 0.32) | 0.00 (0.00, 0.36) | 0.00 (0.00, 0.00) | 0.000, 1.000 | 0.000, 1.000 | 0.000, 1.000 |
| PORB | 0.00 (0.00, 0.48) | 0.00 (0.00, 0.41) | 0.02 (0.00, 0.16) | 0.000, 1.000 | −0.001, 1.000 | −0.001, .748 |
| MOFC | 0.00 (0.00, 0.18) | 0.00 (0.00, 0.05) | 0.01 (0.00, 0.11) | 0.000, 1.000 | −0.001, 1.000 | −0.001, 1.000 |
| LOFC | 0.00 (0.00, 0.28) | 0.02 (0.00, 0.15) | 0.06 (0.00, 0.11) | −0.002, .896 | −0.009, 1.000 | −0.007, .974 |
| CMFG | 0.00 (0.00, 0.08) | 0.00 (0.00, 0.00) | 0.00 (0.00, 0.07) | 0.000, 1.000 | 0.000, 1.000 | 0.000, 1.000 |
| RMFG | 0.00 (0.00, 0.29) | 0.08 (0.00, 0.19) | 0.09 (0.00, 0.18) | −0.013, .640 | −0.013, 1.000 | −0.000, 1.000 |
| SFG | 0.00 (0.00, 0.10) | 0.12 (0.01, 0.21) | 0.00 (0.00, 0.10) | −0.018, .582 | 0.000, 1.000 | 0.018, .336 |
| PARL | 0.00 (0.00, 0.04) | 0.00 (0.00, 0.00) | 0.00 (0.00, 0.00) | 0.000, 1.000 | 0.000, 1.000 | 0.000, 1.000 |
| PREG | 0.00 (0.00, 0.09) | 0.00 (0.00, 0.10) | 0.00 (0.00, 0.08) | 0.000, 1.000 | 0.000, 1.000 | 0.000, 1.000 |
| CACC | 0.00 (0.00, 0.16) | 0.00 (0.00, 0.00) | 0.00 (0.00, 0.00) | 0.000, 1.000 | 0.000, 1.000 | 0.000, 1.000 |
| RACC | 0.00 (0.00, 0.25) | 0.00 (0.00, 0.01) | 0.02 (0.00, 0.12) | 0.000, 1.000 | −0.002, 1.000 | −0.002, .291 |
| PCC | 0.01 (0.00, 0.27) | 0.00 (0.00, 0.02) | 0.00 (0.00, 0.10) | 0.000, .727 | 0.000, 1.000 | 0.000, 1.000 |
| ISTH | 0.31 (0.00, 0.55) | 0.15 (0.00, 0.41) | 0.49 (0.00, 0.50) | 0.027, .896 | −0.025, 1.000 | −0.051, .218 |
| INS | 0.05 (0.00, 0.28) | 0.00 (0.00, 0.14) | 0.00 (0.00, 0.09) | 0.008, .896 | 0.008, 1.000 | 0.000, 1.000 |
| TPOL | 0.30 (0.00, 0.50) | 0.00 (0.00, 0.00) | 0.00 (0.00, 0.38) | 0.046, .465 | 0.046, 1.000 | 0.000, 1.000 |
| ENTH | 0.25 (0.00, 0.50) | 0.00 (0.00, 0.22) | 0.00 (0.00, 0.50) | 0.037, .582 | 0.037, 1.000 | 0.000, 1.000 |
| PHG | 0.52 (0.00, 0.63) | 0.26 (0.00, 0.54) | 0.66 (0.60, 0.72) | 0.043, .727 | −0.017, 1.000 | **−0.060, .004** |
| TVTG | 0.00 (0.00, 0.07) | 0.00 (0.00, 0.11) | 0.00 (0.00, 0.02) | 0.000, 1.000 | 0.000, 1.000 | 0.000, 1.000 |
| ITG | 0.17 (0.00, 0.55) | 0.00 (0.00, 0.02) | 0.05 (0.00, 0.24) | 0.025, .582 | 0.019, 1.000 | −0.007, .589 |
| MTG | 0.19 (0.00, 0.61) | 0.00 (0.00, 0.08) | 0.00 (0.00, 0.10) | 0.029, .465 | 0.029, 1.000 | 0.000, 1.000 |
| STG | 0.02 (0.00, 0.30) | 0.00 (0.00, 0.08) | 0.00 (0.00, 0.08) | 0.002, .896 | 0.002, 1.000 | 0.000, 1.000 |
| BANKS | 0.00 (0.00, 0.32) | 0.00 (0.00, 0.00) | 0.00 (0.00, 0.00) | 0.000, 1.000 | 0.000, 1.000 | 0.000, 1.000 |
| FUSI | 0.37 (0.12, 0.66) | 0.16 (0.01, 0.41) | 0.33 (0.22, 0.45) | 0.032, .597 | 0.005, 1.000 | −0.026, .136 |

| | | | | | | |
|---|---|---|---|---|---|---|
| POSG | 0.00 (0.00, 0.12) | 0.00 (0.00, 0.02) | 0.00 (0.00, 0.11) | 0.000, 1.000 | 0.000, 1.000 | 0.000, 1.000 |
| PREC | 0.08 (0.00, 0.46) | 0.08 (0.00, 0.22) | 0.26 (0.00, 0.33) | −0.002, 1.000 | −0.028, 1.000 | −0.026, .223 |
| SMG | 0.00 (0.00, 0.23) | 0.00 (0.00, 0.00) | 0.00 (0.00, 0.05) | 0.000, 1.000 | 0.000, 1.000 | 0.000, 1.000 |
| IPL | 0.00 (0.00, 0.51) | 0.00 (0.00, 0.05) | 0.00 (0.00, 0.14) | 0.000, 1.000 | 0.000, 1.000 | 0.000, 1.000 |
| SPL | 0.18 (0.00, 0.44) | 0.00 (0.00, 0.16) | 0.32 (0.00, 0.43) | 0.028, .586 | −0.021, 1.000 | −0.049, .223 |
| LING | 0.41 (0.00, 0.49) | 0.07 (0.00, 0.46) | 0.11 (0.00, 0.52) | 0.053, .582 | 0.047, 1.000 | −0.006, 1.000 |
| CUN | 0.35 (0.00, 0.49) | 0.00 (0.00, 0.46) | 0.46 (0.00, 0.48) | 0.054, .641 | −0.016, 1.000 | **−0.070, .018** |
| PERI | 0.07 (0.00, 0.30) | 0.00 (0.00, 0.03) | 0.00 (0.00, 0.19) | 0.009, .582 | 0.009, 1.000 | 0.000, 1.000 |
| LOC | 0.47 (0.00, 0.54) | 0.44 (0.38, 0.50) | 0.43 (0.00, 0.51) | 0.005, .896 | 0.003, 1.000 | −0.002, 1.000 |
| HIPPO | 0.02 (0.00, 0.59) | 0.26 (0.00, 0.49) | 0.35 (0.03, 0.48) | −0.036, .793 | −0.051, 1.000 | −0.015, .971 |
| AMYG | 0.16 (0.00, 0.62) | 0.41 (0.00, 0.53) | 0.18 (0.00, 0.42) | −0.039, .896 | −0.003, 1.000 | 0.037, .748 |
| CAUD | 0.00 (0.00, 0.17) | 0.07 (0.00, 0.14) | 0.00 (0.00, 0.00) | −0.009, .582 | 0.000, 1.000 | 0.009, .223 |
| PUT | 0.00 (0.00, 0.47) | 0.37 (0.00, 0.40) | 0.00 (0.00, 0.11) | −0.056, .597 | 0.000, 1.000 | 0.056, .218 |
| NACC | 0.21 (0.00, 0.58) | 0.08 (0.00, 0.19) | 0.00 (0.00, 0.15) | 0.021, .641 | 0.032, 1.000 | 0.011, .480 |
| PALL | 0.00 (0.00, 0.44) | 0.25 (0.00, 0.31) | 0.00 (0.00, 0.00) | −0.038, .641 | 0.000, 1.000 | **0.038, .004** |
| THAL | 0.16 (0.00, 0.58) | 0.26 (0.00, 0.41) | 0.00 (0.00, 0.26) | −0.015, 1.000 | 0.025, 1.000 | 0.040, .291 |

Means were calculated across a range of density thresholds ($K$ = 0.10–0.25, $\Delta K$ = 0.01) and are reported with 95% bootstrap confidence intervals (CIs; 5000 resamples). Group differences in area under the curve (ΔAUC) were assessed using permutation testing (10 000 permutations). Bold values indicate statistically significant group differences after Benjamini–Hochberg correction for multiple comparisons ($p_{BH} < .05$). Abbreviations: SZ+A, schizophrenia with aggression; SZ−A, schizophrenia without aggression; HC, healthy controls; FPOL, frontal pole; POP, pars opercularis; PTRI, pars triangularis; PORB, pars orbitalis; MOFC, medial orbitofrontal cortex; LOFC, lateral orbitofrontal cortex; CMFG, caudal middle frontal gyrus; RMFG, rostral middle frontal gyrus; SFG, superior frontal gyrus; PARL, paracentral lobule; PREG, precentral gyrus; CACC, caudal anterior cingulate cortex; RACC, rostral anterior cingulate cortex; PCC, posterior cingulate cortex; ISTH, isthmus of the cingulate cortex; INS, insular cortex; TPOL, temporal pole; ENTH, entorhinal cortex; PHG, parahippocampal gyrus; TVTG, transverse temporal gyrus; ITG, inferior temporal gyrus; MTG, middle temporal gyrus; STG, superior temporal gyrus; BANKS, banks of the superior temporal sulcus; FUSI, fusiform gyrus; POSG, postcentral gyrus; PREC, precuneus; SMG, supramarginal gyrus; IPL, inferior parietal lobule; SPL, superior parietal lobule; LING, lingual gyrus; CUN, cuneus; PERI, pericalcarine cortex; LOC, lateral occipital cortex; HIPPO, hippocampus; AMYG, amygdala; CAUD, caudate nucleus; PUT, putamen; NACC, nucleus accumbens; PALL, globus pallidus; THAL, thalamus.

**Table A5.7.** Group differences in canonical brain network density using a mild aggression cut-off.

| | M (95% CI) | | | ΔAUC, $p_{BH}$ | | |
|---|---|---|---|---|---|---|
| Metric | SZ+A (*N* = 429) | SZ-A (*N* = 1831) | HC (*N* = 3058) | SZ+A vs SZ-A | SZ+A vs HC | SZ-A vs HC |
| *Salience network* | | | | | | |
| Network-rest | 0.24 (0.17, 0.29) | 0.18 (0.14, 0.23) | 0.14 (0.12, 0.18) | 0.009, .453 | 0.015, .119 | 0.006, .260 |
| *Central executive network* | | | | | | |
| Within-network | 0.49 (0.20, 0.65) | 0.46 (0.30, 0.59) | 0.29 (0.17, 0.51) | 0.005, 1.000 | 0.031, .756 | 0.026, .427 |
| Network-rest | 0.21 (0.17, 0.25) | 0.23 (0.19, 0.26) | 0.24 (0.21, 0.26) | −0.003, .838 | −0.004, .665 | −0.001, .792 |
| *Default mode network* | | | | | | |
| Within-network | 0.16 (0.13, 0.22) | 0.15 (0.12, 0.17) | 0.18 (0.15, 0.20) | 0.001, 1.000 | −0.003, .756 | −0.004, .427 |
| Network-rest | 0.15 (0.13, 0.16) | 0.15 (0.14, 0.16) | 0.15 (0.14, 0.16) | −0.000, .959 | 0.000, .899 | 0.000, .792 |
| *Language network* | | | | | | |
| Within-network | 0.32 (0.30, 0.49) | 0.37 (0.30, 0.42) | 0.31 (0.30, 0.38) | −0.008, 1.000 | 0.001, 1.000 | 0.009, .427 |
| Network-rest | 0.19 (0.16, 0.23) | 0.20 (0.17, 0.23) | 0.16 (0.14, 0.18) | −0.001, .953 | 0.005, .665 | 0.006, .190 |
| *Reward network* | | | | | | |
| Within-network | 0.32 (0.22, 0.39) | 0.29 (0.22, 0.35) | 0.39 (0.33, 0.43) | 0.005, 1.000 | −0.010, .756 | −0.015, .187 |
| Network-rest | 0.12 (0.10, 0.15) | 0.16 (0.14, 0.17) | 0.14 (0.12, 0.15) | −0.005, .263 | −0.002, .665 | 0.003, .260 |
| *Affective control network* | | | | | | |
| Within-network | 0.29 (0.24, 0.35) | 0.32 (0.28, 0.36) | 0.32 (0.28, 0.34) | −0.005, 1.000 | −0.005, .756 | 0.001, 1.000 |
| Network-rest | 0.15 (0.14, 0.17) | 0.17 (0.15, 0.17) | 0.14 (0.14, 0.15) | −0.002, .263 | 0.001, .768 | **0.003, .016** |

Means were calculated across a range of density thresholds (*K* = 0.10–0.25, Δ*K* = 0.01) and are reported with 95% bootstrap confidence intervals (CIs; 5000 resamples). Group differences in area under the curve (ΔAUC) were assessed using permutation testing (10 000 permutations). Bold values indicate statistically significant group differences after Benjamini–Hochberg correction for multiple comparisons ($p_{BH}$ < .05). Abbreviations: SZ+A, schizophrenia with aggression; SZ−A, schizophrenia without aggression; HC, healthy controls.

**Table A5.8.** Group differences in global network topology using PANSS P7 'Hostility' scores only

| Metric | M (95% CI) | | | ΔAUC, $p_{BH}$ | | |
|---|---|---|---|---|---|---|
| | SZ+A (*N* = 97) | SZ-A (*N* = 1477) | HC (*N* = 3058) | SZ+A vs SZ-A | SZ+A vs HC | SZ-A vs HC |
| Transitivity | 0.39 (0.36, 0.47) | 0.57 (0.50, 0.59) | 0.50 (0.46, 0.52) | **−0.026, .019** | −0.016, .358 | **0.010, .014** |
| Modularity | 0.37 (0.30, 0.43) | 0.31 (0.27, 0.33) | 0.39 (0.36, 0.42) | 0.009, .148 | −0.004, .926 | **−0.013, < .001** |
| Efficiency | 0.51 (0.46, 0.52) | 0.37 (0.36, 0.44) | 0.45 (0.44, 0.48) | **0.021, .004** | 0.008, .358 | **−0.013, < .001** |
| Small-worldness | 1.52 (1.30, 1.94) | 1.16 (1.13, 1.32) | 1.47 (1.33, 1.65) | **0.053, .025** | 0.009, .926 | **−0.044, .014** |

Means were calculated across a range of density thresholds ($K$ = 0.10–0.25, $\Delta K$ = 0.01) and are reported with 95% bootstrap confidence intervals (CIs; 5000 resamples). Group differences in area under the curve (ΔAUC) were assessed using permutation testing (10 000 permutations). Bold values indicate statistically significant group differences after Benjamini–Hochberg correction for multiple comparisons ($p_{BH}$ < .05). Abbreviations: SZ+A, schizophrenia with aggression; SZ−A, schizophrenia without aggression; HC, healthy controls.

**Table A5.9.** Group differences in degree using PANSS P7 'Hostility' scores only

| | M (95% CI) | | | ΔAUC, $p_{BH}$ | | |
|---|---|---|---|---|---|---|
| Node | SZ+A (*N* = 97) | SZ-A (*N* = 1477) | HC (*N* = 3058) | SZ+A vs SZ-A | SZ+A vs HC | SZ-A vs HC |
| FPOL | 2.4 (0.0, 9.5) | 0.0 (0.0, 0.5) | 1.0 (0.0, 2.1) | 0.370, .506 | 0.225, .783 | −0.145, .508 |
| POP | 6.1 (3.6, 11.8) | 2.6 (1.1, 6.3) | 3.9 (2.4, 6.4) | 0.515, .583 | 0.311, .783 | −0.204, .790 |
| PTRI | 5.9 (3.0, 11.6) | 3.8 (2.2, 5.6) | 5.1 (3.3, 6.1) | 0.320, .583 | 0.116, .838 | −0.204, .650 |
| PORB | 5.9 (2.2, 9.6) | 4.2 (2.8, 5.7) | 4.7 (3.4, 7.2) | 0.261, .717 | 0.190, .821 | −0.071, .914 |
| MOFC | 11.5 (4.9, 15.5) | 12.0 (8.4, 15.2) | 9.0 (6.5, 11.2) | −0.080, .937 | 0.376, .783 | 0.455, .570 |
| LOFC | 10.1 (6.6, 15.0) | 18.4 (13.9, 21.1) | 18.2 (13.4, 20.4) | −1.264, .506 | −1.249, .783 | 0.015, .997 |
| CMFG | 8.8 (3.8, 13.3) | 8.4 (5.9, 10.5) | 8.2 (6.8, 10.0) | 0.068, .937 | 0.114, .847 | 0.046, .968 |
| RMFG | 12.2 (5.2, 15.0) | 18.0 (13.1, 20.9) | 14.9 (10.5, 17.8) | −0.870, .583 | −0.395, .802 | 0.474, .650 |
| SFG | 16.7 (9.4, 18.4) | 19.5 (14.7, 23.1) | 17.9 (13.6, 21.3) | −0.409, .583 | −0.188, .826 | 0.221, .908 |
| PARL | 5.1 (1.5, 10.9) | 8.5 (5.6, 10.2) | 8.1 (5.4, 10.4) | −0.514, .583 | −0.435, .783 | 0.079, .963 |
| PREG | 5.6 (2.0, 12.9) | 11.8 (7.6, 15.7) | 9.3 (6.9, 12.4) | −0.931, .583 | −0.556, .783 | 0.375, .650 |
| CACC | 3.8 (1.0, 6.5) | 2.5 (2.0, 3.6) | 3.1 (2.1, 4.4) | 0.195, .611 | 0.105, .821 | −0.090, .650 |
| RACC | 4.4 (2.6, 10.0) | 11.1 (8.4, 13.2) | 8.5 (6.4, 11.2) | −1.011, .506 | −0.610, .783 | 0.401, .596 |
| PCC | 7.4 (2.6, 13.5) | 12.1 (9.1, 14.9) | 10.8 (7.9, 14.8) | −0.740, .583 | −0.529, .783 | 0.211, .893 |
| ISTH | 6.2 (1.4, 11.3) | 4.8 (2.2, 8.3) | 3.6 (2.6, 6.6) | 0.220, .879 | 0.406, .783 | 0.186, .790 |
| INS | 19.5 (9.9, 21.1) | 12.2 (8.7, 15.9) | 7.9 (6.2, 10.7) | 1.113, .506 | 1.744, .115 | 0.630, .339 |
| TPOL | 3.8 (0.6, 9.7) | 0.0 (0.0, 0.7) | 1.0 (0.9, 1.9) | 0.554, .506 | 0.404, .783 | −0.150, .085 |
| ENTH | 2.6 (0.8, 6.4) | 0.2 (0.0, 1.2) | 2.0 (1.9, 2.6) | 0.360, .506 | 0.085, .821 | **−0.275, .008** |
| PHG | 5.2 (2.1, 10.2) | 1.2 (0.4, 2.6) | 3.4 (2.9, 5.4) | 0.595, .506 | 0.260, .783 | **−0.335, .024** |
| TVTG | 4.9 (2.5, 10.5) | 5.2 (3.2, 8.6) | 8.2 (5.6, 9.8) | −0.054, .937 | −0.494, .783 | −0.440, .450 |
| ITG | 8.5 (3.3, 13.4) | 5.6 (3.5, 8.4) | 8.1 (5.4, 11.1) | 0.450, .583 | 0.065, .921 | −0.385, .596 |
| MTG | 4.0 (2.2, 9.1) | 10.0 (7.2, 12.8) | 6.9 (5.4, 9.1) | −0.905, .506 | −0.440, .783 | 0.466, .339 |
| STG | 8.2 (2.8, 15.9) | 12.6 (9.2, 15.1) | 9.8 (7.8, 12.4) | −0.665, .583 | −0.234, .821 | 0.431, .508 |
| BANKS | 5.6 (3.0, 9.6) | 3.2 (3.0, 4.8) | 3.6 (3.0, 4.7) | 0.349, .583 | 0.294, .783 | −0.055, .893 |
| FUSI | 13.3 (5.6, 18.3) | 12.2 (8.1, 15.7) | 10.7 (6.8, 14.6) | 0.180, .937 | 0.399, .783 | 0.219, .908 |

| | | | | | | |
|---|---|---|---|---|---|---|
| POSG | 11.6 (5.9, 16.8) | 8.0 (5.1, 11.9) | 8.2 (5.9, 11.2) | 0.577, .583 | 0.527, .783 | −0.050, .989 |
| PREC | 6.4 (2.8, 11.4) | 18.6 (12.5, 21.6) | 18.9 (15.2, 21.0) | −1.861, .506 | −1.901, .745 | −0.040, .996 |
| SMG | 11.3 (5.3, 15.2) | 5.3 (2.2, 9.1) | 8.6 (6.2, 10.6) | 0.901, .506 | 0.404, .783 | −0.497, .339 |
| IPL | 4.8 (2.0, 10.1) | 10.5 (7.7, 13.2) | 9.2 (7.0, 11.7) | −0.845, .506 | −0.659, .783 | 0.186, .801 |
| SPL | 5.3 (2.7, 9.8) | 6.1 (2.9, 10.7) | 6.8 (4.2, 9.8) | −0.114, .937 | −0.206, .821 | −0.092, .963 |
| LING | 3.6 (2.0, 11.6) | 3.3 (2.9, 4.1) | 4.4 (4.0, 5.8) | 0.050, .937 | −0.105, .821 | −0.155, .210 |
| CUN | 3.3 (2.0, 7.2) | 4.1 (3.0, 5.9) | 5.1 (4.1, 6.0) | −0.115, .879 | −0.265, .783 | −0.151, .650 |
| PERI | 2.7 (2.0, 6.8) | 2.1 (2.0, 2.7) | 3.4 (2.9, 3.9) | 0.095, .583 | −0.115, .783 | **−0.210, .005** |
| LOC | 3.0 (1.1, 8.1) | 6.2 (4.3, 9.0) | 6.1 (4.4, 8.8) | −0.486, .583 | −0.467, .783 | 0.019, .996 |
| HIPPO | 10.6 (4.5, 14.2) | 2.8 (2.1, 5.1) | 4.4 (3.4, 5.5) | 1.195, .506 | 0.930, .745 | −0.265, .339 |
| AMYG | 6.9 (2.0, 12.4) | 3.1 (2.2, 5.6) | 4.2 (3.2, 6.2) | 0.570, .583 | 0.399, .783 | −0.171, .650 |
| CAUD | 2.4 (0.0, 5.2) | 3.2 (2.2, 3.9) | 3.6 (2.8, 4.4) | −0.124, .855 | −0.190, .783 | −0.066, .674 |
| PUT | 7.8 (3.0, 12.3) | 5.0 (3.6, 6.0) | 5.8 (4.9, 6.4) | 0.410, .583 | 0.290, .783 | −0.120, .650 |
| NACC | 9.9 (3.8, 14.3) | 1.8 (1.0, 2.3) | 4.4 (3.3, 5.2) | 1.224, .246 | 0.829, .745 | **−0.395, .005** |
| PALL | 3.1 (1.3, 7.5) | 3.0 (2.4, 3.2) | 2.0 (1.6, 2.0) | 0.005, .937 | 0.155, .783 | **0.150, .004** |
| THAL | 6.7 (3.1, 12.0) | 3.9 (2.5, 6.7) | 3.9 (2.4, 5.6) | 0.410, .583 | 0.410, .783 | −0.001, .997 |

Means were calculated across a range of density thresholds ($K$ = 0.10–0.25, $\Delta K$ = 0.01) and are reported with 95% bootstrap confidence intervals (CIs; 5000 resamples). Group differences in area under the curve (ΔAUC) were assessed using permutation testing (10 000 permutations). Bold values indicate statistically significant group differences after Benjamini–Hochberg correction for multiple comparisons ($p_{BH} < .05$). Abbreviations: SZ+A, schizophrenia with aggression; SZ−A, schizophrenia without aggression; HC, healthy controls; FPOL, frontal pole; POP, pars opercularis; PTRI, pars triangularis; PORB, pars orbitalis; MOFC, medial orbitofrontal cortex; LOFC, lateral orbitofrontal cortex; CMFG, caudal middle frontal gyrus; RMFG, rostral middle frontal gyrus; SFG, superior frontal gyrus; PARL, paracentral lobule; PREG, precentral gyrus; CACC, caudal anterior cingulate cortex; RACC, rostral anterior cingulate cortex; PCC, posterior cingulate cortex; ISTH, isthmus of the cingulate cortex; INS, insular cortex; TPOL, temporal pole; ENTH, entorhinal cortex; PHG, parahippocampal gyrus; TVTG, transverse temporal gyrus; ITG, inferior temporal gyrus; MTG, middle temporal gyrus; STG, superior temporal gyrus; BANKS, banks of the superior temporal sulcus; FUSI, fusiform gyrus; POSG, postcentral gyrus; PREC, precuneus; SMG, supramarginal gyrus; IPL, inferior parietal lobule; SPL, superior parietal lobule; LING, lingual gyrus; CUN, cuneus; PERI, pericalcarine cortex; LOC, lateral occipital cortex; HIPPO, hippocampus; AMYG, amygdala; CAUD, caudate nucleus; PUT, putamen; NACC, nucleus accumbens; PALL, globus pallidus; THAL, thalamus.

**Table A5.10.** Group differences in betweenness centrality using PANSS P7 'Hostility' scores only

| | M (95% CI) | | | ΔAUC, $p_{BH}$ | | |
|---|---|---|---|---|---|---|
| Node | SZ+A (*N* = 97) | SZ-A (*N* = 1477) | HC (*N* = 3058) | SZ+A vs SZ-A | SZ+A vs HC | SZ-A vs HC |
| FPOL | 0.000 (0.000, 0.039) | 0.000 (0.000, 0.000) | 0.000 (0.000, 0.000) | 0.00001, .523 | 0.00001, .910 | 0.00000, 1.000 |
| POP | 0.008 (0.001, 0.064) | 0.001 (0.000, 0.011) | 0.001 (0.000, 0.007) | 0.00098, .815 | 0.00101, .910 | 0.00003, 1.000 |
| PTRI | 0.003 (0.000, 0.055) | 0.023 (0.001, 0.038) | 0.004 (0.001, 0.022) | −0.00313, .732 | −0.00017, .955 | 0.00296, .414 |
| PORB | 0.001 (0.000, 0.042) | 0.031 (0.001, 0.055) | 0.002 (0.000, 0.027) | −0.00436, .523 | −0.00010, .965 | 0.00427, .278 |
| MOFC | 0.031 (0.004, 0.113) | 0.009 (0.002, 0.038) | 0.018 (0.002, 0.043) | 0.00329, .815 | 0.00191, .910 | −0.00138, .797 |
| LOFC | 0.026 (0.016, 0.135) | 0.065 (0.029, 0.160) | 0.117 (0.037, 0.192) | −0.00575, .815 | −0.01355, .820 | −0.00781, .757 |
| CMFG | 0.030 (0.002, 0.087) | 0.001 (0.000, 0.008) | 0.016 (0.002, 0.028) | 0.00435, .523 | 0.00230, .820 | −0.00205, .347 |
| RMFG | 0.051 (0.004, 0.106) | 0.079 (0.025, 0.161) | 0.069 (0.022, 0.131) | −0.00414, .865 | −0.00251, .910 | 0.00163, 1.000 |
| SFG | 0.095 (0.032, 0.159) | 0.106 (0.019, 0.255) | 0.081 (0.030, 0.210) | −0.00059, .982 | 0.00266, .910 | 0.00326, 1.000 |
| PARL | 0.001 (0.000, 0.070) | 0.001 (0.000, 0.005) | 0.004 (0.001, 0.012) | 0.00001, .993 | −0.00042, .910 | −0.00043, .531 |
| PREG | 0.026 (0.000, 0.093) | 0.007 (0.002, 0.111) | 0.011 (0.004, 0.068) | 0.00298, .815 | 0.00259, .910 | −0.00039, 1.000 |
| CACC | 0.001 (0.000, 0.020) | 0.000 (0.000, 0.000) | 0.000 (0.000, 0.000) | 0.00018, .815 | 0.00018, .910 | 0.00000, 1.000 |
| RACC | 0.004 (0.002, 0.075) | 0.014 (0.008, 0.028) | 0.011 (0.005, 0.040) | −0.00163, .815 | −0.00109, .910 | 0.00054, .985 |
| PCC | 0.020 (0.002, 0.091) | 0.018 (0.010, 0.054) | 0.036 (0.012, 0.119) | 0.00023, .982 | −0.00254, .910 | −0.00277, .754 |
| ISTH | 0.044 (0.000, 0.126) | 0.007 (0.000, 0.034) | 0.022 (0.007, 0.056) | 0.00569, .523 | 0.00371, .820 | −0.00198, .575 |
| INS | 0.207 (0.052, 0.258) | 0.016 (0.005, 0.157) | 0.005 (0.002, 0.052) | 0.02849, .295 | 0.03022, .102 | 0.00173, .586 |
| TPOL | 0.008 (0.000, 0.079) | 0.000 (0.000, 0.002) | 0.000 (0.000, 0.013) | 0.00120, .523 | 0.00120, .910 | 0.00000, 1.000 |
| ENTH | 0.004 (0.000, 0.038) | 0.000 (0.000, 0.018) | 0.049 (0.018, 0.050) | 0.00042, .815 | −0.00698, .820 | **−0.00740, .004** |
| PHG | 0.026 (0.001, 0.077) | 0.009 (0.000, 0.091) | 0.297 (0.131, 0.346) | 0.00240, .815 | −0.04126, .910 | −0.04366, .078 |
| TVTG | 0.005 (0.001, 0.049) | 0.000 (0.000, 0.010) | 0.008 (0.002, 0.026) | 0.00063, .815 | −0.00054, .910 | −0.00117, .491 |
| ITG | 0.041 (0.003, 0.111) | 0.000 (0.000, 0.017) | 0.086 (0.006, 0.173) | 0.00625, .523 | −0.00638, .910 | −0.01263, .262 |
| MTG | 0.004 (0.000, 0.060) | 0.012 (0.006, 0.062) | 0.012 (0.004, 0.050) | −0.00128, .865 | −0.00122, .910 | 0.00006, 1.000 |
| STG | 0.016 (0.001, 0.139) | 0.024 (0.014, 0.063) | 0.037 (0.017, 0.088) | −0.00135, .886 | −0.00319, .910 | −0.00184, .811 |
| BANKS | 0.017 (0.004, 0.065) | 0.000 (0.000, 0.000) | 0.001 (0.000, 0.002) | 0.00222, .523 | 0.00213, .820 | −0.00009, .262 |
| FUSI | 0.179 (0.025, 0.194) | 0.076 (0.029, 0.197) | 0.322 (0.157, 0.382) | 0.01603, .523 | −0.02193, .965 | −0.03796, .262 |

| | | | | | | |
|---|---|---|---|---|---|---|
| POSG | 0.064 (0.010, 0.153) | 0.004 (0.001, 0.019) | 0.013 (0.003, 0.035) | 0.00928, .523 | 0.00791, .820 | −0.00137, .576 |
| PREC | 0.006 (0.001, 0.064) | 0.097 (0.032, 0.221) | 0.255 (0.128, 0.336) | −0.01398, .523 | −0.03659, .820 | −0.02261, .262 |
| SMG | 0.055 (0.005, 0.113) | 0.002 (0.000, 0.017) | 0.011 (0.004, 0.027) | 0.00790, .478 | 0.00656, .820 | −0.00134, .262 |
| IPL | 0.008 (0.000, 0.061) | 0.017 (0.008, 0.052) | 0.029 (0.013, 0.111) | −0.00127, .865 | −0.00311, .910 | −0.00183, .785 |
| SPL | 0.008 (0.001, 0.061) | 0.006 (0.000, 0.049) | 0.019 (0.003, 0.045) | 0.00031, .982 | −0.00122, .910 | −0.00153, .725 |
| LING | 0.016 (0.000, 0.108) | 0.011 (0.002, 0.023) | 0.010 (0.002, 0.056) | 0.00084, .886 | 0.00133, .910 | 0.00049, .860 |
| CUN | 0.014 (0.000, 0.078) | 0.027 (0.011, 0.050) | 0.020 (0.005, 0.039) | −0.00193, .865 | −0.00053, .934 | 0.00140, .725 |
| PERI | 0.000 (0.000, 0.039) | 0.000 (0.000, 0.000) | 0.000 (0.000, 0.001) | 0.00006, .815 | 0.00003, .910 | −0.00003, .262 |
| LOC | 0.037 (0.000, 0.090) | 0.066 (0.029, 0.116) | 0.051 (0.019, 0.100) | −0.00404, .875 | −0.00247, .910 | 0.00157, .977 |
| HIPPO | 0.056 (0.006, 0.129) | 0.029 (0.000, 0.126) | 0.173 (0.054, 0.228) | 0.00430, .815 | −0.01783, .910 | −0.02213, .350 |
| AMYG | 0.017 (0.000, 0.095) | 0.015 (0.001, 0.128) | 0.104 (0.015, 0.176) | 0.00086, .904 | −0.01203, .910 | −0.01290, .540 |
| CAUD | 0.002 (0.000, 0.023) | 0.001 (0.000, 0.017) | 0.001 (0.001, 0.014) | 0.00011, .982 | 0.00009, .955 | −0.00003, 1.000 |
| PUT | 0.057 (0.008, 0.110) | 0.023 (0.010, 0.087) | 0.074 (0.042, 0.092) | 0.00564, .732 | −0.00240, .910 | −0.00804, .346 |
| NACC | 0.070 (0.007, 0.129) | 0.000 (0.000, 0.002) | 0.013 (0.003, 0.061) | 0.01047, .478 | 0.00855, .820 | −0.00192, .540 |
| PALL | 0.007 (0.000, 0.043) | 0.002 (0.000, 0.034) | 0.000 (0.000, 0.000) | 0.00063, .815 | 0.00089, .820 | **0.00026, .018** |
| THAL | 0.039 (0.003, 0.100) | 0.010 (0.005, 0.149) | 0.019 (0.002, 0.115) | 0.00428, .815 | 0.00277, .910 | −0.00151, 1.000 |

Means were calculated across a range of density thresholds ($K$ = 0.10–0.25, $\Delta K$ = 0.01) and are reported with 95% bootstrap confidence intervals (CIs; 5000 resamples). Group differences in area under the curve (ΔAUC) were assessed using permutation testing (10 000 permutations). Bold values indicate statistically significant group differences after Benjamini–Hochberg correction for multiple comparisons ($p_{BH}$ < .05). Abbreviations: SZ+A, schizophrenia with aggression; SZ−A, schizophrenia without aggression; HC, healthy controls; FPOL, frontal pole; POP, pars opercularis; PTRI, pars triangularis; PORB, pars orbitalis; MOFC, medial orbitofrontal cortex; LOFC, lateral orbitofrontal cortex; CMFG, caudal middle frontal gyrus; RMFG, rostral middle frontal gyrus; SFG, superior frontal gyrus; PARL, paracentral lobule; PREG, precentral gyrus; CACC, caudal anterior cingulate cortex; RACC, rostral anterior cingulate cortex; PCC, posterior cingulate cortex; ISTH, isthmus of the cingulate cortex; INS, insular cortex; TPOL, temporal pole; ENTH, entorhinal cortex; PHG, parahippocampal gyrus; TVTG, transverse temporal gyrus; ITG, inferior temporal gyrus; MTG, middle temporal gyrus; STG, superior temporal gyrus; BANKS, banks of the superior temporal sulcus; FUSI, fusiform gyrus; POSG, postcentral gyrus; PREC, precuneus; SMG, supramarginal gyrus; IPL, inferior parietal lobule; SPL, superior parietal lobule; LING, lingual gyrus; CUN, cuneus; PERI, pericalcarine cortex; LOC, lateral occipital cortex; HIPPO, hippocampus; AMYG, amygdala; CAUD, caudate nucleus; PUT, putamen; NACC, nucleus accumbens; PALL, globus pallidus; THAL, thalamus.

**Table A5.11.** Group differences in harmonic centrality using PANSS P7 'Hostility' scores only

| | M (95% CI) | | | ΔAUC, $p_{BH}$ | | |
|---|---|---|---|---|---|---|
| Node | SZ+A (*N* = 97) | SZ-A (*N* = 1477) | HC (*N* = 3058) | SZ+A vs SZ-A | SZ+A vs HC | SZ-A vs HC |
| FPOL | 0.405 (0.000, 0.561) | 0.000 (0.000, 0.184) | 0.204 (0.000, 0.377) | 0.062, .366 | 0.032, .824 | −0.030, .365 |
| POP | 0.498 (0.427, 0.592) | 0.302 (0.259, 0.449) | 0.412 (0.359, 0.472) | 0.030, .404 | 0.013, .824 | −0.017, .133 |
| PTRI | 0.505 (0.415, 0.592) | 0.360 (0.314, 0.447) | 0.441 (0.393, 0.469) | 0.022, .366 | 0.010, .824 | −0.012, .088 |
| PORB | 0.509 (0.391, 0.563) | 0.401 (0.381, 0.452) | 0.445 (0.423, 0.496) | 0.016, .404 | 0.010, .842 | −0.007, .175 |
| MOFC | 0.580 (0.460, 0.656) | 0.514 (0.485, 0.585) | 0.520 (0.480, 0.562) | 0.010, .545 | 0.009, .824 | −0.001, .890 |
| LOFC | 0.577 (0.508, 0.650) | 0.604 (0.563, 0.678) | 0.655 (0.590, 0.696) | −0.004, .849 | −0.012, .816 | −0.008, .333 |
| CMFG | 0.568 (0.450, 0.625) | 0.457 (0.441, 0.517) | 0.495 (0.479, 0.538) | 0.017, .366 | 0.011, .816 | −0.006, .218 |
| RMFG | 0.615 (0.467, 0.648) | 0.603 (0.555, 0.674) | 0.602 (0.539, 0.654) | 0.002, .882 | 0.002, .899 | 0.000, .996 |
| SFG | 0.679 (0.560, 0.696) | 0.620 (0.570, 0.715) | 0.648 (0.592, 0.711) | 0.009, .545 | 0.005, .845 | −0.004, .754 |
| PARL | 0.500 (0.382, 0.595) | 0.458 (0.431, 0.517) | 0.508 (0.465, 0.547) | 0.006, .729 | −0.001, .899 | −0.008, .251 |
| PREG | 0.488 (0.380, 0.613) | 0.511 (0.470, 0.603) | 0.527 (0.492, 0.580) | −0.003, .898 | −0.006, .845 | −0.002, .810 |
| CACC | 0.438 (0.293, 0.503) | 0.350 (0.335, 0.408) | 0.406 (0.370, 0.440) | 0.014, .534 | 0.005, .845 | −0.008, .080 |
| RACC | 0.455 (0.386, 0.573) | 0.501 (0.480, 0.562) | 0.508 (0.475, 0.565) | −0.007, .729 | −0.008, .842 | −0.001, .890 |
| PCC | 0.543 (0.367, 0.628) | 0.516 (0.487, 0.582) | 0.553 (0.505, 0.622) | 0.004, .849 | −0.002, .899 | −0.006, .503 |
| ISTH | 0.516 (0.264, 0.598) | 0.412 (0.377, 0.493) | 0.448 (0.425, 0.498) | 0.016, .575 | 0.011, .842 | −0.005, .444 |
| INS | 0.726 (0.569, 0.737) | 0.520 (0.484, 0.613) | 0.505 (0.482, 0.560) | 0.031, .119 | 0.033, .529 | 0.002, .810 |
| TPOL | 0.437 (0.138, 0.577) | 0.000 (0.000, 0.137) | 0.245 (0.226, 0.352) | 0.066, .366 | 0.029, .816 | **−0.037, .025** |
| ENTH | 0.391 (0.165, 0.507) | 0.055 (0.000, 0.248) | 0.320 (0.300, 0.377) | 0.052, .366 | 0.011, .845 | **−0.041, .025** |
| PHG | 0.480 (0.348, 0.576) | 0.305 (0.093, 0.392) | 0.434 (0.405, 0.481) | 0.025, .445 | 0.007, .845 | **−0.018, .031** |
| TVTG | 0.491 (0.402, 0.582) | 0.419 (0.390, 0.498) | 0.517 (0.470, 0.547) | 0.011, .729 | −0.004, .847 | **−0.015, .031** |
| ITG | 0.552 (0.412, 0.629) | 0.418 (0.388, 0.498) | 0.529 (0.471, 0.589) | 0.021, .366 | 0.003, .896 | **−0.017, .031** |
| MTG | 0.446 (0.363, 0.553) | 0.487 (0.458, 0.556) | 0.493 (0.457, 0.541) | −0.006, .849 | −0.007, .845 | −0.001, .890 |
| STG | 0.552 (0.407, 0.665) | 0.523 (0.497, 0.590) | 0.538 (0.506, 0.585) | 0.004, .849 | 0.002, .899 | −0.002, .766 |
| BANKS | 0.487 (0.396, 0.562) | 0.354 (0.355, 0.427) | 0.408 (0.390, 0.448) | 0.020, .381 | 0.012, .816 | −0.008, .098 |
| FUSI | 0.647 (0.482, 0.696) | 0.528 (0.481, 0.614) | 0.574 (0.516, 0.643) | 0.018, .366 | 0.011, .824 | −0.007, .503 |

| | | | | | | |
|---|---|---|---|---|---|---|
| POSG | 0.595 (0.487, 0.675) | 0.456 (0.427, 0.539) | 0.520 (0.478, 0.567) | 0.022, .366 | 0.012, .816 | −0.010, .175 |
| PREC | 0.526 (0.392, 0.595) | 0.611 (0.547, 0.691) | 0.676 (0.632, 0.716) | −0.013, .418 | −0.023, .816 | −0.010, .161 |
| SMG | 0.599 (0.476, 0.650) | 0.395 (0.278, 0.496) | 0.520 (0.484, 0.557) | 0.030, .366 | 0.012, .816 | **−0.018, .008** |
| IPL | 0.473 (0.352, 0.575) | 0.499 (0.471, 0.570) | 0.538 (0.507, 0.585) | −0.003, .871 | −0.010, .824 | −0.006, .303 |
| SPL | 0.505 (0.393, 0.570) | 0.432 (0.391, 0.531) | 0.490 (0.447, 0.545) | 0.011, .729 | 0.003, .899 | −0.009, .317 |
| LING | 0.350 (0.140, 0.592) | 0.323 (0.306, 0.375) | 0.401 (0.372, 0.467) | 0.005, .871 | −0.007, .845 | −0**.012, .034** |
| CUN | 0.337 (0.137, 0.513) | 0.365 (0.321, 0.449) | 0.446 (0.391, 0.480) | −0.003, .898 | −0.016, .816 | −0.012, .115 |
| PERI | 0.310 (0.131, 0.506) | 0.267 (0.248, 0.328) | 0.362 (0.338, 0.392) | 0.007, .849 | −0.007, .842 | **−0.015, .014** |
| LOC | 0.451 (0.205, 0.540) | 0.443 (0.407, 0.516) | 0.478 (0.417, 0.541) | 0.001, .934 | −0.005, .891 | −0.006, .461 |
| HIPPO | 0.609 (0.455, 0.646) | 0.174 (0.128, 0.421) | 0.377 (0.335, 0.453) | 0.067, .119 | 0.035, .543 | **−0.032, .031** |
| AMYG | 0.519 (0.358, 0.614) | 0.175 (0.136, 0.423) | 0.366 (0.310, 0.468) | 0.053, .366 | 0.023, .816 | **−0.030, .049** |
| CAUD | 0.382 (0.000, 0.473) | 0.149 (0.131, 0.316) | 0.278 (0.254, 0.347) | 0.036, .404 | 0.016, .845 | **−0.020, .034** |
| PUT | 0.540 (0.357, 0.607) | 0.185 (0.154, 0.378) | 0.346 (0.309, 0.406) | 0.054, .119 | 0.029, .816 | **−0.025, .031** |
| NACC | 0.587 (0.442, 0.640) | 0.129 (0.108, 0.274) | 0.323 (0.286, 0.400) | **0.070, .025** | 0.040, .543 | **−0.030, .014** |
| PALL | 0.392 (0.291, 0.514) | 0.149 (0.132, 0.322) | 0.258 (0.230, 0.291) | 0.037, .366 | 0.020, .816 | **−0.017, .031** |
| THAL | 0.523 (0.416, 0.605) | 0.170 (0.143, 0.442) | 0.328 (0.290, 0.439) | 0.054, .366 | 0.029, .816 | −0.025, .127 |

Means were calculated across a range of density thresholds ($K$ = 0.10–0.25, $\Delta K$ = 0.01) and are reported with 95% bootstrap confidence intervals (CIs; 5000 resamples). Group differences in area under the curve (ΔAUC) were assessed using permutation testing (10 000 permutations). Bold values indicate statistically significant group differences after Benjamini–Hochberg correction for multiple comparisons ($p_{BH} < .05$). Abbreviations: SZ+A, schizophrenia with aggression; SZ−A, schizophrenia without aggression; HC, healthy controls; FPOL, frontal pole; POP, pars opercularis; PTRI, pars triangularis; PORB, pars orbitalis; MOFC, medial orbitofrontal cortex; LOFC, lateral orbitofrontal cortex; CMFG, caudal middle frontal gyrus; RMFG, rostral middle frontal gyrus; SFG, superior frontal gyrus; PARL, paracentral lobule; PREG, precentral gyrus; CACC, caudal anterior cingulate cortex; RACC, rostral anterior cingulate cortex; PCC, posterior cingulate cortex; ISTH, isthmus of the cingulate cortex; INS, insular cortex; TPOL, temporal pole; ENTH, entorhinal cortex; PHG, parahippocampal gyrus; TVTG, transverse temporal gyrus; ITG, inferior temporal gyrus; MTG, middle temporal gyrus; STG, superior temporal gyrus; BANKS, banks of the superior temporal sulcus; FUSI, fusiform gyrus; POSG, postcentral gyrus; PREC, precuneus; SMG, supramarginal gyrus; IPL, inferior parietal lobule; SPL, superior parietal lobule; LING, lingual gyrus; CUN, cuneus; PERI, pericalcarine cortex; LOC, lateral occipital cortex; HIPPO, hippocampus; AMYG, amygdala; CAUD, caudate nucleus; PUT, putamen; NACC, nucleus accumbens; PALL, globus pallidus; THAL, thalamus.

**Table A5.12.** Group differences in within-module degree using PANSS P7 'Hostility' scores only

| | M (95% CI) | | | ΔAUC, $p_{BH}$ | | |
|---|---|---|---|---|---|---|
| Node | SZ+A (*N* = 97) | SZ-A (*N* = 1477) | HC (*N* = 3058) | SZ+A vs SZ-A | SZ+A vs HC | SZ-A vs HC |
| FPOL | −1.21 (−1.77, 0.48) | 0.00 (0.00, 0.00) | −1.76 (−1.87, 0.00) | −0.180, .594 | 0.083, .920 | 0.263, .123 |
| POP | −0.41 (−0.99, 1.14) | −0.32 (−1.42, 0.00) | −1.07 (−1.46, −0.50) | −0.015, .998 | 0.096, .867 | 0.110, .312 |
| PTRI | −0.34 (−1.12, 1.04) | 0.65 (−1.20, 1.15) | −0.78 (−1.24, −0.50) | −0.149, .582 | 0.067, .867 | 0.215, .115 |
| PORB | −0.30 (−1.32, 0.67) | −0.32 (−1.22, −0.11) | −0.87 (−1.20, −0.35) | 0.003, .998 | 0.084, .867 | 0.081, .286 |
| MOFC | 1.05 (−0.59, 2.14) | 0.37 (−0.19, 1.17) | 0.08 (−0.50, 0.62) | 0.100, .659 | 0.143, .867 | 0.043, .863 |
| LOFC | 0.69 (−0.44, 1.91) | 1.42 (0.81, 2.28) | 2.10 (1.13, 2.68) | −0.114, .622 | −0.217, .867 | −0.103, .576 |
| CMFG | 0.35 (−1.05, 1.41) | −0.20 (−0.66, 0.28) | −0.02 (−0.36, 0.37) | 0.083, .646 | 0.061, .867 | −0.023, .869 |
| RMFG | 1.23 (−0.62, 1.81) | 1.25 (0.61, 2.12) | 1.25 (0.38, 1.90) | 0.000, .998 | 0.000, .998 | 0.000, .999 |
| SFG | 2.37 (0.50, 2.80) | 1.83 (1.11, 2.49) | 2.22 (1.31, 2.85) | 0.082, .619 | 0.024, .997 | −0.058, .863 |
| PARL | −0.54 (−1.58, 0.78) | −0.21 (−0.70, 0.23) | −0.11 (−0.67, 0.46) | −0.052, .756 | −0.064, .867 | −0.012, .952 |
| PREG | −0.60 (−1.37, 1.37) | 0.44 (−0.30, 1.17) | 0.18 (−0.36, 0.85) | −0.159, .606 | −0.118, .867 | 0.041, .869 |
| CACC | −0.85 (−1.63, 0.13) | −1.39 (−1.61, −1.04) | −1.24 (−1.53, −0.93) | 0.084, .594 | 0.061, .867 | −0.023, .836 |
| RACC | −0.68 (−1.16, 0.89) | 0.35 (−0.13, 0.83) | −0.02 (−0.49, 0.62) | −0.157, .582 | −0.099, .867 | 0.058, .824 |
| PCC | 0.03 (−1.38, 1.49) | 0.54 (−0.03, 1.11) | 0.56 (−0.13, 1.33) | −0.082, .734 | −0.082, .867 | 0.000, .999 |
| ISTH | −0.66 (−1.49, 1.25) | −1.11 (−1.48, −0.38) | −1.38 (−1.63, −0.52) | 0.067, .756 | 0.112, .867 | 0.045, .836 |
| INS | 3.11 (0.08, 3.24) | 0.36 (−0.18, 1.16) | −0.11 (−0.52, 0.48) | **0.413, .012** | **0.483, .029** | 0.070, .762 |
| TPOL | −0.90 (−1.60, 0.67) | 0.00 (0.00, 0.00) | −0.58 (−0.61, 0.00) | −0.136, .619 | −0.049, .996 | 0.087, .392 |
| ENTH | −1.15 (−1.75, 0.38) | 0.00 (0.00, 0.00) | 1.15 (0.00, 1.15) | −0.172, .594 | −0.346, .867 | −0.173, .286 |
| PHG | −0.54 (−1.35, 1.05) | −1.71 (−1.79, 0.00) | −0.58 (−1.04, −0.43) | 0.173, .622 | 0.004, .997 | −0.169, .115 |
| TVTG | −0.60 (−1.51, 0.82) | −0.85 (−1.24, −0.13) | −0.11 (−0.71, 0.27) | 0.039, .855 | −0.073, .867 | −0.112, .282 |
| ITG | 0.27 (−1.04, 1.53) | −0.79 (−1.16, −0.16) | −0.20 (−0.77, 0.37) | 0.161, .582 | 0.071, .867 | −0.090, .448 |
| MTG | −0.81 (−1.35, 0.68) | 0.12 (−0.38, 0.66) | −0.36 (−0.71, 0.12) | −0.141, .594 | −0.068, .867 | 0.073, .532 |
| STG | 0.20 (−1.32, 2.00) | 0.64 (0.02, 1.15) | 0.31 (−0.14, 0.88) | −0.070, .756 | −0.016, .997 | 0.053, .810 |
| BANKS | −0.45 (−1.21, 1.02) | −1.24 (−1.39, −0.81) | −1.12 (−1.33, −0.85) | 0.118, .594 | 0.099, .867 | −0.020, .845 |
| FUSI | 1.25 (−0.68, 2.54) | 0.38 (−0.51, 1.02) | −0.01 (−0.77, 0.77) | 0.136, .619 | 0.189, .867 | 0.053, .869 |

| | | | | | | |
|---|---|---|---|---|---|---|
| POSG | 0.99 (−0.48, 2.25) | −0.30 (−0.79, 0.51) | −0.10 (−0.62, 0.54) | 0.203, .582 | 0.170, .867 | −0.033, .869 |
| PREC | −0.25 (−1.15, 1.10) | 1.61 (0.53, 2.14) | 1.78 (1.11, 2.51) | −0.281, .582 | −0.306, .867 | −0.025, .947 |
| SMG | 0.94 (−0.62, 1.93) | −0.92 (−1.40, −0.00) | −0.01 (−0.51, 0.48) | 0.279, .384 | 0.140, .867 | −0.138, .115 |
| IPL | −0.66 (−1.35, 0.93) | 0.22 (−0.31, 0.73) | 0.14 (−0.34, 0.68) | −0.129, .594 | −0.120, .867 | 0.009, .952 |
| SPL | −0.47 (−1.22, 0.99) | −0.82 (−1.28, 0.08) | −0.73 (−1.30, −0.01) | 0.053, .844 | 0.041, .996 | −0.012, .952 |
| LING | −0.07 (−0.87, 1.42) | 0.81 (0.00, 0.87) | 0.92 (−0.74, 1.10) | −0.135, .619 | −0.151, .867 | −0.016, .952 |
| CUN | 1.02 (−1.38, 1.23) | 0.81 (0.00, 0.91) | −0.20 (−0.63, 0.38) | 0.031, .947 | 0.190, .738 | 0.159, .060 |
| PERI | 0.26 (−1.35, 0.84) | −0.81 (−0.92, 0.00) | 0.40 (−1.07, 0.65) | 0.160, .582 | −0.029, .997 | −0.189, .115 |
| LOC | −1.21 (−1.61, 0.47) | −0.81 (−0.94, −0.02) | 0.26 (−0.40, 0.62) | −0.056, .756 | −0.222, .738 | −0.166, .121 |
| HIPPO | 0.80 (−0.82, 1.88) | −0.70 (−1.01, 0.43) | −0.31 (−0.80, 0.37) | 0.229, .582 | 0.168, .867 | −0.061, .834 |
| AMYG | −0.11 (−1.41, 1.44) | −0.09 (−0.68, 0.23) | −0.18 (−0.45, 0.46) | −0.003, .998 | 0.013, .997 | 0.017, .952 |
| CAUD | −1.21 (−1.74, 0.05) | −0.03 (−0.46, 0.70) | −0.13 (−0.48, 0.36) | −0.179, .582 | −0.159, .867 | 0.020, .947 |
| PUT | 0.10 (−0.86, 1.67) | 1.63 (0.95, 1.90) | 1.59 (1.27, 1.84) | −0.231, .594 | −0.227, .867 | 0.005, .980 |
| NACC | 0.50 (−1.08, 1.62) | −1.33 (−1.53, −0.85) | 0.46 (−0.05, 0.81) | 0.273, .384 | 0.006, .997 | **−0.267, .004** |
| PALL | −1.01 (−1.54, 1.08) | −0.09 (−0.65, 0.40) | −1.45 (−1.73, −1.03) | −0.139, .582 | 0.068, .878 | **0.207, .004** |
| THAL | −0.13 (−1.14, 1.28) | 0.60 (−0.25, 1.03) | 0.02 (−0.82, 0.45) | −0.109, .756 | −0.030, .997 | 0.079, .810 |

Means were calculated across a range of density thresholds ($K$ = 0.10–0.25, $\Delta K$ = 0.01) and are reported with 95% bootstrap confidence intervals (CIs; 5000 resamples). Group differences in area under the curve (ΔAUC) were assessed using permutation testing (10 000 permutations). Bold values indicate statistically significant group differences after Benjamini–Hochberg correction for multiple comparisons ($p_{BH} < .05$). Abbreviations: SZ+A, schizophrenia with aggression; SZ−A, schizophrenia without aggression; HC, healthy controls; FPOL, frontal pole; POP, pars opercularis; PTRI, pars triangularis; PORB, pars orbitalis; MOFC, medial orbitofrontal cortex; LOFC, lateral orbitofrontal cortex; CMFG, caudal middle frontal gyrus; RMFG, rostral middle frontal gyrus; SFG, superior frontal gyrus; PARL, paracentral lobule; PREG, precentral gyrus; CACC, caudal anterior cingulate cortex; RACC, rostral anterior cingulate cortex; PCC, posterior cingulate cortex; ISTH, isthmus of the cingulate cortex; INS, insular cortex; TPOL, temporal pole; ENTH, entorhinal cortex; PHG, parahippocampal gyrus; TVTG, transverse temporal gyrus; ITG, inferior temporal gyrus; MTG, middle temporal gyrus; STG, superior temporal gyrus; BANKS, banks of the superior temporal sulcus; FUSI, fusiform gyrus; POSG, postcentral gyrus; PREC, precuneus; SMG, supramarginal gyrus; IPL, inferior parietal lobule; SPL, superior parietal lobule; LING, lingual gyrus; CUN, cuneus; PERI, pericalcarine cortex; LOC, lateral occipital cortex; HIPPO, hippocampus; AMYG, amygdala; CAUD, caudate nucleus; PUT, putamen; NACC, nucleus accumbens; PALL, globus pallidus; THAL, thalamus.

**Table A5.13.** Group differences in participation coefficient using PANSS P7 'Hostility' scores only

| | M (95% CI) | | | ΔAUC, $p_{BH}$ | | |
|---|---|---|---|---|---|---|
| Node | SZ+A (*N* = 97) | SZ-A (*N* = 1477) | HC (*N* = 3058) | SZ+A vs SZ-A | SZ+A vs HC | SZ-A vs HC |
| FPOL | 0.00 (0.00, 0.43) | 0.00 (0.00, 0.00) | 0.00 (0.00, 0.00) | 0.000, 1.000 | 0.000, 1.000 | 0.000, 1.000 |
| POP | 0.09 (0.00, 0.44) | 0.14 (0.00, 0.33) | 0.00 (0.00, 0.00) | −0.007, .984 | 0.014, 1.000 | 0.020, .127 |
| PTRI | 0.00 (0.00, 0.45) | 0.30 (0.00, 0.43) | 0.00 (0.00, 0.00) | −0.045, .735 | 0.000, 1.000 | 0.045, .127 |
| PORB | 0.00 (0.00, 0.39) | 0.44 (0.00, 0.49) | 0.02 (0.00, 0.16) | −0.066, .735 | −0.001, 1.000 | 0.065, .113 |
| MOFC | 0.00 (0.00, 0.46) | 0.09 (0.00, 0.14) | 0.01 (0.00, 0.11) | −0.014, .735 | −0.001, 1.000 | 0.014, .113 |
| LOFC | 0.00 (0.00, 0.54) | 0.19 (0.00, 0.22) | 0.06 (0.00, 0.11) | −0.028, .735 | −0.009, 1.000 | 0.019, .118 |
| CMFG | 0.00 (0.00, 0.55) | 0.00 (0.00, 0.03) | 0.00 (0.00, 0.07) | 0.000, 1.000 | 0.000, 1.000 | 0.000, 1.000 |
| RMFG | 0.00 (0.00, 0.48) | 0.23 (0.00, 0.26) | 0.09 (0.00, 0.18) | −0.035, .735 | −0.013, 1.000 | 0.022, .175 |
| SFG | 0.00 (0.00, 0.54) | 0.11 (0.00, 0.20) | 0.00 (0.00, 0.10) | −0.016, .735 | 0.000, 1.000 | 0.016, .371 |
| PARL | 0.00 (0.00, 0.50) | 0.00 (0.00, 0.00) | 0.00 (0.00, 0.00) | 0.000, 1.000 | 0.000, 1.000 | 0.000, 1.000 |
| PREG | 0.20 (0.00, 0.51) | 0.00 (0.00, 0.12) | 0.00 (0.00, 0.08) | 0.031, .735 | 0.031, 1.000 | 0.000, 1.000 |
| CACC | 0.00 (0.00, 0.42) | 0.00 (0.00, 0.00) | 0.00 (0.00, 0.00) | 0.000, 1.000 | 0.000, 1.000 | 0.000, 1.000 |
| RACC | 0.00 (0.00, 0.52) | 0.00 (0.00, 0.02) | 0.02 (0.00, 0.12) | 0.000, 1.000 | −0.002, 1.000 | −0.002, .565 |
| PCC | 0.00 (0.00, 0.53) | 0.00 (0.00, 0.07) | 0.00 (0.00, 0.10) | 0.000, 1.000 | 0.000, 1.000 | 0.000, 1.000 |
| ISTH | 0.26 (0.00, 0.60) | 0.14 (0.00, 0.39) | 0.49 (0.00, 0.50) | 0.019, 1.000 | −0.033, 1.000 | −0.051, .371 |
| INS | 0.00 (0.00, 0.63) | 0.11 (0.00, 0.20) | 0.00 (0.00, 0.09) | −0.017, .924 | 0.000, 1.000 | 0.017, .113 |
| TPOL | 0.00 (0.00, 0.58) | 0.00 (0.00, 0.06) | 0.00 (0.00, 0.38) | 0.000, 1.000 | 0.000, 1.000 | 0.000, 1.000 |
| ENTH | 0.00 (0.00, 0.50) | 0.00 (0.00, 0.22) | 0.00 (0.00, 0.50) | 0.000, 1.000 | 0.000, 1.000 | 0.000, 1.000 |
| PHG | 0.03 (0.00, 0.55) | 0.10 (0.00, 0.46) | 0.66 (0.60, 0.72) | −0.010, 1.000 | −0.095, 1.000 | **−0.085, .008** |
| TVTG | 0.00 (0.00, 0.55) | 0.00 (0.00, 0.09) | 0.00 (0.00, 0.02) | 0.000, 1.000 | 0.000, 1.000 | 0.000, 1.000 |
| ITG | 0.00 (0.00, 0.60) | 0.00 (0.00, 0.06) | 0.05 (0.00, 0.24) | 0.000, 1.000 | −0.007, 1.000 | −0.007, .872 |
| MTG | 0.00 (0.00, 0.51) | 0.00 (0.00, 0.08) | 0.00 (0.00, 0.10) | 0.000, 1.000 | 0.000, 1.000 | 0.000, 1.000 |
| STG | 0.00 (0.00, 0.56) | 0.00 (0.00, 0.08) | 0.00 (0.00, 0.08) | 0.000, 1.000 | 0.000, 1.000 | 0.000, 1.000 |
| BANKS | 0.00 (0.00, 0.57) | 0.00 (0.00, 0.00) | 0.00 (0.00, 0.00) | 0.000, 1.000 | 0.000, 1.000 | 0.000, 1.000 |
| FUSI | 0.14 (0.00, 0.63) | 0.16 (0.00, 0.37) | 0.33 (0.22, 0.45) | −0.002, 1.000 | −0.028, 1.000 | −0.026, .165 |

| | | | | | | |
|---|---|---|---|---|---|---|
| POSG | 0.03 (0.00, 0.56) | 0.00 (0.00, 0.03) | 0.00 (0.00, 0.11) | 0.004, .924 | 0.004, 1.000 | 0.000, 1.000 |
| PREC | 0.00 (0.00, 0.53) | 0.13 (0.01, 0.24) | 0.26 (0.00, 0.33) | −0.020, .735 | −0.039, 1.000 | −0.019, .669 |
| SMG | 0.00 (0.00, 0.53) | 0.04 (0.00, 0.13) | 0.00 (0.00, 0.05) | −0.006, .735 | 0.000, 1.000 | 0.006, .113 |
| IPL | 0.00 (0.00, 0.54) | 0.01 (0.00, 0.10) | 0.00 (0.00, 0.14) | −0.001, 1.000 | 0.000, 1.000 | 0.001, 1.000 |
| SPL | 0.00 (0.00, 0.54) | 0.11 (0.00, 0.29) | 0.32 (0.00, 0.43) | −0.016, .924 | −0.049, 1.000 | −0.033, .566 |
| LING | 0.36 (0.00, 0.57) | 0.12 (0.00, 0.48) | 0.11 (0.00, 0.52) | 0.039, .735 | 0.040, 1.000 | 0.002, 1.000 |
| CUN | 0.19 (0.00, 0.50) | 0.26 (0.00, 0.48) | 0.46 (0.00, 0.48) | −0.011, 1.000 | −0.042, 1.000 | −0.031, .566 |
| PERI | 0.21 (0.00, 0.47) | 0.00 (0.00, 0.14) | 0.00 (0.00, 0.19) | 0.032, .735 | 0.032, 1.000 | 0.000, 1.000 |
| LOC | 0.37 (0.00, 0.57) | 0.43 (0.34, 0.49) | 0.43 (0.00, 0.51) | −0.008, 1.000 | −0.010, 1.000 | −0.002, 1.000 |
| HIPPO | 0.00 (0.00, 0.59) | 0.08 (0.00, 0.41) | 0.35 (0.03, 0.48) | −0.010, 1.000 | −0.053, 1.000 | −0.043, .229 |
| AMYG | 0.00 (0.00, 0.55) | 0.05 (0.00, 0.43) | 0.18 (0.00, 0.42) | −0.005, 1.000 | −0.026, 1.000 | −0.020, 1.000 |
| CAUD | 0.00 (0.00, 0.43) | 0.00 (0.00, 0.07) | 0.00 (0.00, 0.00) | 0.000, 1.000 | 0.000, 1.000 | 0.000, 1.000 |
| PUT | 0.00 (0.00, 0.57) | 0.00 (0.00, 0.32) | 0.00 (0.00, 0.11) | 0.000, 1.000 | 0.000, 1.000 | 0.000, 1.000 |
| NACC | 0.06 (0.00, 0.60) | 0.00 (0.00, 0.00) | 0.00 (0.00, 0.15) | 0.009, 1.000 | 0.009, 1.000 | 0.000, 1.000 |
| PALL | 0.00 (0.00, 0.46) | 0.00 (0.00, 0.39) | 0.00 (0.00, 0.00) | 0.000, 1.000 | 0.000, 1.000 | 0.000, 1.000 |
| THAL | 0.00 (0.00, 0.57) | 0.00 (0.00, 0.47) | 0.00 (0.00, 0.26) | 0.000, 1.000 | 0.000, 1.000 | 0.000, 1.000 |

Means were calculated across a range of density thresholds ($K$ = 0.10–0.25, $\Delta K$ = 0.01) and are reported with 95% bootstrap confidence intervals (CIs; 5000 resamples). Group differences in area under the curve (ΔAUC) were assessed using permutation testing (10 000 permutations). Bold values indicate statistically significant group differences after Benjamini–Hochberg correction for multiple comparisons ($p_{BH} < .05$). Abbreviations: SZ+A, schizophrenia with aggression; SZ−A, schizophrenia without aggression; HC, healthy controls; FPOL, frontal pole; POP, pars opercularis; PTRI, pars triangularis; PORB, pars orbitalis; MOFC, medial orbitofrontal cortex; LOFC, lateral orbitofrontal cortex; CMFG, caudal middle frontal gyrus; RMFG, rostral middle frontal gyrus; SFG, superior frontal gyrus; PARL, paracentral lobule; PREG, precentral gyrus; CACC, caudal anterior cingulate cortex; RACC, rostral anterior cingulate cortex; PCC, posterior cingulate cortex; ISTH, isthmus of the cingulate cortex; INS, insular cortex; TPOL, temporal pole; ENTH, entorhinal cortex; PHG, parahippocampal gyrus; TVTG, transverse temporal gyrus; ITG, inferior temporal gyrus; MTG, middle temporal gyrus; STG, superior temporal gyrus; BANKS, banks of the superior temporal sulcus; FUSI, fusiform gyrus; POSG, postcentral gyrus; PREC, precuneus; SMG, supramarginal gyrus; IPL, inferior parietal lobule; SPL, superior parietal lobule; LING, lingual gyrus; CUN, cuneus; PERI, pericalcarine cortex; LOC, lateral occipital cortex; HIPPO, hippocampus; AMYG, amygdala; CAUD, caudate nucleus; PUT, putamen; NACC, nucleus accumbens; PALL, globus pallidus; THAL, thalamus.

**Table A5.14.** Group differences in canonical brain network density using PANSS P7 'Hostility' scores only

| | M (95% CI) | | | ΔAUC, $p_{BH}$ | | |
|---|---|---|---|---|---|---|
| Metric | SZ+A (*N* = 97) | SZ-A (*N* = 1477) | HC (*N* = 3058) | SZ+A vs SZ-A | SZ+A vs HC | SZ-A vs HC |
| *Salience network* | | | | | | |
| Network-rest | 0.30 (0.16, 0.32) | 0.19 (0.14, 0.24) | 0.14 (0.12, 0.18) | 0.017, .154 | **0.024, .035** | 0.007, .406 |
| *Central executive network* | | | | | | |
| Within-network | 0.34 (0.00, 0.48) | 0.52 (0.36, 0.66) | 0.29 (0.17, 0.51) | −0.026, .735 | 0.010, 1.000 | 0.036, .322 |
| Network-rest | 0.18 (0.12, 0.24) | 0.25 (0.21, 0.28) | 0.24 (0.21, 0.26) | −0.010, .249 | −0.009, .673 | 0.001, .737 |
| *Default mode network* | | | | | | |
| Within-network | 0.19 (0.11, 0.27) | 0.15 (0.13, 0.19) | 0.18 (0.15, 0.20) | 0.005, .735 | 0.001, 1.000 | −0.003, .487 |
| Network-rest | 0.15 (0.13, 0.18) | 0.16 (0.14, 0.17) | 0.15 (0.14, 0.16) | −0.001, .776 | 0.001, .828 | 0.001, .472 |
| *Language network* | | | | | | |
| Within-network | 0.28 (0.10, 0.55) | 0.37 (0.30, 0.40) | 0.31 (0.30, 0.38) | −0.014, .735 | −0.005, 1.000 | 0.009, .487 |
| Network-rest | 0.13 (0.11, 0.20) | 0.18 (0.15, 0.21) | 0.16 (0.14, 0.18) | −0.007, .360 | −0.004, .828 | 0.003, .472 |
| *Reward network* | | | | | | |
| Within-network | 0.38 (0.18, 0.46) | 0.31 (0.24, 0.36) | 0.39 (0.33, 0.43) | 0.009, .735 | −0.003, 1.000 | −0.012, .322 |
| Network-rest | 0.15 (0.12, 0.19) | 0.16 (0.14, 0.17) | 0.14 (0.12, 0.15) | −0.002, .776 | 0.001, .828 | 0.003, .407 |
| *Affective control network* | | | | | | |
| Within-network | 0.38 (0.25, 0.42) | 0.31 (0.27, 0.36) | 0.32 (0.28, 0.34) | 0.010, .735 | 0.009, 1.000 | −0.001, 1.000 |
| Network-rest | 0.14 (0.12, 0.17) | 0.17 (0.16, 0.18) | 0.14 (0.14, 0.15) | −0.005, .098 | −0.001, .828 | **0.004, .007** |

Means were calculated across a range of density thresholds ($K$ = 0.10–0.25, $\Delta K$ = 0.01) and are reported with 95% bootstrap confidence intervals (CIs; 5000 resamples). Group differences in area under the curve (ΔAUC) were assessed using permutation testing (10 000 permutations). Bold values indicate statistically significant group differences after Benjamini–Hochberg correction for multiple comparisons ($p_{BH}$ < .05). Abbreviations: SZ+A, schizophrenia with aggression; SZ−A, schizophrenia without aggression; HC, healthy controls.

**Table A5.15.** Group differences in global network topology using regional volumes residualized for CPZ-equivalent dose.

| | M (95% CI) | | | ΔAUC, $p_{BH}$ | | |
|---|---|---|---|---|---|---|
| Metric | SZ+A (*N* = 212) | SZ-A (*N* = 1533) | HC (*N* = 3058) | SZ+A vs SZ-A | SZ+A vs HC | SZ-A vs HC |
| Transitivity | 0.45 (0.39, 0.51) | 0.55 (0.50, 0.58) | 0.50 (0.46, 0.52) | −0.015, .166 | −0.007, .893 | **0.008, .037** |
| Modularity | 0.39 (0.31, 0.42) | 0.31 (0.28, 0.34) | 0.39 (0.36, 0.42) | **0.012, .040** | −0.001, .964 | **−0.013, .005** |
| Efficiency | 0.47 (0.44, 0.51) | 0.40 (0.38, 0.45) | 0.45 (0.44, 0.48) | 0.010, .166 | 0.003, .893 | **−0.008, .010** |
| Small-worldness | 1.58 (1.33, 1.83) | 1.19 (1.15, 1.37) | 1.47 (1.33, 1.65) | **0.058, .040** | 0.018, .893 | **−0.040, .037** |

Means were calculated across a range of density thresholds ($K = 0.10–0.25$, $\Delta K = 0.01$) and are reported with 95% bootstrap confidence intervals (CIs; 5000 resamples). Group differences in area under the curve (ΔAUC) were assessed using permutation testing (10 000 permutations). Bold values indicate statistically significant group differences after Benjamini–Hochberg correction for multiple comparisons ($p_{BH} < .05$). Abbreviations: SZ+A, schizophrenia with aggression; SZ−A, schizophrenia without aggression; HC, healthy controls.

**Table A5.16.** Group differences in degree using regional volumes residualized for CPZ-equivalent dose.

| | M (95% CI) | | | ΔAUC, $p_{BH}$ | | |
|---|---|---|---|---|---|---|
| Node | SZ+A (*N* = 212) | SZ-A (*N* = 1533) | HC (*N* = 3058) | SZ+A vs SZ-A | SZ+A vs HC | SZ-A vs HC |
| FPOL | 1.2 (0.0, 6.1) | 0.1 (0.0, 1.2) | 1.0 (0.0, 2.1) | 0.185, .748 | 0.045, .882 | −0.140, .640 |
| POP | 2.1 (1.0, 8.4) | 3.4 (1.4, 6.9) | 3.9 (2.4, 6.4) | −0.196, .899 | −0.280, .860 | −0.084, .956 |
| PTRI | 4.6 (1.9, 8.3) | 5.5 (3.5, 6.9) | 5.1 (3.3, 6.1) | −0.135, .899 | −0.079, .882 | 0.055, .956 |
| PORB | 5.1 (2.4, 8.1) | 4.8 (3.2, 6.1) | 4.7 (3.4, 7.2) | 0.050, .944 | 0.076, .882 | 0.026, .975 |
| MOFC | 12.5 (7.3, 16.9) | 13.1 (9.5, 15.9) | 9.0 (6.5, 11.2) | −0.070, .944 | 0.537, .860 | 0.606, .302 |
| LOFC | 9.6 (6.5, 13.9) | 18.1 (13.8, 20.7) | 18.2 (13.4, 20.4) | −1.305, .692 | −1.326, .860 | −0.020, .985 |
| CMFG | 6.1 (2.8, 10.6) | 7.6 (5.3, 9.9) | 8.2 (6.8, 10.0) | −0.239, .899 | −0.304, .860 | −0.065, .956 |
| RMFG | 12.8 (6.8, 16.2) | 18.1 (13.4, 20.9) | 14.9 (10.5, 17.8) | −0.801, .748 | −0.305, .860 | 0.495, .649 |
| SFG | 15.2 (10.6, 18.0) | 18.5 (13.8, 22.4) | 17.9 (13.6, 21.3) | −0.497, .748 | −0.430, .860 | 0.067, .975 |
| PARL | 5.6 (1.9, 9.9) | 9.3 (6.6, 10.6) | 8.1 (5.4, 10.4) | −0.566, .748 | −0.361, .860 | 0.205, .854 |
| PREG | 7.5 (4.0, 12.7) | 10.0 (6.3, 14.2) | 9.3 (6.9, 12.4) | −0.376, .899 | −0.270, .860 | 0.105, .956 |
| CACC | 4.4 (2.6, 9.2) | 2.7 (2.0, 4.0) | 3.1 (2.1, 4.4) | 0.240, .794 | 0.180, .860 | −0.060, .854 |
| RACC | 10.4 (6.7, 13.1) | 11.7 (8.5, 13.6) | 8.5 (6.4, 11.2) | −0.190, .899 | 0.305, .860 | 0.495, .383 |
| PCC | 8.2 (2.9, 12.9) | 13.1 (9.6, 15.9) | 10.8 (7.9, 14.8) | −0.726, .748 | −0.379, .860 | 0.347, .759 |
| ISTH | 4.6 (1.0, 10.0) | 3.6 (1.6, 6.8) | 3.6 (2.6, 6.6) | 0.155, .899 | 0.151, .860 | −0.005, .985 |
| INS | 17.6 (9.8, 19.9) | 12.2 (8.6, 16.0) | 7.9 (6.2, 10.7) | 0.816, .748 | 1.461, .076 | 0.645, .242 |
| TPOL | 1.6 (0.0, 5.2) | 0.2 (0.0, 1.0) | 1.0 (0.9, 1.9) | 0.215, .748 | 0.090, .860 | −0.125, .066 |
| ENTH | 2.0 (0.1, 4.3) | 0.4 (0.0, 1.7) | 2.0 (1.9, 2.6) | 0.255, .692 | 0.005, .887 | **−0.250, .004** |
| PHG | 4.8 (1.6, 7.2) | 1.9 (1.0, 3.3) | 3.4 (2.9, 5.4) | 0.450, .692 | 0.215, .860 | −0.235, .069 |
| TVTG | 6.6 (4.4, 10.9) | 5.0 (2.8, 8.7) | 8.2 (5.6, 9.8) | 0.245, .899 | −0.240, .860 | −0.485, .283 |
| ITG | 10.8 (5.8, 15.1) | 5.1 (2.9, 8.1) | 8.1 (5.4, 11.1) | 0.865, .692 | 0.399, .860 | −0.466, .380 |
| MTG | 8.1 (5.1, 13.0) | 10.9 (7.8, 13.9) | 6.9 (5.4, 9.1) | −0.430, .854 | 0.170, .860 | 0.600, .200 |
| STG | 14.8 (9.5, 19.2) | 12.9 (9.4, 15.5) | 9.8 (7.8, 12.4) | 0.266, .899 | 0.741, .860 | 0.476, .380 |
| BANKS | 3.8 (2.0, 6.3) | 3.0 (3.0, 4.0) | 3.6 (3.0, 4.7) | 0.115, .899 | 0.035, .882 | −0.080, .649 |
| FUSI | 13.0 (6.6, 17.3) | 12.8 (8.9, 16.4) | 10.7 (6.8, 14.6) | 0.051, .944 | 0.349, .860 | 0.299, .854 |

| | | | | | | |
|---|---|---|---|---|---|---|
| POSG | 12.6 (7.0, 16.9) | 8.8 (5.4, 12.9) | 8.2 (5.9, 11.2) | 0.604, .789 | 0.671, .860 | 0.066, .975 |
| PREC | 11.2 (5.8, 15.3) | 16.6 (10.3, 20.1) | 18.9 (15.2, 21.0) | −0.820, .755 | −1.167, .860 | −0.347, .709 |
| SMG | 7.2 (4.5, 11.3) | 5.2 (2.6, 8.8) | 8.6 (6.2, 10.6) | 0.315, .899 | −0.202, .860 | −0.518, .242 |
| IPL | 5.8 (3.2, 10.4) | 9.0 (6.0, 12.4) | 9.2 (7.0, 11.7) | −0.495, .852 | −0.524, .860 | −0.029, .975 |
| SPL | 6.1 (3.0, 11.0) | 5.4 (2.6, 9.9) | 6.8 (4.2, 9.8) | 0.079, .944 | −0.101, .882 | −0.180, .917 |
| LING | 3.5 (2.3, 7.0) | 3.2 (2.8, 4.0) | 4.4 (4.0, 5.8) | 0.050, .942 | −0.125, .860 | −0.175, .051 |
| CUN | 3.9 (2.8, 7.9) | 3.0 (3.0, 4.0) | 5.1 (4.1, 6.0) | 0.130, .854 | −0.185, .860 | −0.315, .058 |
| PERI | 3.0 (2.0, 4.8) | 2.0 (2.0, 2.6) | 3.4 (2.9, 3.9) | 0.150, .692 | −0.065, .860 | **−0.215, .004** |
| LOC | 4.4 (1.6, 10.2) | 5.4 (3.9, 8.1) | 6.1 (4.4, 8.8) | −0.150, .899 | −0.251, .860 | −0.101, .923 |
| HIPPO | 3.8 (2.2, 7.6) | 3.6 (2.4, 6.3) | 4.4 (3.4, 5.5) | 0.030, .944 | −0.105, .860 | −0.136, .649 |
| AMYG | 6.9 (3.1, 10.9) | 3.6 (2.6, 6.8) | 4.2 (3.2, 6.2) | 0.510, .748 | 0.399, .860 | −0.111, .854 |
| CAUD | 3.0 (1.1, 4.2) | 3.2 (2.4, 4.0) | 3.6 (2.8, 4.4) | −0.040, .944 | −0.095, .860 | −0.055, .807 |
| PUT | 7.0 (3.6, 9.3) | 5.2 (3.7, 6.1) | 5.8 (4.9, 6.4) | 0.276, .748 | 0.185, .860 | −0.091, .648 |
| NACC | 6.0 (2.4, 10.8) | 1.9 (1.2, 2.8) | 4.4 (3.3, 5.2) | 0.595, .692 | 0.230, .860 | **−0.365, .004** |
| PALL | 4.6 (2.2, 8.4) | 2.9 (2.1, 3.2) | 2.0 (1.6, 2.0) | 0.234, .692 | 0.379, .076 | **0.145, .007** |
| THAL | 5.1 (2.2, 9.2) | 4.1 (2.3, 6.2) | 3.9 (2.4, 5.6) | 0.151, .899 | 0.170, .860 | 0.020, .975 |

Means were calculated across a range of density thresholds ($K$ = 0.10–0.25, $\Delta K$ = 0.01) and are reported with 95% bootstrap confidence intervals (CIs; 5000 resamples). Group differences in area under the curve (ΔAUC) were assessed using permutation testing (10 000 permutations). Bold values indicate statistically significant group differences after Benjamini–Hochberg correction for multiple comparisons ($p_{BH}$ < .05). Abbreviations: SZ+A, schizophrenia with aggression; SZ−A, schizophrenia without aggression; HC, healthy controls; FPOL, frontal pole; POP, pars opercularis; PTRI, pars triangularis; PORB, pars orbitalis; MOFC, medial orbitofrontal cortex; LOFC, lateral orbitofrontal cortex; CMFG, caudal middle frontal gyrus; RMFG, rostral middle frontal gyrus; SFG, superior frontal gyrus; PARL, paracentral lobule; PREG, precentral gyrus; CACC, caudal anterior cingulate cortex; RACC, rostral anterior cingulate cortex; PCC, posterior cingulate cortex; ISTH, isthmus of the cingulate cortex; INS, insular cortex; TPOL, temporal pole; ENTH, entorhinal cortex; PHG, parahippocampal gyrus; TVTG, transverse temporal gyrus; ITG, inferior temporal gyrus; MTG, middle temporal gyrus; STG, superior temporal gyrus; BANKS, banks of the superior temporal sulcus; FUSI, fusiform gyrus; POSG, postcentral gyrus; PREC, precuneus; SMG, supramarginal gyrus; IPL, inferior parietal lobule; SPL, superior parietal lobule; LING, lingual gyrus; CUN, cuneus; PERI, pericalcarine cortex; LOC, lateral occipital cortex; HIPPO, hippocampus; AMYG, amygdala; CAUD, caudate nucleus; PUT, putamen; NACC, nucleus accumbens; PALL, globus pallidus; THAL, thalamus.

**Table A5.17.** Group differences in betweenness centrality using regional volumes residualized for CPZ-equivalent dose.

| | M (95% CI) | | | ΔAUC, $p_{BH}$ | | |
|---|---|---|---|---|---|---|
| Node | SZ+A (*N* = 212) | SZ-A (*N* = 1533) | HC (*N* = 3058) | SZ+A vs SZ-A | SZ+A vs HC | SZ-A vs HC |
| FPOL | 0.000 (0.000, 0.007) | 0.000 (0.000, 0.000) | 0.000 (0.000, 0.000) | 0.00000, 1.000 | 0.00000, 1.000 | 0.00000, 1.000 |
| POP | 0.000 (0.000, 0.031) | 0.000 (0.000, 0.006) | 0.001 (0.000, 0.007) | 0.00000, 1.000 | −0.00007, .995 | −0.00007, .896 |
| PTRI | 0.025 (0.001, 0.049) | 0.015 (0.001, 0.035) | 0.004 (0.001, 0.022) | 0.00151, 1.000 | 0.00314, .835 | 0.00163, .885 |
| PORB | 0.003 (0.000, 0.039) | 0.003 (0.000, 0.032) | 0.002 (0.000, 0.027) | −0.00002, 1.000 | 0.00021, .995 | 0.00022, 1.000 |
| MOFC | 0.035 (0.009, 0.135) | 0.017 (0.005, 0.055) | 0.018 (0.002, 0.043) | 0.00296, 1.000 | 0.00260, .893 | −0.00035, 1.000 |
| LOFC | 0.017 (0.007, 0.106) | 0.070 (0.028, 0.154) | 0.117 (0.037, 0.192) | −0.00816, 1.000 | −0.01520, .843 | −0.00704, .885 |
| CMFG | 0.003 (0.000, 0.050) | 0.001 (0.000, 0.007) | 0.016 (0.002, 0.028) | 0.00032, 1.000 | −0.00179, .893 | −0.00211, .266 |
| RMFG | 0.058 (0.014, 0.125) | 0.091 (0.031, 0.185) | 0.069 (0.022, 0.131) | −0.00507, 1.000 | −0.00160, .995 | 0.00347, .896 |
| SFG | 0.070 (0.024, 0.135) | 0.111 (0.020, 0.259) | 0.081 (0.030, 0.210) | −0.00619, 1.000 | −0.00127, .995 | 0.00492, .896 |
| PARL | 0.002 (0.000, 0.023) | 0.003 (0.001, 0.008) | 0.004 (0.001, 0.012) | −0.00008, 1.000 | −0.00028, .995 | −0.00020, .896 |
| PREG | 0.005 (0.001, 0.069) | 0.005 (0.001, 0.033) | 0.011 (0.004, 0.068) | −0.00001, 1.000 | −0.00075, .995 | −0.00074, .885 |
| CACC | 0.002 (0.000, 0.035) | 0.000 (0.000, 0.000) | 0.000 (0.000, 0.000) | 0.00032, .984 | 0.00032, .893 | 0.00000, 1.000 |
| RACC | 0.050 (0.013, 0.120) | 0.013 (0.007, 0.029) | 0.011 (0.005, 0.040) | 0.00542, .707 | 0.00578, .843 | 0.00036, 1.000 |
| PCC | 0.014 (0.002, 0.089) | 0.022 (0.012, 0.065) | 0.036 (0.012, 0.119) | −0.00120, 1.000 | −0.00337, .979 | −0.00217, .885 |
| ISTH | 0.034 (0.000, 0.125) | 0.008 (0.000, 0.036) | 0.022 (0.007, 0.056) | 0.00413, .984 | 0.00222, .930 | −0.00191, .718 |
| INS | 0.179 (0.033, 0.244) | 0.028 (0.006, 0.157) | 0.005 (0.002, 0.052) | 0.02357, .707 | 0.02641, .369 | 0.00284, .588 |
| TPOL | 0.007 (0.000, 0.054) | 0.000 (0.000, 0.006) | 0.000 (0.000, 0.013) | 0.00106, .984 | 0.00106, .893 | 0.00000, 1.000 |
| ENTH | 0.004 (0.000, 0.035) | 0.009 (0.000, 0.030) | 0.049 (0.018, 0.050) | −0.00058, 1.000 | −0.00676, .843 | −0.00618, .057 |
| PHG | 0.017 (0.000, 0.097) | 0.071 (0.000, 0.160) | 0.297 (0.131, 0.346) | −0.00842, 1.000 | −0.04230, .835 | −0.03388, .150 |
| TVTG | 0.002 (0.001, 0.031) | 0.001 (0.000, 0.008) | 0.008 (0.002, 0.026) | 0.00022, 1.000 | −0.00087, .930 | −0.00109, .494 |
| ITG | 0.067 (0.011, 0.160) | 0.000 (0.000, 0.011) | 0.086 (0.006, 0.173) | 0.01011, .707 | −0.00250, .995 | −0.01261, .266 |
| MTG | 0.026 (0.007, 0.107) | 0.020 (0.009, 0.063) | 0.012 (0.004, 0.050) | 0.00092, 1.000 | 0.00206, .930 | 0.00114, .885 |
| STG | 0.062 (0.017, 0.197) | 0.036 (0.021, 0.087) | 0.037 (0.017, 0.088) | 0.00365, 1.000 | 0.00347, .893 | −0.00018, 1.000 |
| BANKS | 0.001 (0.000, 0.018) | 0.000 (0.000, 0.001) | 0.001 (0.000, 0.002) | 0.00011, 1.000 | 0.00005, .995 | −0.00006, .425 |
| FUSI | 0.175 (0.052, 0.264) | 0.212 (0.066, 0.304) | 0.322 (0.157, 0.382) | −0.00546, 1.000 | −0.02216, .995 | −0.01671, .688 |

| | | | | | | |
|---|---|---|---|---|---|---|
| POSG | 0.051 (0.009, 0.144) | 0.007 (0.002, 0.025) | 0.013 (0.003, 0.035) | 0.00690, .707 | 0.00589, .835 | −0.00101, .885 |
| PREC | 0.067 (0.010, 0.163) | 0.083 (0.031, 0.187) | 0.255 (0.128, 0.336) | −0.00256, 1.000 | −0.02752, .893 | −0.02496, .266 |
| SMG | 0.007 (0.001, 0.045) | 0.002 (0.000, 0.010) | 0.011 (0.004, 0.027) | 0.00075, 1.000 | −0.00061, .995 | −0.00137, .266 |
| IPL | 0.008 (0.001, 0.056) | 0.016 (0.007, 0.045) | 0.029 (0.013, 0.111) | −0.00118, 1.000 | −0.00309, .893 | −0.00192, .885 |
| SPL | 0.012 (0.000, 0.077) | 0.002 (0.000, 0.034) | 0.019 (0.003, 0.045) | 0.00152, 1.000 | −0.00055, .995 | −0.00207, .722 |
| LING | 0.004 (0.000, 0.081) | 0.023 (0.008, 0.031) | 0.010 (0.002, 0.056) | −0.00282, 1.000 | −0.00061, .995 | 0.00222, .328 |
| CUN | 0.008 (0.001, 0.096) | 0.019 (0.008, 0.038) | 0.020 (0.005, 0.039) | −0.00164, 1.000 | −0.00152, .930 | 0.00012, 1.000 |
| PERI | 0.000 (0.000, 0.018) | 0.000 (0.000, 0.000) | 0.000 (0.000, 0.001) | 0.00002, 1.000 | −0.00001, .995 | −0.00003, .150 |
| LOC | 0.054 (0.001, 0.134) | 0.115 (0.070, 0.137) | 0.051 (0.019, 0.100) | −0.00909, 1.000 | 0.00016, 1.000 | 0.00925, .150 |
| HIPPO | 0.003 (0.001, 0.064) | 0.089 (0.002, 0.179) | 0.173 (0.054, 0.228) | −0.01349, 1.000 | −0.02585, .467 | −0.01235, .718 |
| AMYG | 0.027 (0.003, 0.129) | 0.056 (0.006, 0.164) | 0.104 (0.015, 0.176) | −0.00470, 1.000 | −0.01066, .930 | −0.00596, .885 |
| CAUD | 0.001 (0.000, 0.015) | 0.001 (0.000, 0.011) | 0.001 (0.001, 0.014) | −0.00004, 1.000 | −0.00004, .995 | −0.00000, 1.000 |
| PUT | 0.030 (0.007, 0.094) | 0.056 (0.016, 0.103) | 0.074 (0.042, 0.092) | −0.00394, 1.000 | −0.00648, .893 | −0.00254, .885 |
| NACC | 0.042 (0.003, 0.127) | 0.000 (0.000, 0.007) | 0.013 (0.003, 0.061) | 0.00610, .707 | 0.00418, .893 | −0.00192, .300 |
| PALL | 0.006 (0.001, 0.055) | 0.004 (0.000, 0.028) | 0.000 (0.000, 0.000) | 0.00027, 1.000 | 0.00084, .503 | **0.00057, .025** |
| THAL | 0.051 (0.001, 0.106) | 0.041 (0.006, 0.115) | 0.019 (0.002, 0.115) | 0.00167, 1.000 | 0.00488, .893 | 0.00322, .885 |

Means were calculated across a range of density thresholds ($K$ = 0.10–0.25, $\Delta K$ = 0.01) and are reported with 95% bootstrap confidence intervals (CIs; 5000 resamples). Group differences in area under the curve (ΔAUC) were assessed using permutation testing (10 000 permutations). Bold values indicate statistically significant group differences after Benjamini–Hochberg correction for multiple comparisons ($p_{BH} < .05$). Abbreviations: SZ+A, schizophrenia with aggression; SZ−A, schizophrenia without aggression; HC, healthy controls; FPOL, frontal pole; POP, pars opercularis; PTRI, pars triangularis; PORB, pars orbitalis; MOFC, medial orbitofrontal cortex; LOFC, lateral orbitofrontal cortex; CMFG, caudal middle frontal gyrus; RMFG, rostral middle frontal gyrus; SFG, superior frontal gyrus; PARL, paracentral lobule; PREG, precentral gyrus; CACC, caudal anterior cingulate cortex; RACC, rostral anterior cingulate cortex; PCC, posterior cingulate cortex; ISTH, isthmus of the cingulate cortex; INS, insular cortex; TPOL, temporal pole; ENTH, entorhinal cortex; PHG, parahippocampal gyrus; TVTG, transverse temporal gyrus; ITG, inferior temporal gyrus; MTG, middle temporal gyrus; STG, superior temporal gyrus; BANKS, banks of the superior temporal sulcus; FUSI, fusiform gyrus; POSG, postcentral gyrus; PREC, precuneus; SMG, supramarginal gyrus; IPL, inferior parietal lobule; SPL, superior parietal lobule; LING, lingual gyrus; CUN, cuneus; PERI, pericalcarine cortex; LOC, lateral occipital cortex; HIPPO, hippocampus; AMYG, amygdala; CAUD, caudate nucleus; PUT, putamen; NACC, nucleus accumbens; PALL, globus pallidus; THAL, thalamus.

**Table A5.18.** Group differences in harmonic centrality using regional volumes residualized for CPZ-equivalent dose.

| | M (95% CI) | | | ΔAUC, $p_{BH}$ | | |
|---|---|---|---|---|---|---|
| Node | SZ+A (*N* = 212) | SZ-A (*N* = 1533) | HC (*N* = 3058) | SZ+A vs SZ-A | SZ+A vs HC | SZ-A vs HC |
| FPOL | 0.309 (0.000, 0.492) | 0.026 (0.000, 0.304) | 0.204 (0.000, 0.377) | 0.045, .749 | 0.017, .920 | −0.028, .589 |
| POP | 0.369 (0.261, 0.527) | 0.376 (0.301, 0.471) | 0.412 (0.359, 0.472) | −0.001, .961 | −0.007, .920 | −0.006, .671 |
| PTRI | 0.461 (0.301, 0.527) | 0.438 (0.381, 0.480) | 0.441 (0.393, 0.469) | 0.004, .908 | 0.003, .920 | −0.001, .921 |
| PORB | 0.469 (0.393, 0.536) | 0.428 (0.396, 0.467) | 0.445 (0.423, 0.496) | 0.007, .759 | 0.004, .920 | −0.003, .624 |
| MOFC | 0.597 (0.513, 0.671) | 0.551 (0.511, 0.608) | 0.520 (0.480, 0.562) | 0.007, .759 | 0.012, .730 | 0.004, .624 |
| LOFC | 0.560 (0.501, 0.631) | 0.625 (0.572, 0.682) | 0.655 (0.590, 0.696) | −0.010, .749 | −0.015, .730 | −0.005, .624 |
| CMFG | 0.482 (0.410, 0.562) | 0.467 (0.442, 0.514) | 0.495 (0.479, 0.538) | 0.002, .908 | −0.002, .932 | −0.004, .558 |
| RMFG | 0.590 (0.501, 0.650) | 0.629 (0.569, 0.687) | 0.602 (0.539, 0.654) | −0.006, .875 | −0.002, .932 | 0.004, .718 |
| SFG | 0.620 (0.557, 0.671) | 0.637 (0.570, 0.714) | 0.648 (0.592, 0.711) | −0.002, .908 | −0.004, .920 | −0.002, .892 |
| PARL | 0.469 (0.389, 0.550) | 0.490 (0.453, 0.532) | 0.508 (0.465, 0.547) | −0.003, .908 | −0.006, .920 | −0.003, .720 |
| PREG | 0.489 (0.432, 0.596) | 0.506 (0.459, 0.585) | 0.527 (0.492, 0.580) | −0.003, .946 | −0.006, .920 | −0.003, .717 |
| CACC | 0.444 (0.398, 0.546) | 0.379 (0.350, 0.426) | 0.406 (0.370, 0.440) | 0.010, .749 | 0.005, .920 | −0.004, .589 |
| RACC | 0.578 (0.504, 0.620) | 0.530 (0.493, 0.577) | 0.508 (0.475, 0.565) | 0.007, .749 | 0.011, .730 | 0.003, .708 |
| PCC | 0.536 (0.395, 0.613) | 0.552 (0.509, 0.606) | 0.553 (0.505, 0.622) | −0.002, .908 | −0.002, .932 | −0.000, .959 |
| ISTH | 0.357 (0.180, 0.565) | 0.406 (0.360, 0.480) | 0.448 (0.425, 0.498) | −0.007, .908 | −0.013, .730 | −0.006, .526 |
| INS | 0.676 (0.555, 0.717) | 0.543 (0.494, 0.626) | 0.505 (0.482, 0.560) | 0.020, .703 | 0.026, .389 | 0.006, .589 |
| TPOL | 0.302 (0.000, 0.478) | 0.047 (0.000, 0.189) | 0.245 (0.226, 0.352) | 0.040, .749 | 0.009, .920 | **−0.030, .025** |
| ENTH | 0.311 (0.020, 0.451) | 0.063 (0.000, 0.288) | 0.320 (0.300, 0.377) | 0.039, .749 | −0.001, .959 | **−0.040, .018** |
| PHG | 0.442 (0.289, 0.506) | 0.371 (0.310, 0.428) | 0.434 (0.405, 0.481) | 0.011, .749 | 0.002, .932 | −0.010, .089 |
| TVTG | 0.493 (0.458, 0.570) | 0.432 (0.395, 0.509) | 0.517 (0.470, 0.547) | 0.009, .759 | −0.004, .920 | −0.013, .089 |
| ITG | 0.569 (0.486, 0.649) | 0.435 (0.392, 0.505) | 0.529 (0.471, 0.589) | 0.021, .703 | 0.006, .920 | −0.015, .112 |
| MTG | 0.527 (0.462, 0.612) | 0.525 (0.481, 0.587) | 0.493 (0.457, 0.541) | 0.000, .961 | 0.005, .920 | 0.005, .624 |
| STG | 0.626 (0.551, 0.703) | 0.553 (0.513, 0.610) | 0.538 (0.506, 0.585) | 0.011, .749 | 0.013, .730 | 0.002, .720 |
| BANKS | 0.435 (0.370, 0.503) | 0.372 (0.365, 0.419) | 0.408 (0.390, 0.448) | 0.010, .749 | 0.004, .920 | −0.005, .343 |
| FUSI | 0.614 (0.498, 0.685) | 0.574 (0.513, 0.646) | 0.574 (0.516, 0.643) | 0.006, .875 | 0.006, .920 | −0.000, .959 |

| | | | | | | |
|---|---|---|---|---|---|---|
| POSG | 0.588 (0.501, 0.660) | 0.484 (0.436, 0.563) | 0.520 (0.478, 0.567) | 0.016, .749 | 0.011, .730 | −0.006, .608 |
| PREC | 0.586 (0.478, 0.651) | 0.612 (0.530, 0.677) | 0.676 (0.632, 0.716) | −0.004, .908 | −0.014, .730 | −0.010, .308 |
| SMG | 0.501 (0.447, 0.578) | 0.428 (0.342, 0.496) | 0.520 (0.484, 0.557) | 0.011, .749 | −0.003, .920 | **−0.014, .030** |
| IPL | 0.484 (0.410, 0.574) | 0.502 (0.459, 0.570) | 0.538 (0.507, 0.585) | −0.003, .908 | −0.008, .811 | −0.006, .558 |
| SPL | 0.493 (0.406, 0.583) | 0.440 (0.389, 0.523) | 0.490 (0.447, 0.545) | 0.008, .875 | 0.001, .959 | −0.007, .589 |
| LING | 0.297 (0.218, 0.496) | 0.333 (0.308, 0.377) | 0.401 (0.372, 0.467) | −0.005, .908 | −0.015, .730 | −0.010, .070 |
| CUN | 0.316 (0.227, 0.518) | 0.328 (0.309, 0.394) | 0.446 (0.391, 0.480) | −0.001, .961 | −0.019, .728 | **−0.018, .025** |
| PERI | 0.283 (0.198, 0.439) | 0.254 (0.243, 0.321) | 0.362 (0.338, 0.392) | 0.005, .908 | −0.012, .730 | **−0.016, .012** |
| LOC | 0.386 (0.234, 0.573) | 0.452 (0.410, 0.520) | 0.478 (0.417, 0.541) | −0.010, .759 | −0.014, .730 | −0.004, .624 |
| HIPPO | 0.405 (0.324, 0.526) | 0.303 (0.190, 0.479) | 0.377 (0.335, 0.453) | 0.015, .875 | 0.004, .932 | −0.011, .520 |
| AMYG | 0.483 (0.352, 0.581) | 0.299 (0.201, 0.480) | 0.366 (0.310, 0.468) | 0.028, .749 | 0.018, .811 | −0.010, .624 |
| CAUD | 0.359 (0.234, 0.425) | 0.222 (0.160, 0.327) | 0.278 (0.254, 0.347) | 0.021, .749 | 0.012, .920 | −0.009, .473 |
| PUT | 0.476 (0.358, 0.538) | 0.279 (0.196, 0.406) | 0.346 (0.309, 0.406) | 0.030, .703 | 0.020, .730 | −0.010, .456 |
| NACC | 0.482 (0.344, 0.571) | 0.203 (0.142, 0.316) | 0.323 (0.286, 0.400) | 0.042, .652 | 0.024, .730 | −0.018, .089 |
| PALL | 0.403 (0.306, 0.515) | 0.225 (0.159, 0.327) | 0.258 (0.230, 0.291) | 0.027, .703 | 0.022, .730 | −0.005, .589 |
| THAL | 0.479 (0.320, 0.556) | 0.274 (0.189, 0.442) | 0.328 (0.290, 0.439) | 0.031, .749 | 0.023, .730 | −0.008, .608 |

Means were calculated across a range of density thresholds ($K$ = 0.10–0.25, $\Delta K$ = 0.01) and are reported with 95% bootstrap confidence intervals (CIs; 5000 resamples). Group differences in area under the curve (ΔAUC) were assessed using permutation testing (10 000 permutations). Bold values indicate statistically significant group differences after Benjamini–Hochberg correction for multiple comparisons ($p_{BH} < .05$). Abbreviations: SZ+A, schizophrenia with aggression; SZ−A, schizophrenia without aggression; HC, healthy controls; FPOL, frontal pole; POP, pars opercularis; PTRI, pars triangularis; PORB, pars orbitalis; MOFC, medial orbitofrontal cortex; LOFC, lateral orbitofrontal cortex; CMFG, caudal middle frontal gyrus; RMFG, rostral middle frontal gyrus; SFG, superior frontal gyrus; PARL, paracentral lobule; PREG, precentral gyrus; CACC, caudal anterior cingulate cortex; RACC, rostral anterior cingulate cortex; PCC, posterior cingulate cortex; ISTH, isthmus of the cingulate cortex; INS, insular cortex; TPOL, temporal pole; ENTH, entorhinal cortex; PHG, parahippocampal gyrus; TVTG, transverse temporal gyrus; ITG, inferior temporal gyrus; MTG, middle temporal gyrus; STG, superior temporal gyrus; BANKS, banks of the superior temporal sulcus; FUSI, fusiform gyrus; POSG, postcentral gyrus; PREC, precuneus; SMG, supramarginal gyrus; IPL, inferior parietal lobule; SPL, superior parietal lobule; LING, lingual gyrus; CUN, cuneus; PERI, pericalcarine cortex; LOC, lateral occipital cortex; HIPPO, hippocampus; AMYG, amygdala; CAUD, caudate nucleus; PUT, putamen; NACC, nucleus accumbens; PALL, globus pallidus; THAL, thalamus.

**Table A5.19.** Group differences in within-module degree using regional volumes residualized for CPZ-equivalent dose.

| | M (95% CI) | | | ΔAUC, $p_{BH}$ | | |
|---|---|---|---|---|---|---|
| Node | SZ+A (*N* = 212) | SZ-A (*N* = 1533) | HC (*N* = 3058) | SZ+A vs SZ-A | SZ+A vs HC | SZ-A vs HC |
| FPOL | −1.73 (−1.94, 0.00) | 0.00 (−1.63, 0.00) | −1.76 (−1.87, 0.00) | −0.258, .900 | 0.005, .968 | 0.263, .219 |
| POP | −1.51 (−1.71, 0.13) | −1.20 (−1.48, −0.11) | −1.07 (−1.46, −0.50) | −0.046, .983 | −0.068, .968 | −0.022, .934 |
| PTRI | −0.85 (−1.47, 0.56) | −0.74 (−1.04, 0.51) | −0.78 (−1.24, −0.50) | −0.017, .999 | −0.012, .968 | 0.006, .971 |
| PORB | −0.71 (−1.50, 0.12) | −0.88 (−1.16, −0.22) | −0.87 (−1.20, −0.35) | 0.026, .983 | 0.026, .968 | −0.001, .987 |
| MOFC | 1.13 (−0.26, 2.09) | 0.73 (0.05, 1.37) | 0.08 (−0.50, 0.62) | 0.061, .983 | 0.158, .753 | 0.097, .604 |
| LOFC | 0.35 (−0.50, 1.33) | 1.81 (0.89, 2.29) | 2.10 (1.13, 2.68) | −0.220, .900 | −0.266, .753 | −0.045, .918 |
| CMFG | −0.53 (−1.30, 0.60) | −0.32 (−0.76, 0.19) | −0.02 (−0.36, 0.37) | −0.034, .983 | −0.074, .968 | −0.040, .856 |
| RMFG | 1.17 (−0.31, 1.85) | 1.66 (0.78, 2.19) | 1.25 (0.38, 1.90) | −0.071, .983 | −0.008, .968 | 0.063, .918 |
| SFG | 1.71 (0.71, 2.52) | 1.74 (0.97, 2.40) | 2.22 (1.31, 2.85) | −0.006, .999 | −0.080, .968 | −0.074, .867 |
| PARL | −0.67 (−1.51, 0.45) | 0.01 (−0.50, 0.37) | −0.11 (−0.67, 0.46) | −0.102, .983 | −0.081, .968 | 0.021, .934 |
| PREG | −0.11 (−0.96, 1.11) | 0.14 (−0.52, 1.01) | 0.18 (−0.36, 0.85) | −0.037, .999 | −0.043, .968 | −0.006, .971 |
| CACC | −1.00 (−1.45, 0.20) | −1.30 (−1.49, −0.96) | −1.24 (−1.53, −0.93) | 0.043, .983 | 0.034, .968 | −0.010, .934 |
| RACC | 0.40 (−0.57, 1.14) | 0.49 (−0.09, 0.94) | −0.02 (−0.49, 0.62) | −0.015, .999 | 0.064, .968 | 0.080, .655 |
| PCC | 0.03 (−1.36, 1.04) | 0.77 (0.12, 1.38) | 0.56 (−0.13, 1.33) | −0.111, .983 | −0.077, .968 | 0.034, .934 |
| ISTH | −0.36 (−1.56, 0.77) | −1.28 (−1.55, −0.67) | −1.38 (−1.63, −0.52) | 0.140, .983 | 0.160, .830 | 0.019, .934 |
| INS | 1.93 (0.24, 2.50) | 0.51 (−0.15, 1.16) | −0.11 (−0.52, 0.48) | 0.215, .900 | 0.312, .096 | 0.097, .474 |
| TPOL | 0.00 (−1.78, 0.00) | 0.00 (−0.51, 0.00) | −0.58 (−0.61, 0.00) | 0.000, 1.000 | 0.087, .968 | 0.087, .134 |
| ENTH | 0.00 (−1.91, 0.72) | 0.00 (−1.31, 0.79) | 1.15 (0.00, 1.15) | 0.000, 1.000 | −0.173, .968 | −0.173, .134 |
| PHG | −0.75 (−1.63, 1.04) | 0.00 (−1.78, 0.00) | −0.58 (−1.04, −0.43) | −0.111, .983 | −0.024, .968 | 0.087, .134 |
| TVTG | −0.34 (−0.88, 0.70) | −0.86 (−1.24, −0.10) | −0.11 (−0.71, 0.27) | 0.078, .983 | −0.037, .968 | −0.115, .219 |
| ITG | 0.29 (−0.77, 1.58) | −0.87 (−1.27, −0.19) | −0.20 (−0.77, 0.37) | 0.175, .900 | 0.072, .968 | −0.103, .367 |
| MTG | −0.15 (−0.79, 1.03) | 0.34 (−0.24, 0.93) | −0.36 (−0.71, 0.12) | −0.075, .983 | 0.030, .968 | 0.104, .341 |
| STG | 1.52 (0.31, 2.51) | 0.74 (0.10, 1.25) | 0.31 (−0.14, 0.88) | 0.115, .983 | 0.182, .753 | 0.067, .690 |
| BANKS | −1.09 (−1.54, 0.26) | −1.22 (−1.33, −0.94) | −1.12 (−1.33, −0.85) | 0.022, .983 | 0.006, .968 | −0.016, .918 |
| FUSI | 0.09 (−0.97, 1.94) | 0.19 (−0.48, 1.02) | −0.01 (−0.77, 0.77) | −0.010, .999 | 0.017, .968 | 0.027, .934 |

| | | | | | | |
|---|---|---|---|---|---|---|
| POSG | 0.96 (−0.28, 2.01) | −0.13 (−0.74, 0.77) | −0.10 (−0.62, 0.54) | 0.166, .983 | 0.161, .753 | −0.005, .971 |
| PREC | 0.52 (−0.78, 1.55) | 1.37 (0.18, 1.97) | 1.78 (1.11, 2.51) | −0.129, .983 | −0.191, .753 | −0.062, .867 |
| SMG | −0.16 (−0.89, 0.85) | −0.83 (−1.26, −0.10) | −0.01 (−0.51, 0.48) | 0.102, .983 | −0.025, .968 | −0.127, .148 |
| IPL | −0.57 (−1.13, 0.87) | −0.07 (−0.61, 0.63) | 0.14 (−0.34, 0.68) | −0.078, .983 | −0.110, .911 | −0.032, .918 |
| SPL | −0.69 (−1.34, 0.59) | −0.81 (−1.30, 0.05) | −0.73 (−1.30, −0.01) | 0.013, .999 | 0.005, .968 | −0.008, .971 |
| LING | 0.53 (−0.87, 1.25) | 0.87 (0.00, 1.19) | 0.92 (−0.74, 1.10) | −0.050, .983 | −0.061, .968 | −0.012, .934 |
| CUN | 0.59 (−0.70, 1.26) | 0.87 (0.00, 0.93) | −0.20 (−0.63, 0.38) | −0.040, .983 | 0.123, .753 | **0.163, .020** |
| PERI | −0.03 (−1.15, 0.79) | −0.87 (−0.87, 0.00) | 0.40 (−1.07, 0.65) | 0.125, .983 | −0.068, .968 | **−0.194, .023** |
| LOC | −0.73 (−1.55, 0.58) | −0.87 (−0.93, 0.16) | 0.26 (−0.40, 0.62) | 0.017, .996 | −0.153, .753 | −0.170, .075 |
| HIPPO | −0.41 (−1.21, 1.05) | −0.79 (−1.14, 1.11) | −0.31 (−0.80, 0.37) | 0.057, .983 | −0.019, .968 | −0.077, .474 |
| AMYG | 0.89 (−0.79, 1.41) | −0.16 (−0.57, 0.62) | −0.18 (−0.45, 0.46) | 0.158, .900 | 0.161, .753 | 0.003, .971 |
| CAUD | −0.94 (−1.67, 0.54) | 0.00 (−0.37, 0.87) | −0.13 (−0.48, 0.36) | −0.143, .900 | −0.119, .753 | 0.023, .918 |
| PUT | 1.55 (−0.50, 1.92) | 1.74 (0.87, 1.92) | 1.59 (1.27, 1.84) | −0.025, .999 | −0.006, .968 | 0.020, .918 |
| NACC | −0.50 (−1.40, 1.08) | −1.18 (−1.38, −0.49) | 0.46 (−0.05, 0.81) | 0.101, .983 | −0.144, .753 | **−0.245, .002** |
| PALL | 0.32 (−0.96, 1.45) | −0.23 (−0.87, 0.24) | −1.45 (−1.73, −1.03) | 0.080, .983 | 0.269, .096 | **0.189, .002** |
| THAL | −0.16 (−1.32, 0.98) | 0.62 (−0.57, 1.06) | 0.02 (−0.82, 0.45) | −0.117, .983 | −0.031, .968 | 0.086, .803 |

Means were calculated across a range of density thresholds ($K$ = 0.10–0.25, $\Delta K$ = 0.01) and are reported with 95% bootstrap confidence intervals (CIs; 5000 resamples). Group differences in area under the curve (ΔAUC) were assessed using permutation testing (10 000 permutations). Bold values indicate statistically significant group differences after Benjamini–Hochberg correction for multiple comparisons ($p_{BH} < .05$). Abbreviations: SZ+A, schizophrenia with aggression; SZ−A, schizophrenia without aggression; HC, healthy controls; FPOL, frontal pole; POP, pars opercularis; PTRI, pars triangularis; PORB, pars orbitalis; MOFC, medial orbitofrontal cortex; LOFC, lateral orbitofrontal cortex; CMFG, caudal middle frontal gyrus; RMFG, rostral middle frontal gyrus; SFG, superior frontal gyrus; PARL, paracentral lobule; PREG, precentral gyrus; CACC, caudal anterior cingulate cortex; RACC, rostral anterior cingulate cortex; PCC, posterior cingulate cortex; ISTH, isthmus of the cingulate cortex; INS, insular cortex; TPOL, temporal pole; ENTH, entorhinal cortex; PHG, parahippocampal gyrus; TVTG, transverse temporal gyrus; ITG, inferior temporal gyrus; MTG, middle temporal gyrus; STG, superior temporal gyrus; BANKS, banks of the superior temporal sulcus; FUSI, fusiform gyrus; POSG, postcentral gyrus; PREC, precuneus; SMG, supramarginal gyrus; IPL, inferior parietal lobule; SPL, superior parietal lobule; LING, lingual gyrus; CUN, cuneus; PERI, pericalcarine cortex; LOC, lateral occipital cortex; HIPPO, hippocampus; AMYG, amygdala; CAUD, caudate nucleus; PUT, putamen; NACC, nucleus accumbens; PALL, globus pallidus; THAL, thalamus.

**Table A5.20.** Group differences in participation coefficient using regional volumes residualized for CPZ-equivalent dose.

| | M (95% CI) | | | ΔAUC, $p_{BH}$ | | |
|---|---|---|---|---|---|---|
| Node | SZ+A (*N* = 212) | SZ-A (*N* = 1533) | HC (*N* = 3058) | SZ+A vs SZ-A | SZ+A vs HC | SZ-A vs HC |
| FPOL | 0.00 (0.00, 0.18) | 0.00 (0.00, 0.00) | 0.00 (0.00, 0.00) | 0.000, 1.000 | 0.000, 1.000 | 0.000, 1.000 |
| POP | 0.00 (0.00, 0.35) | 0.00 (0.00, 0.18) | 0.00 (0.00, 0.00) | 0.000, 1.000 | 0.000, 1.000 | 0.000, 1.000 |
| PTRI | 0.00 (0.00, 0.46) | 0.00 (0.00, 0.41) | 0.00 (0.00, 0.00) | 0.000, 1.000 | 0.000, 1.000 | 0.000, 1.000 |
| PORB | 0.00 (0.00, 0.37) | 0.00 (0.00, 0.47) | 0.02 (0.00, 0.16) | 0.000, 1.000 | −0.001, .899 | −0.001, .921 |
| MOFC | 0.00 (0.00, 0.41) | 0.01 (0.00, 0.09) | 0.01 (0.00, 0.11) | −0.000, .998 | −0.001, 1.000 | −0.000, 1.000 |
| LOFC | 0.02 (0.00, 0.41) | 0.00 (0.00, 0.20) | 0.06 (0.00, 0.11) | 0.003, 1.000 | −0.006, 1.000 | −0.009, 1.000 |
| CMFG | 0.04 (0.00, 0.28) | 0.00 (0.00, 0.00) | 0.00 (0.00, 0.07) | 0.005, .538 | 0.005, .886 | 0.000, 1.000 |
| RMFG | 0.00 (0.00, 0.37) | 0.06 (0.00, 0.22) | 0.09 (0.00, 0.18) | −0.010, .867 | −0.013, .899 | −0.003, 1.000 |
| SFG | 0.04 (0.00, 0.21) | 0.08 (0.00, 0.20) | 0.00 (0.00, 0.10) | −0.006, 1.000 | 0.005, .935 | 0.011, .921 |
| PARL | 0.00 (0.00, 0.21) | 0.00 (0.00, 0.00) | 0.00 (0.00, 0.00) | 0.000, 1.000 | 0.000, 1.000 | 0.000, 1.000 |
| PREG | 0.00 (0.00, 0.24) | 0.00 (0.00, 0.06) | 0.00 (0.00, 0.08) | 0.000, 1.000 | 0.000, 1.000 | 0.000, 1.000 |
| CACC | 0.08 (0.00, 0.44) | 0.00 (0.00, 0.00) | 0.00 (0.00, 0.00) | 0.011, .403 | 0.011, .886 | 0.000, 1.000 |
| RACC | 0.18 (0.00, 0.49) | 0.00 (0.00, 0.02) | 0.02 (0.00, 0.12) | 0.026, .403 | 0.024, .886 | −0.002, .921 |
| PCC | 0.01 (0.00, 0.45) | 0.00 (0.00, 0.09) | 0.00 (0.00, 0.10) | 0.001, .867 | 0.001, 1.000 | 0.000, 1.000 |
| ISTH | 0.33 (0.00, 0.56) | 0.12 (0.00, 0.40) | 0.49 (0.00, 0.50) | 0.032, .867 | −0.023, 1.000 | −0.055, .201 |
| INS | 0.18 (0.00, 0.41) | 0.02 (0.00, 0.22) | 0.00 (0.00, 0.09) | 0.023, .844 | 0.026, .887 | 0.002, 1.000 |
| TPOL | 0.38 (0.00, 0.59) | 0.00 (0.00, 0.12) | 0.00 (0.00, 0.38) | 0.057, .448 | 0.057, .886 | 0.000, 1.000 |
| ENTH | 0.41 (0.00, 0.57) | 0.09 (0.00, 0.26) | 0.00 (0.00, 0.50) | 0.049, .448 | 0.062, .886 | 0.012, .921 |
| PHG | 0.47 (0.00, 0.60) | 0.34 (0.00, 0.59) | 0.66 (0.60, 0.72) | 0.018, 1.000 | −0.028, .936 | **−0.047, .012** |
| TVTG | 0.00 (0.00, 0.26) | 0.00 (0.00, 0.08) | 0.00 (0.00, 0.02) | 0.000, 1.000 | 0.000, 1.000 | 0.000, 1.000 |
| ITG | 0.20 (0.00, 0.62) | 0.00 (0.00, 0.07) | 0.05 (0.00, 0.24) | 0.030, .772 | 0.023, .899 | −0.007, 1.000 |
| MTG | 0.12 (0.00, 0.59) | 0.00 (0.00, 0.09) | 0.00 (0.00, 0.10) | 0.018, .772 | 0.018, .899 | 0.000, 1.000 |
| STG | 0.08 (0.00, 0.37) | 0.00 (0.00, 0.10) | 0.00 (0.00, 0.08) | 0.012, .863 | 0.012, .899 | 0.000, 1.000 |
| BANKS | 0.00 (0.00, 0.46) | 0.00 (0.00, 0.00) | 0.00 (0.00, 0.00) | 0.000, 1.000 | 0.000, 1.000 | 0.000, 1.000 |
| FUSI | 0.49 (0.00, 0.68) | 0.35 (0.14, 0.47) | 0.33 (0.22, 0.45) | 0.022, .998 | 0.024, .899 | 0.001, 1.000 |

| | | | | | | |
|---|---|---|---|---|---|---|
| POSG | 0.10 (0.00, 0.40) | 0.00 (0.00, 0.02) | 0.00 (0.00, 0.11) | 0.014, .772 | 0.014, .887 | 0.000, 1.000 |
| PREC | 0.13 (0.00, 0.49) | 0.08 (0.00, 0.22) | 0.26 (0.00, 0.33) | 0.007, 1.000 | −0.020, 1.000 | −0.027, .367 |
| SMG | 0.00 (0.00, 0.24) | 0.00 (0.00, 0.00) | 0.00 (0.00, 0.05) | 0.000, 1.000 | 0.000, 1.000 | 0.000, 1.000 |
| IPL | 0.00 (0.00, 0.53) | 0.00 (0.00, 0.10) | 0.00 (0.00, 0.14) | 0.000, 1.000 | 0.000, 1.000 | 0.000, 1.000 |
| SPL | 0.17 (0.00, 0.44) | 0.00 (0.00, 0.19) | 0.32 (0.00, 0.43) | 0.027, .844 | −0.022, 1.000 | −0.049, .358 |
| LING | 0.00 (0.00, 0.55) | 0.07 (0.00, 0.45) | 0.11 (0.00, 0.52) | −0.009, 1.000 | −0.015, 1.000 | −0.006, 1.000 |
| CUN | 0.10 (0.00, 0.49) | 0.00 (0.00, 0.45) | 0.46 (0.00, 0.48) | 0.014, 1.000 | −0.056, .899 | **−0.070, .014** |
| PERI | 0.00 (0.00, 0.35) | 0.00 (0.00, 0.00) | 0.00 (0.00, 0.19) | 0.000, 1.000 | 0.000, 1.000 | 0.000, 1.000 |
| LOC | 0.33 (0.00, 0.58) | 0.44 (0.40, 0.52) | 0.43 (0.00, 0.51) | −0.015, .867 | −0.016, 1.000 | −0.001, 1.000 |
| HIPPO | 0.05 (0.00, 0.51) | 0.31 (0.04, 0.55) | 0.35 (0.03, 0.48) | −0.040, .867 | −0.046, .886 | −0.006, 1.000 |
| AMYG | 0.27 (0.00, 0.59) | 0.18 (0.02, 0.56) | 0.18 (0.00, 0.42) | 0.016, 1.000 | 0.016, 1.000 | −0.000, 1.000 |
| CAUD | 0.00 (0.00, 0.38) | 0.00 (0.00, 0.12) | 0.00 (0.00, 0.00) | 0.000, 1.000 | 0.000, 1.000 | 0.000, 1.000 |
| PUT | 0.27 (0.00, 0.54) | 0.00 (0.00, 0.42) | 0.00 (0.00, 0.11) | 0.041, .867 | 0.041, .899 | 0.000, 1.000 |
| NACC | 0.48 (0.00, 0.61) | 0.00 (0.00, 0.08) | 0.00 (0.00, 0.15) | 0.071, .403 | 0.071, .886 | 0.000, 1.000 |
| PALL | 0.03 (0.00, 0.48) | 0.00 (0.00, 0.33) | 0.00 (0.00, 0.00) | 0.004, .867 | 0.004, .886 | 0.000, 1.000 |
| THAL | 0.35 (0.00, 0.57) | 0.02 (0.00, 0.47) | 0.00 (0.00, 0.26) | 0.051, .867 | 0.052, .899 | 0.001, 1.000 |

Means were calculated across a range of density thresholds ($K$ = 0.10–0.25, $\Delta K$ = 0.01) and are reported with 95% bootstrap confidence intervals (CIs; 5000 resamples). Group differences in area under the curve (ΔAUC) were assessed using permutation testing (10 000 permutations). Bold values indicate statistically significant group differences after Benjamini–Hochberg correction for multiple comparisons ($p_{BH} < .05$). Abbreviations: SZ+A, schizophrenia with aggression; SZ−A, schizophrenia without aggression; HC, healthy controls; FPOL, frontal pole; POP, pars opercularis; PTRI, pars triangularis; PORB, pars orbitalis; MOFC, medial orbitofrontal cortex; LOFC, lateral orbitofrontal cortex; CMFG, caudal middle frontal gyrus; RMFG, rostral middle frontal gyrus; SFG, superior frontal gyrus; PARL, paracentral lobule; PREG, precentral gyrus; CACC, caudal anterior cingulate cortex; RACC, rostral anterior cingulate cortex; PCC, posterior cingulate cortex; ISTH, isthmus of the cingulate cortex; INS, insular cortex; TPOL, temporal pole; ENTH, entorhinal cortex; PHG, parahippocampal gyrus; TVTG, transverse temporal gyrus; ITG, inferior temporal gyrus; MTG, middle temporal gyrus; STG, superior temporal gyrus; BANKS, banks of the superior temporal sulcus; FUSI, fusiform gyrus; POSG, postcentral gyrus; PREC, precuneus; SMG, supramarginal gyrus; IPL, inferior parietal lobule; SPL, superior parietal lobule; LING, lingual gyrus; CUN, cuneus; PERI, pericalcarine cortex; LOC, lateral occipital cortex; HIPPO, hippocampus; AMYG, amygdala; CAUD, caudate nucleus; PUT, putamen; NACC, nucleus accumbens; PALL, globus pallidus; THAL, thalamus.

**Table A5.21.** Group differences in canonical brain network density using regional volumes residualized for CPZ-equivalent dose.

| Metric | M (95% CI) | | | ΔAUC, $p_{BH}$ | | |
|---|---|---|---|---|---|---|
| | SZ+A (*N* = 212) | SZ-A (*N* = 1533) | HC (*N* = 3058) | SZ+A vs SZ-A | SZ+A vs HC | SZ-A vs HC |
| *Salience network* | | | | | | |
| Network-rest | 0.28 (0.19, 0.33) | 0.19 (0.15, 0.24) | 0.14 (0.12, 0.18) | 0.014, .292 | **0.021, .027** | 0.007, .172 |
| *Central executive network* | | | | | | |
| Within-network | 0.34 (0.05, 0.55) | 0.55 (0.35, 0.66) | 0.29 (0.17, 0.51) | −0.032, .960 | 0.009, 1.000 | 0.041, .377 |
| Network-rest | 0.18 (0.14, 0.24) | 0.23 (0.19, 0.26) | 0.24 (0.21, 0.26) | −0.007, .403 | −0.009, .358 | −0.002, .608 |
| *Default mode network* | | | | | | |
| Within-network | 0.14 (0.10, 0.21) | 0.17 (0.14, 0.20) | 0.18 (0.15, 0.20) | −0.004, .960 | −0.006, 1.000 | −0.002, .872 |
| Network-rest | 0.15 (0.13, 0.17) | 0.15 (0.14, 0.17) | 0.15 (0.14, 0.16) | 0.000, .882 | 0.001, .970 | 0.001, .608 |
| *Language network* | | | | | | |
| Within-network | 0.34 (0.24, 0.52) | 0.35 (0.30, 0.40) | 0.31 (0.30, 0.38) | −0.002, 1.000 | 0.004, 1.000 | 0.006, .872 |
| Network-rest | 0.16 (0.13, 0.21) | 0.19 (0.16, 0.22) | 0.16 (0.14, 0.18) | −0.005, .512 | −0.000, .970 | 0.005, .172 |
| *Reward network* | | | | | | |
| Within-network | 0.39 (0.26, 0.48) | 0.32 (0.25, 0.38) | 0.39 (0.33, 0.43) | 0.011, .960 | 0.000, 1.000 | −0.010, .377 |
| Network-rest | 0.15 (0.12, 0.17) | 0.16 (0.14, 0.18) | 0.14 (0.12, 0.15) | −0.003, .528 | 0.001, .970 | 0.004, .172 |
| *Affective control network* | | | | | | |
| Within-network | 0.32 (0.25, 0.38) | 0.33 (0.28, 0.37) | 0.32 (0.28, 0.34) | −0.001, 1.000 | 0.000, 1.000 | 0.002, .872 |
| Network-rest | 0.14 (0.13, 0.17) | 0.17 (0.15, 0.17) | 0.14 (0.14, 0.15) | −0.003, .270 | 0.000, .970 | **0.003, .026** |

Means were calculated across a range of density thresholds ($K$ = 0.10–0.25, $\Delta K$ = 0.01) and are reported with 95% bootstrap confidence intervals (CIs; 5000 resamples). Group differences in area under the curve (ΔAUC) were assessed using permutation testing (10 000 permutations). Bold values indicate statistically significant group differences after Benjamini–Hochberg correction for multiple comparisons ($p_{BH} < .05$). Abbreviations: SZ+A, schizophrenia with aggression; SZ−A, schizophrenia without aggression; HC, healthy controls.

### Appendix 6
References

93. Menon V. 20 years of the default mode network: a review and synthesis. Neuron. 2023;111(16):2469-87.
94. Hickok G, Poeppel D. The cortical organization of speech processing. Nat Rev Neurosci. 2007;8(5):393-402.
95. Bartra O, McGuire JT, Kable JW. The valuation system: a coordinate-based meta-analysis of BOLD fMRI experiments examining neural correlates of subjective value. Neuroimage. 2013;76:412-27.
96. Kesby JP, Murray GK, Knolle F. Neural circuitry of salience and reward processing in psychosis. Biol Psychiatry Glob Open Sci. 2023;3(1):33-46.
